# Synchrony between brain age and internalising and externalising symptoms across adolescence

**DOI:** 10.64898/2025.12.31.25343265

**Authors:** Dani Beck, Chloe Carrick, Eira R. Aksnes, Niamh MacSweeney, Lars T. Westlye, Delia Fuhrmann, Christian K. Tamnes

## Abstract

Adolescence is a period of rapid neurobiological and behavioural change, yet it remains unclear how deviations from normative brain maturation relate to psychopathology. Using data from the Adolescent Brain Cognitive Development Study, we combined brain age prediction with bivariate latent growth curve models to examine whether brain maturation deviations relate to mental health across late childhood and adolescence. Brain age was estimated from T1-weighted, diffusion, resting-state fMRI, and multimodal MRI across four waves (ages ∼8.3–17.5); symptoms were assessed across ten waves using the Brief Problem Monitor. Across T1, dMRI, and multimodal models, brain maturation deviations and symptoms showed coordinated nonlinear development. Adolescents whose brains increasingly diverged from age-expected maturation also showed increasing symptom trajectories. Effects were small to moderate, most consistent for internalising symptoms in females (r = .15–.23), while externalising showed broader nonlinear associations in both sexes (r = .15–.32). Intercept-level associations were weaker, less consistent, and limited to externalising in specific modalities (r = .08–.09). Formal tests found no robust sex differences after multiple comparison correction. These results suggest vulnerability to adolescent psychopathology is more strongly linked to nonlinear changes in brain maturational tempo than to fixed differences in brain age.

## 1. Introduction

Internalising and externalising symptoms in childhood and adolescence predict later mental health problems^1^, and have been linked to a wide range of neurobiological alterations in youth^2^. Several neuroimaging studies have reported attenuated or delayed cortical maturation in youth with elevated symptoms, including reduced thinning or slower surface-area development^2–9^. Conversely, other work finds accelerated cortical change, including increased thinning or volume loss^3,10,11^. Evidence from subcortical^3,12^ and white matter development^13,14^ is similarly mixed, ranging from attenuated to accelerated maturational trajectories, and with some studies reporting no robust associations^15^. Functional neuroimaging findings show altered responsivity in affective and reward circuits accompanying elevated symptoms and, in some cases, preceding later symptom progression^16–21^.

Adolescence is also a period of rising sex differences in internalising and externalising psychopathology: internalising symptoms – including anxiety, depression, and somatic complaints – show a marked female predominance that emerges during early adolescence and peaks in mid-adolescence, whereas externalising symptoms – including attention problems, aggression, and rule-breaking behaviour – tend to be more prevalent in males, particularly for conduct and attention-related difficulties^22–24^. These diverging patterns coincide with sex differences in the timing and tempo of normative brain maturation^2,25^. Together, these findings suggest that deviations from normative brain maturation trajectories – whether accelerated or delayed – may be centrally implicated in the emergence of youth psychopathology^26^.

From a developmental perspective, both accelerated and delayed patterns of brain maturation may confer risk through altered developmental timing. In particular, risk may emerge when neural systems develop out of synchrony with each other or with environmental and socio-emotional demands, potentially increasing vulnerability during sensitive periods of development^27^. Neurodevelopment is highly heterochronous throughout childhood and adolescence, with regional and network-specific trajectories unfolding at different rates and with different sensitivity to environmental and biological influences^28,29^. As a result, deviations between chronological age and neurobiological maturity are not inherently pathological but may reflect individual differences in the timing and coordination of developmental processes. A “more mature” brain at a given chronological age may therefore index earlier maturation of specific systems, altered pruning or myelination schedules, or shifts in functional integration. Such changes may interact with emotional and behavioural regulation during development.

Several theoretical frameworks suggest that such maturational deviations could be relevant for psychopathology. The stress-acceleration model proposes that early adversity or heightened environmental demands may hasten aspects of neurobiological development^30^, potentially conferring short-term adaptive advantages but increasing vulnerability to internalising or externalising problems when regulatory systems mature out of synchrony. Relatedly, developmental mismatch accounts emphasise that earlier maturation of affective or motivational systems relative to cognitive control networks may bias behaviour toward heightened emotional reactivity or impulsivity^31^. From such perspectives, deviation from age-expected brain development may capture individual differences in developmental timing that intersect with known pathways to internalising and externalising symptoms across adolescence.

One approach to quantify maturational deviations involves brain age prediction models, which estimate an individual’s age from multivariate magnetic resonance imaging (MRI) features^32,33^. The brain age gap (BAG) – predicted minus chronological age – provides an interpretable proxy for individual differences in neurodevelopmental timing, with positive BAG values indicating an older-appearing brain relative to age and negative values indicating a younger-appearing brain. In adults, BAG has been linked to multiple contextual factors and outcomes, including socioeconomic status^34^, cardiometabolic risk and lifestyle factors^35–41^, cognition^42^, and mental health and brain disorders^43–46^. In youth, deviations from age-expected brain maturation have also been associated with several mental health profiles. Negative BAG values have been linked to higher anxiety and depression symptoms, attention difficulties, autism spectrum traits, and elevated internalising and externalising symptom scores^47–52^. Positive BAG values, in contrast, have been associated with greater depressive and obsessive–compulsive symptoms, higher general psychopathology, and increased psychosis risk in clinically vulnerable youth^53–56^. Longitudinal evidence, although limited, suggests that BAG trajectories may carry prognostic significance for later mental health outcomes^57,58^.

Taken together, discrepant findings suggest that BAG–psychopathology associations in youth may be developmentally heterogeneous, potentially depending on when brain maturation is assessed, which MRI modalities are indexed, and whether analyses capture static deviation or change over time. Notably, associations have been reported at both positive and negative BAG values for some symptom domains, potentially indicating that deviation from age-expected brain maturation – rather than acceleration or delay in a single direction – may be relevant for psychopathology risk in youth. Because cross-sectional BAG estimates conflate stable between-person differences with time-varying deviations, they cannot determine whether symptoms relate to persistent offsets, coordinated change, or divergence across development^59^. Addressing these gaps requires longitudinal, multimodal designs and modelling approaches that can characterise nonlinear, person-specific developmental dynamics and clarify whether brain–behaviour coupling reflects persistent offsets, coordinated change, or divergence across development.

To address these challenges, we leverage the Adolescent Brain Cognitive Development (ABCD) Study (release 6.0) to build T1-weighted, diffusion MRI (dMRI), and resting-state functional (rs-fMRI) brain age models, as well as a multimodal model combining features from all three modalities. Focusing on internalising and externalising symptoms as broad, transdiagnostic dimensions with well-characterised developmental trajectories, we used the cohort’s four MRI waves and ten Brief Problem Monitor (BPM) waves to examine how developmental trajectories in BAG relate to trajectories of self-reported internalising and externalising symptoms. Using bivariate latent growth curve (BLGC) models, we examined (i) individual differences in BAG at the midpoint of the observation window, as well as linear and quadratic rates of BAG change across late childhood and adolescence and (ii) the degree to which these developmental components co-vary with the corresponding growth components of internalising and externalising symptoms.

We hypothesised that trajectories of BAG and internalising and externalising symptoms would be developmentally coupled across adolescence, such that individual differences in change over time in BAG would be associated with corresponding differences in symptom trajectories. Given mixed prior evidence linking both accelerated and delayed patterns of brain maturation to youth psychopathology, and the relative scarcity of longitudinal BAG studies, we did not specify a directional hypothesis regarding whether older- or younger-appearing brains would be associated with more adverse symptom trajectories. We further tested whether the magnitude and form of BAG–symptom coupling differed by sex, motivated by established sex differences in the prevalence and development of internalising and externalising symptoms and in normative brain maturation across adolescence^24,25^. By combining multimodal brain age prediction with longitudinal growth-curve modelling, this study addresses a key gap in understanding how the tempo of brain maturation co-evolves with internalising and externalising symptom trajectories across adolescence.

## 2. Methodology

### 2.1. Sample and ethical approvals

The Adolescent Brain Cognitive Development (ABCD) Study ®^60^ is an ongoing longitudinal study comprising ∼11,800 children and adolescents. Participants were recruited from 21 sites across the United States via outreach to caregivers through local schools, followed by telephone screening to determine eligibility. Participants were excluded using the ABCD Study exclusion criteria listed elsewhere^61^. Data used in the present study were drawn from the ABCD Study release 6.0, containing data from baseline up until the six-year visit (nbdc-datahub.org/abcd-release-6-0). All ABCD Study data is stored in the NIH Brain Development Cohorts (NBDC) Data Hub, which is available for authorised users with approved Data Use Certification (DUC) (lead investigator: Westlye). The 6.0 release has been assigned the DOI <u>10.82525/jy7n-g441</u>. The Institutional Review Board (IRB) at the University of California, San Diego, approved all aspects of the ABCD Study^62^. Parents or guardians provided written consent, while the child provided written assent. The current study was conducted in line with the Declaration of Helsinki and was approved by the Norwegian Regional Committee for Medical and Health Research Ethics (REK 2019/943).

### 2.2. Demographic information and data quality assurance

For each modality, the full sample was divided into training (50%) and hold-out test (50%) sets prior to analysis, with all observations from a given individual assigned to the same set; the sample sizes reported below refer to the hold-out test set used in all subsequent analyses. T1 included 14,838 observations (male = 7,907 [53.3%], female = 6,931 [46.7%], unique N = 5866), dMRI 12,975 (male = 6,853 [52.8%], female = 6,122 [47.2%], unique N = 5112), rs-fMRI 12,990 (male = 6,931 [53.4%], female = 6,059 [46.6%], unique N = 5562), and multimodal 11,770 (male = 6139 [52.2%], female = 5,631 [47.8%], unique N = 4934). The overall mean age was ∼12.3 years (SD ≈ 2.3; range ≈ 8.3–17.7). These MRI observations were then linked to all available mental health BPM assessments for the same individuals, with up to ten data waves. Demographic information for each brain MRI modality-specific test sample can be found in SI Table 1. Comparable distributions for the brain age prediction training sets can be found SI Table 2, while the full analytic sample is illustrated in Figure 1.

**Figure 1.**
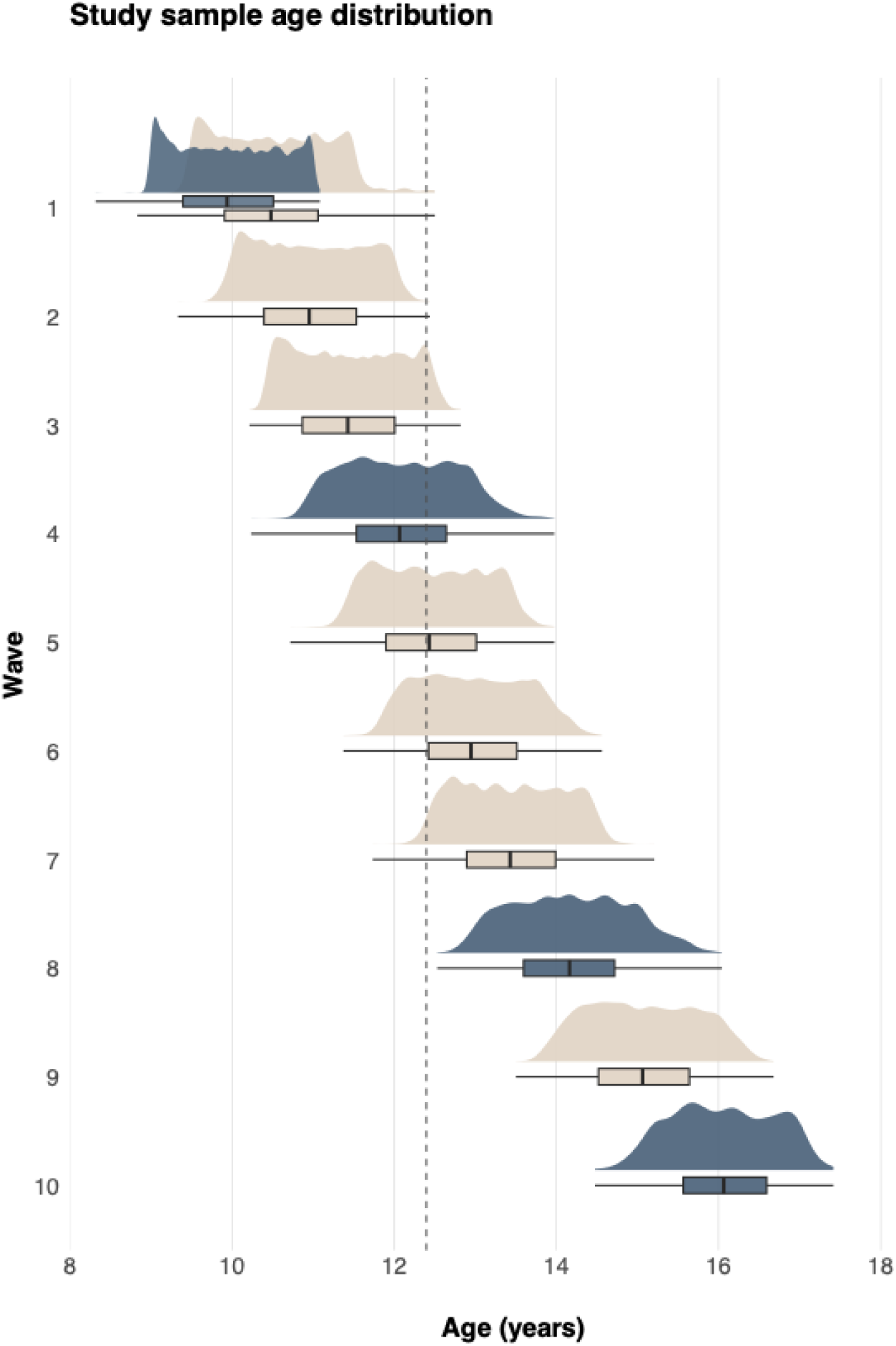
Age distributions across MRI and BPM waves for the analytic sample. Density plots and boxplots show participant age distributions across the ten assessment waves. At Wave 1, two distributions are shown because MRI Wave 1 (ses-00A) and BPM Wave 1 (ses-00M) occurred at different visits. Waves 2–10 correspond to session identifier: ses-01A, ses-01M, ses-02A, ses-02M, ses-03A, ses-03M, ses-04A, ses-05A, and ses-06A, respectively. Beige densities/boxplots represent BPM-only waves, whereas blue indicates waves in which participants provided both BPM and MRI data at the same ABCD session (except wave 1). Boxplots summarise the median and interquartile range, and density plots show the full distributional shape.

### 2.3. MRI acquisition and processing

Neuroimaging data were acquired at 21 different sites (using 34 scanners) and processed by the ABCD Study team. A 3-T Siemens Prisma, General Electric 750, or Philips scanner was used for data acquisition. Protocols used for data acquisition and processing are described in detail elsewhere^60,63^ and described in SI Section 1. Three modalities of brain structural and functional measures were used in the present study: structural grey matter measures (T1), white matter microstructural measures from diffusion tensor imaging (DTI), and resting-state functional connectivity measures (rs-fMRI)^63^. Cortical surface reconstruction and subcortical segmentation was performed with FreeSurfer v7.1.1^64,65^. White matter microstructural measures were generated using AtlasTrack, a probabilistic atlas-based method for automated segmentation of white matter fibre tracts^66^. Measures of functional connectivity were computed using a seed-based, correlational approach^67^, where average time courses were calculated for cortical surface-based ROIs using a functionally-defined parcellation based on resting-state functional connectivity patterns^68^ and subcortical ROIs^65^.

For T1, we extracted tabulated total and regional measures of cortical surface area, thickness, volume, sulcal depth, and intensity grey/white matter contrast, as well as subcortical volume (397 measures). For DTI, full and inner (multi) shell tissue properties including fractional anisotropy (FA) and mean (MD), longitudinal (or axial, AD), and transverse (or radial, RD) diffusivity were extracted for total and regional features (576 measures). For rs-fMRI, functional connectivity within and between parcellations from the Gordon network, including subcortical data,^68^ were extracted (416 measures). MRI data underwent quality assurance (QA) per standard procedures outlined elsewhere^63^, and detailed in SI Section 1. Briefly, participants with excessive head motion or poor data quality were excluded from the curated data release, in additional to removal of data using the recommendations for data cleaning provided by the ABCD Study team. Following QA procedures, harmonisation of multi-scanner effects was carried out using *longComBat*^69^ (see SI Section 2 and SI Figures 1-3).

### 2.4. Brain age prediction

Brain age prediction was carried out using the eXtreme Gradient Boosting (XGBoost) regression model^70^. Parameters were tuned using five-fold cross-validation (stratified by age) with two-repeats to average out partition noise. The models were fitted using the best estimators and optimised models were applied to the (hold-out) test sample. R^2^, RMSE, and MAE were calculated to evaluate prediction accuracy in the test set. For each brain modality (T1, DTI, rs-fMRI, and multimodal; combining features from all three modalities), 50% of the data was used as the hold-out test sample and 50% was used for model training and validation, with all observations from a given individual assigned to the same split.

Consistent with recent ABCD brain age work^58,71,72^, confounding effects from complex family-related factors were minimised using a group shuffle split with family ID as the group indicator to ensure that siblings were not split across training and test sets. The brain age model used for the study is publicly available: https://github.com/dani-beck/brainage-prediction. A detailed overview of T1, DTI, rs-fMRI, and multimodal training and test samples, including demographic information, are provided in SI Section 3 and SI Tables 1 and 2. Complete lists of all the extracted measures used for brain age prediction per modality are provided in SI Tables 3-5. Feature importance scores for each model are provided in SI Figure 4. To adjust for commonly observed age-bias (overestimated predictions for younger participants and underestimated predictions for older participants)^73^, we applied a statistical correction previously described^74^ that involves fitting a linear regression of predicted age on chronological age and using the residuals. This is visualised in SI Figure 5. The difference between an individual’s predicted brain age and their chronological age (BAG) was calculated (BAG = predicted age - chronological age) for each of the models, providing T1, DTI, rs-fMRI, and multimodal-based BAG values for all participants. This procedure, including age-bias correction of raw BAG values and evaluation of model performance in an independent hold-out sample, is consistent with methodological recommendations for brain age gap estimation^59,75^.

### 2.5. Brief Problem Monitor measures

Youth internalising and externalising problems were measured using the self-report version of the Brief Problem Monitor (BPM)^76^. The BPM is a brief 19-item instrument derived from the longer Child Behaviour Checklist (CBCL) for ages 6-18^77^, Teacher’s Report Form, and Youth Self-Report^76^, and is designed for repeated monitoring of emotional and behavioural functioning. The BPM was selected over the parent-reported CBCL, which is also collected in ABCD, because it was administered at up to ten longitudinal waves in ABCD release 6.0 compared with approximately four waves for the CBCL, providing substantially greater temporal resolution for modelling symptom trajectories within the BLGC framework. Prior work indicates that the BPM has good internal consistency (Cronbach’s α = .78–.91 across scales) and strong convergent validity with the full CBCL (total score *r* = .95; subscale correlations *r* = .86–.97), supporting its reliability and validity as a brief assessment tool in community and clinical samples^78^. Each item is rated as *0 = not true, 1 = somewhat true, or 2 = very true* over the past week, and sum scores form subscales for internalising and externalising (and attention) problems. In the present study, ten measurement waves were available: 6 months after baseline, followed by semi-annual collection until the final two waves which were annual. For information pertaining to quality control and missing data, see SI Section 4 and SI Figure 6.

### 2.6. Statistical analysis

All analyses were carried out using R version 4.3.2^79^. Longitudinal associations between brain-age gap (BAG) trajectories and internalising and externalising symptom development were examined using bivariate latent growth curve (BLGC) models estimated in *lavaan* (version 0.6-20)^80,81^. Latent growth curve models capture development by treating its model parameters (i.e., the intercept, slope, and where applicable quadratic) as latent variables, and using factor loadings to indicate the association of each parameter with the observed timepoints^82,83^.

For each imaging modality (T1, dMRI, rs-fMRI, multimodal), we compared linear, latent-basis, and quadratic growth specifications and selected the most appropriate form based on model fit and convergence. Quadratic models provided a parsimonious and stable representation of nonlinear change for all modalities except rs-fMRI, for which change was adequately captured by a linear slope due to convergence instability in nonlinear specifications (see SI Section 5 for full model selection procedure). The BLGC framework was selected because it simultaneously decomposes individual differences in two parallel developmental processes into interpretable latent growth components and permits direct estimation of their covariance structure. In this framework, the intercept captures each individual’s characteristic level at the centred timepoint, the linear slope captures the overall pace and direction of change across adolescence, and the quadratic component captures whether that rate of change itself accelerates or decelerates – a distinction particularly relevant given the nonlinear developmental patterns characteristic of both brain maturation and psychopathology in adolescence. Developmental coupling was evaluated via cross-process covariances between corresponding BAG and symptom growth factors (intercept–intercept, slope–slope, and quadratic–quadratic where supported), indexing associations at the centred time point and coordinated change across the observation window. Although the multivariate growth model estimates a broader covariance structure among growth factors, interpretation focused on these corresponding BAG–symptom covariances as primary indices of developmental synchrony. Statistical significance of the BAG–symptom coupling estimates was evaluated with Benjamini–Hochberg^84^ false discovery rate (FDR) correction across all coupling tests.

All BLGC models were estimated as multigroup models by sex, using robust maximum likelihood (MLR) with full-information maximum likelihood (FIML) for missing data. Sex was handled in two stages. First, to determine whether males and females could be modelled with a shared versus sex-specific latent covariance structure, we compared freely estimated multigroup models to models in which all latent covariances were constrained equal across groups. Scaled Satorra–Bentler χ² difference tests were used to evaluate these omnibus constraints, with significant results indicating that at least one latent covariance differed by sex; this step was used to guide model specification rather than to draw conclusions about specific BAG–symptom associations. Second, we tested sex moderation of the BAG–symptom couplings using targeted likelihood-ratio tests that constrained each cross-process coupling to be equal across sex one at a time, with multiple-comparison control using FDR^84^ correction. To assess robustness of primary coupling estimates to socioeconomic confounding, a sensitivity analysis was conducted including a composite baseline measure of socioeconomic status (SES; combining parental and partner income and educational attainment) as a time-invariant covariate regressed onto all latent growth factors; results are reported in Section 3.2.4 and SI Table 11.

Model fit was assessed using the χ^2^ test, the Root Mean Square Error of Approximation (RMSEA), the Comparative Fit Index (CFI), and the Standardised Root Mean Square Residual (SRMR). Models were defined as having good fit if CFI > .97, SRMR < .05, and RMSEA <.05. Acceptable fit was defined as CFI = .95-.97, SRMR = .05-.10, and RMSEA = .05-.08^85^. We characterised standardised parameter estimates above 0.10 as small effects, between 0.20 and 0.30 as moderate, and over 0.30 as large effect sizes^86^. All analysis code for the BLGC models is publicly available at https://osf.io/dmv2f/overview.

## 3. Results

### 3.1. Brain age prediction model performance

Model performance metrics for each brain age prediction model are provided in Table 1. Training and internal validation set performance metrics are reported in SI Section 3 for comparison. The correlation between predicted and chronological age for each model is illustrated in Figure 2. SI Figures 7-10 show raw, corrected, age-binned, and per-wave distributions of corrected BAG per brain age model. SI Figure 11 shows within-person change and longitudinal stability of corrected BAG using intercept intraclass correlations (ICC), per modality. SI Figure 12 illustrates associations between raw and corrected BAG values for all modalities.

**Table 1.** Model performance metrics (*r, r*^2^*, RMSE, MAE*) for each brain age prediction model in the hold-out (test) sample.

|  | T1 | dMRI | rs-fMRI | Multimodal |
| --- | --- | --- | --- | --- |
| Pearson's $r$ | 0.83 [0.82, 0.83] | 0.86 [0.86, 0.87] | 0.69 [0.68, 0.70] | 0.88 [0.87, 0.88] |
| $r^2$ | 0.69 | 0.75 | 0.47 | 0.78 |
| RMSE | 1.28 | 1.15 | 1.67 | 1.09 |
| MAE | 1.00 | 0.90 | 1.34 | 0.85 |

**Note:** RMSE = root mean squared error, MAE = mean absolute error.

**Figure 2.**
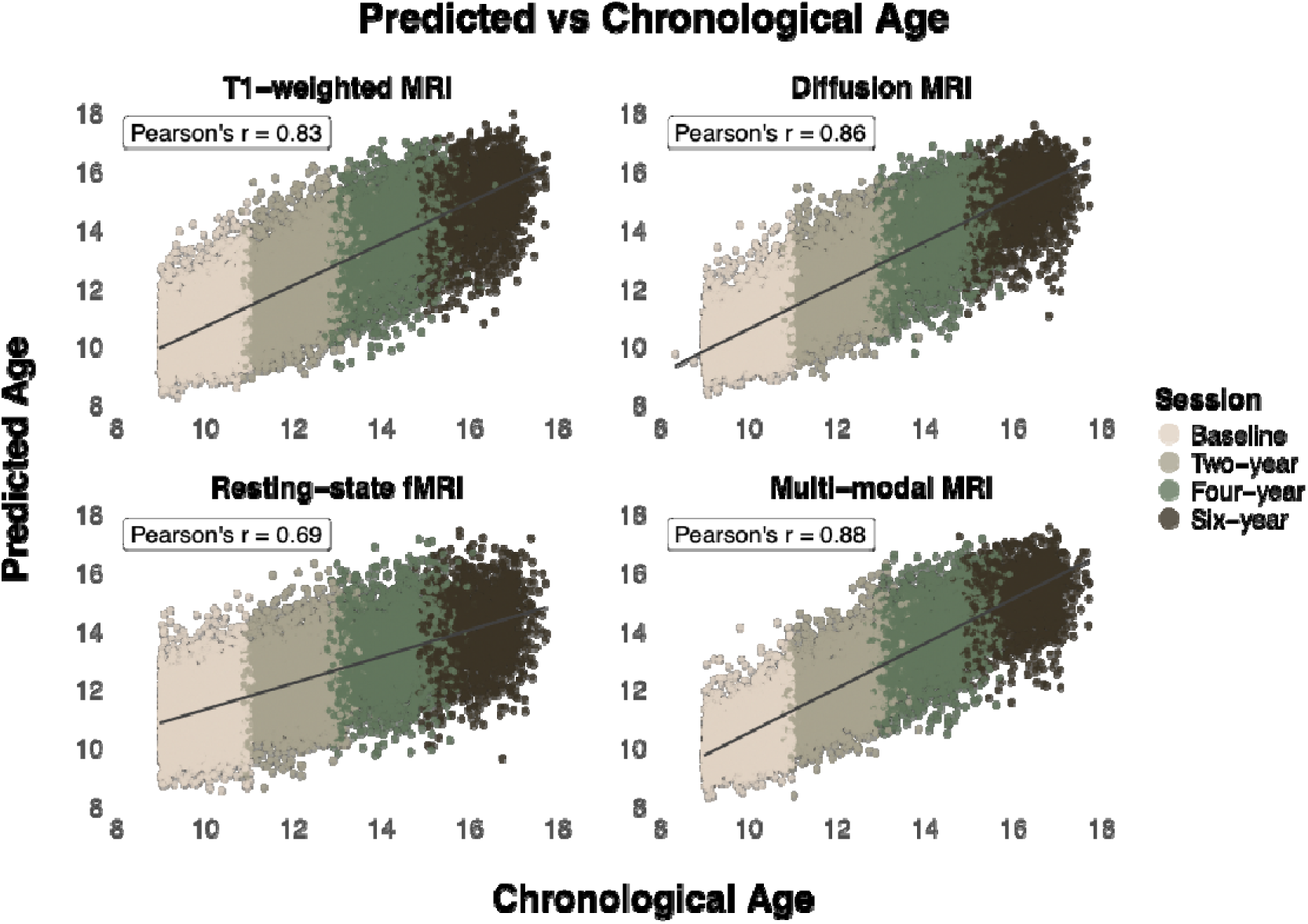
Predicted age as a function of chronological age. Scatter plots show the correlation (Pearson’s r) between predicted age and chronological age for each modality (T1, dMRI, rs-fMRI, multimodal MRI) across four waves (session).

### 3.2. Bivariate latent growth curve modelling framework and model fit

Across imaging modalities, the quadratic bivariate latent growth curve (BLGC) models provided acceptable-to-good fit (CFI = .95–.98). Full model fit indices are provided in SI Table 6. Multigroup tests indicated that latent covariances differed by sex for all modality-outcome models except dMRI–externalising, for which equality constraints were supported (SI Table 7). We therefore retained sex-stratified models for all other analyses and treated the dMRI–externalising model as sex-invariant.

All BAG–symptom associations reported below reflect standardised latent covariances from these final models. We focus interpretation on the corresponding cross-process covariances (intercept–intercept, slope–slope, and quadratic–quadratic, where applicable) as primary parameters of interest. Intercept–intercept covariances reflect associations between BAG and symptom levels at the centred time point. Linear slope–slope covariances index coordinated linear change over time, such that individuals showing steeper increases in BAG also show steeper increases in symptoms. Quadratic slope–slope covariances index coordinated nonlinear change across the modelled window, indicating that individuals exhibiting greater acceleration (i.e., curvature) in BAG trajectories across adolescence also exhibit greater acceleration in symptom trajectories over the same period.

All reported results below reflect FDR corrected p values. The full results of the final eight multigroup BLGC models, including all the model parameters, are available in SI Table 8. The results of the parameters of interest are summarised in SI Table 9 and visualised in Figures 3 and 4. For symptom trajectories over time at low versus high BAG and coupling scatter plots between BAG and symptom growth components, see SI Figure 13 and 14.

**Figure 3.**
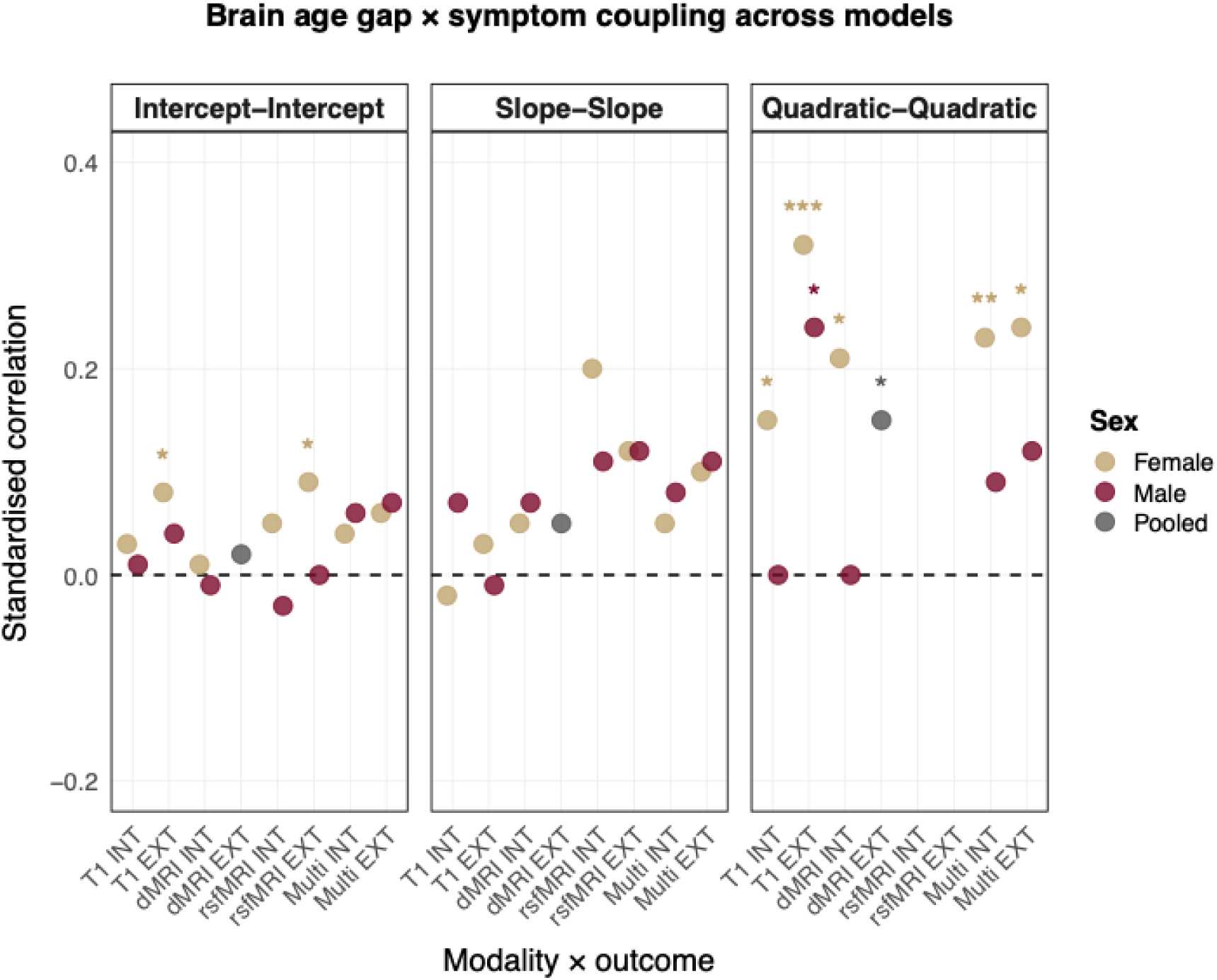
Standardised correlations between BAG and symptom growth factors across imaging modalities and developmental processes. Points represent sex-stratified coupling estimates for all models (female = gold; male = burgundy), except for the dMRI–externalising model, where a single pooled estimate is shown (grey). For T1, dMRI, and multimodal modalities, BAG and symptoms were modelled with centred quadratic growth factors; for rs-fMRI, only linear BAG models converged, so no quadratic coupling is shown. Columns correspond to associations among intercepts (centred levels), linear slopes (rates of change), and quadratic components (curvature). Asterisks indicate statistical significance of the within-sex BAG–symptom covariance *after* FDR correction across all coupling tests (\**p* FDR < .05, \*\**p* FDR < .01, \*\*\**p* FDR < .001).

**Figure 4.**
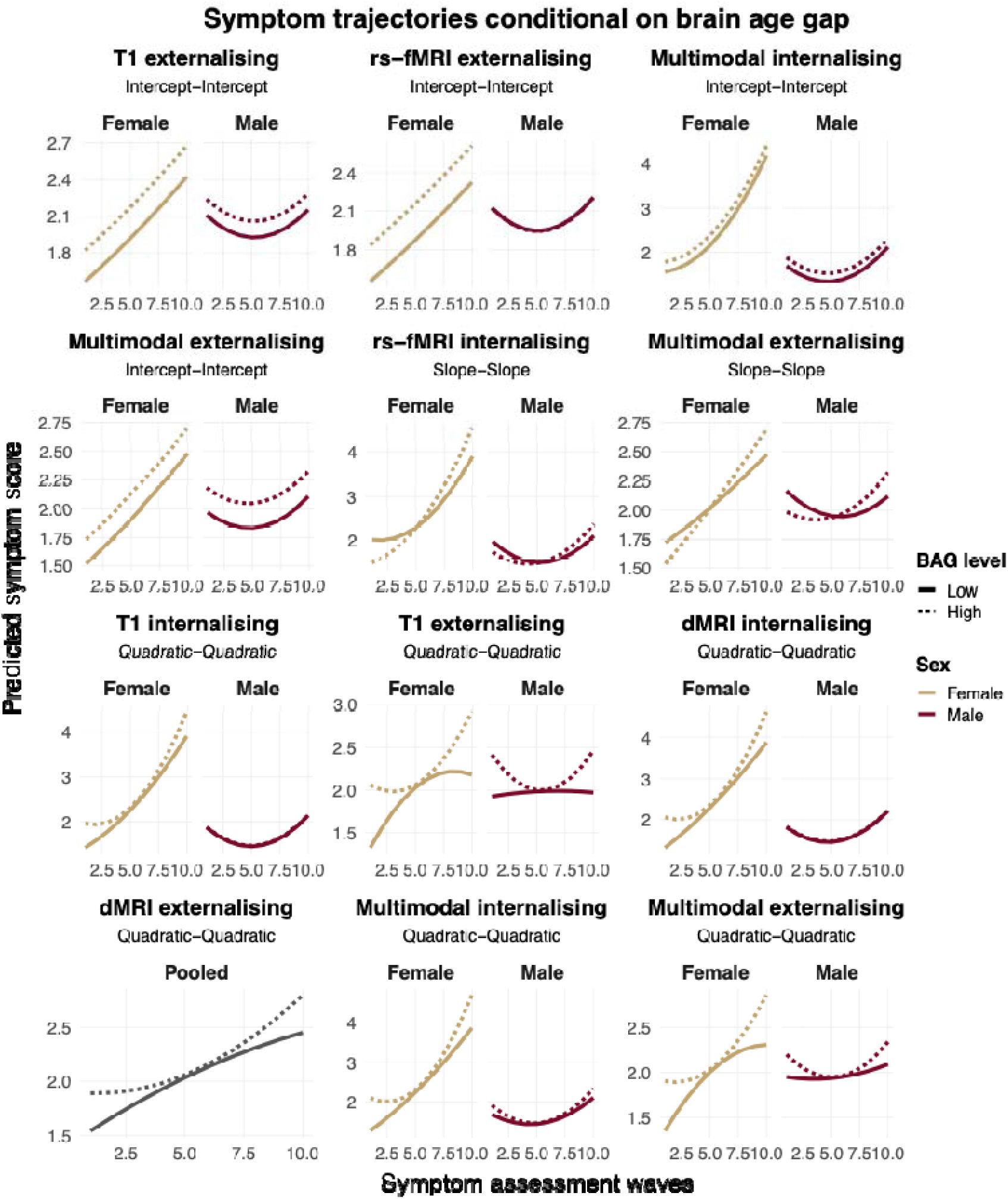
Predicted symptom trajectories conditional on brain age gap. The figure visualises the BLGC coupling effects by plotting predicted internalising and externalising symptom trajectories at low (−1 SD; solid lines) versus high (+1 SD; dotted lines) values of the corresponding BAG growth component. Panels are ordered by modality (T1, dMRI, rs-fMRI, multimodal) with internalising preceding externalising within each modality. Trajectories are shown separately for females (gold) and males (burgundy) where sex-stratified models were estimated; for dMRI externalising, trajectories reflect pooled-sex estimates (grey), consistent with the equality-constrained model selected for that modality. For each modality × outcome coupling combination, the plotted trajectories represent expected symptom development obtained by conditioning the relevant symptom growth factor (intercept, linear slope, or quadratic component) on the corresponding BAG factor, while holding all remaining latent growth factors fixed at their estimated means. Because symptom trajectories are parameterised with quadratic growth functions in the final models, conditional trajectories may appear curved even when visualising intercept–intercept or slope–slope couplings.

#### 3.2.1. Longitudinal coupling between brain age gap and internalising symptoms

All significant BAG–internalising couplings were positive in direction, and associations appeared primarily in the quadratic domain. In the T1 model, the quadratic component of BAG was positively coupled with the quadratic component of internalising in females (*r* = .15, *p* FDR = .044), while the corresponding estimate in males was not statistically significant. The dMRI model showed a similar pattern, with a significant quadratic BAG–internalising coupling in females (*r* = .21, *p* FDR < .044) but not in males. In the multimodal model, the quadratic component of BAG was again significantly coupled with the quadratic component of internalising in females (*r* = .23, *p* FDR < .009), whereas the male estimate was not statistically significant. Together, these results suggest that greater acceleration in BAG trajectories (i.e., increasing deviation from age-expected maturation) tended to co-occur with greater acceleration in internalising symptoms in females over adolescence.

No intercept–intercept or slope–slope associations survived FDR correction in the internalising models. For rs-fMRI, where BAG was modelled linearly, a slope–slope coupling observed in females (r = .20) did not survive FDR correction (*p* FDR = .06). Importantly, although several estimates were significant in one sex but not the other, targeted coupling-specific likelihood-ratio tests did not indicate reliable sex differences in these BAG–internalising couplings (Section 3.2.3; SI Table 10).

#### 3.2.2. Longitudinal coupling between brain age gap and externalising symptoms

Associations between the quadratic components of BAG and externalising symptom trajectories were the most consistent and strongest effects across modalities. In the T1 model, the quadratic component of BAG was coupled with the quadratic component of externalising for both females (r = .32, *p* FDR < .001) and males (r = .24, *p* FDR = .030). In the dMRI model, equality-constrained latent covariances were supported; the BAG–externalising quadratic coupling was therefore estimated as a single pooled effect across sex (r = .15, *p* FDR = .044). In the multimodal model, the quadratic BAG–externalising coupling was significant in females (r = .24, *p* FDR = .028) but not in males after FDR correction.

At the level of intercepts, a significant intercept–intercept association was observed in the T1 model in females (r = .08, *p* FDR = .030) but not in males, and in the rs-fMRI model in females (r = .09, *p* FDR = .044) but not in males. Intercept–intercept associations in the multimodal externalising model did not survive FDR correction in either sex (females: *p* FDR = .097; males: *p* FDR = .062). No slope–slope associations survived FDR correction in the externalising models, including the multimodal slope coupling that was nominally significant prior to correction. The rs-fMRI linear model showed no significant BAG–externalising couplings.

Overall, externalising symptoms showed consistent positive quadratic coupling with BAG trajectories across modalities, indicating that faster or increasingly nonlinear deviations from age-expected brain maturation tended to co-occur with faster acceleration in externalising symptoms over adolescence. As with internalising, targeted coupling-specific tests did not provide evidence for robust sex differences in BAG–externalising couplings (Section 3.2.3; SI Table 10).

#### 3.2.3. Sex differences in brain age gap–symptom coupling

We conducted tests to assess whether specific cross-process couplings between brain age and psychopathology differed by sex. For each imaging modality and outcome, we tested equality constraints on individual latent covariances linking corresponding growth factors. At the nominal level, two coupling-specific sex differences were observed, both involving quadratic component associations with internalising symptoms. Specifically, the quadratic brain age–internalising coupling differed by sex in the T1-weighted model (χ² = 4.42, *p* = .036) and in the diffusion MRI model (χ² = 4.41, *p* = .036). However, neither of these effects survived correction for multiple testing (*p* FDR = .285). No other coupling-specific sex differences were observed. Thus, coupling-specific tests provided no robust evidence that the BAG–symptom cross-process couplings differed by sex. The full set of sex-difference tests is reported in SI Table 10.

#### 3.2.4. Sensitivity analyses

To assess the robustness of primary coupling estimates to socioeconomic confounding, a sensitivity analysis was conducted in which a composite measure of socioeconomic status (SES; combining parental and partner income and educational attainment at baseline) was included as a time-invariant covariate regressed onto all latent growth factors in each BLGC model. Coupling estimates from the SES-adjusted models were highly consistent with primary model estimates across all modalities and symptom domains, supporting the robustness of the reported associations (SI Table 11).

## 4. Discussion

Adolescence is a dynamic period of rapid neurobiological change that coincides with the emergence and consolidation of psychopathology^23,28,87,88^, making it a critical window for studying how these processes co-develop. In this longitudinal study spanning late childhood through mid-adolescence, we examined how developmental deviations in brain age align with changes in internalising and externalising symptoms. Building on prior longitudinal work from our group examining multimodal brain age and internalising symptoms across two waves in ABCD Study^58^, the present study extends this framework to four MRI waves, ten symptom assessment waves, both internalising and externalising symptom domains, and bivariate latent growth curve modelling that explicitly characterises coupled developmental trajectories and their growth factor components.

Using growth curve models, we observed that changes in the pace of brain age maturation were systematically linked to changes in symptom trajectories over time. The most consistent associations emerged for nonlinear components, indicating that accelerations in deviation from age-expected brain maturation were associated with accelerating internalising and externalising symptoms during adolescence. This suggests that nonlinear changes in how individuals diverge from normative brain maturation trajectories, rather than static brain age deviation, may play a key role in linking brain maturation to emerging psychopathology. For internalising symptoms, nonlinear BAG–symptom associations were most consistently detected in females across T1-weighted, diffusion, and multimodal models, whereas corresponding male estimates were smaller and not statistically significant. For externalising symptoms, nonlinear associations were evident in T1 models for both sexes, in diffusion MRI as a sex-invariant pooled effect, and in multimodal models for females. Together, these results highlight synchrony in developmental change as a central feature of brain–psychopathology coupling across adolescence.

Beyond nonlinear effects, a subset of models showed associations between brain age deviation and symptom levels at the centred time point of the observation window. These associations were observed for externalising symptoms in T1-weighted and rs-fMRI models for females. Linear slope associations between rates of brain age change and symptom change did not survive FDR correction across any modality–domain combination. In contrast, rs-fMRI–based brain age showed limited evidence of coupling with symptom trajectories. This modality did not support nonlinear brain age factors and yielded relatively few associations overall, consistent with prior reports of relatively lower reliability in functional connectivity–based brain age estimates in youth samples^89,90^. Our findings partially support our hypothesis that developmental changes in BAG would be coupled with changes in internalising and externalising symptom trajectories. Importantly, coupling-specific tests did not provide robust evidence that these cross-process associations differed by sex after correction for multiple comparisons. As such, sex-stratified estimates should not be interpreted as evidence of sex-specific coupling.

Our findings are broadly in line with structural and multimodal MRI studies reporting that more positive brain age gaps – reflecting older-appearing brains – are associated with greater psychopathology in adolescents^54,55,58^. At the same time, they contrast with reports that relatively younger-appearing brains are linked to higher symptom burden^47,49,52^. These discrepancies indicate that BAG–psychopathology relationships in youth may be heterogeneous across modalities, samples, and analytic approaches. However, most prior studies are cross-sectional or rely on only two time points, limiting their ability to disentangle developmental tempo from stable between-person differences. By modelling intercepts, slopes, and curvature, the present study shows that nonlinear changes in deviation from age-expected brain maturation, rather than static age deviation or constant rates of change, were most consistently aligned with increasing symptoms.

Accelerated patterns of brain maturation during adolescence may reflect developmental mismatch processes in which affective and socioemotional systems mature more rapidly than regulatory and cognitive control systems^91,92^. Adolescence is characterised by heightened neuroplasticity and sensitivity to environmental inputs, such that this maturational asynchrony may increase vulnerability to internalising symptoms when emotional reactivity outpaces regulatory capacity^93^. In line with the stress-acceleration and pubertal timing frameworks, faster biological maturation – particularly in females – has been associated with elevated risk for depressive and internalising symptoms, potentially reflecting heightened responsivity to psychosocial demands during sensitive developmental windows^94–97^. Within this framework, the present findings suggest that accelerating deviations from age-expected brain maturation trajectories may capture an intensified developmental tempo that aligns with increasing internalising symptom trajectories, particularly in modalities capturing structural and multimodal maturation, although this is speculative.

The more consistent detectability of acceleration-linked associations in females is directionally compatible with well-documented sex differences in developmental timing and risk for internalising psychopathology. Females typically enter puberty earlier and show earlier developmental inflection points in cortical and white-matter maturation^95,98^, coinciding with the marked rise in internalising symptoms during early–mid adolescence^22^, and broader sex disparities in internalising symptom prevalence and persistence across adolescence^24,99,100^. However, targeted coupling-specific tests did not provide robust evidence of sex moderation after correction for multiple comparisons, indicating that apparent differences in within-sex significance should not be interpreted as strong evidence that coupling strength differs between females and males. One possibility is that differences in precision (e.g., variability, measurement noise, missingness, or power) contribute to more consistent detection of these associations in females in the present data. Future work incorporating pubertal, hormonal, and sex-related measures, alongside adequately powered moderation tests, will be needed to clarify whether and how sex-linked developmental timing and biological or socio-cultural factors shape the coupling between brain maturation tempo and evolving emotional symptoms. Together, these results position nonlinear deviations in brain maturation tempo as a developmentally meaningful correlate of adolescent psychopathology, while providing limited evidence for robust sex-specific coupling.

### 4.1. Strengths and limitations

A major strength of this study is the combination of four waves of multimodal MRI with ten waves of psychopathology assessment in a large, diverse, community-based cohort. This design enabled trajectory-level analyses that have not been possible in prior brain age research in youth, allowing us to examine how brain maturation and mental health symptoms unfold together across development. The use of bivariate latent growth curve models permitted decomposition of brain and symptom development into intercept, slope, and quadratic components, enabling explicit tests of synchrony in development. Our systematic model-selection process is another key strength. By evaluating linear, interval-based, latent-basis, and centred-quadratic specifications, we ensured that our models balanced empirical fit with parameter stability. This allowed us to identify modalities where nonlinear BAG could be reliably estimated (T1, dMRI, multimodal) and to simplify the rs-fMRI BAG model where quadratic terms were not supported.

Some limitations should also be acknowledged. First, at the time of analysis, release 6.0 of the ABCD Study only had partial data available from the fourth imaging time point, limiting the sample size of participants that had four available scans. Second, rs-fMRI BAG showed low temporal reliability, potentially preventing estimation of curvature and precluding conclusions about functional maturational dynamics. Third, BAG is a statistical index of multivariate deviation rather than a direct biological marker, and its interpretation should remain cautious. Relatedly, the aggregate nature of the BAG index precludes attribution of findings to specific neural networks, regions, or maturational processes, and mechanistic interpretations at the circuit or network level remain an important direction for future work. Regarding the brain age prediction pipeline, although the primary train/test split was fully family-aware, family structure was not preserved within the internal cross-validation folds used for hyperparameter tuning or the 80/20 bias-correction split; while the hold-out test set remained family-clean throughout, this represents a minor limitation that may have introduced slight optimism in hyperparameter selection. Fourth, although the BPM provided dense longitudinal symptom data, it is a brief self-report instrument with only a one-week recall window and does not capture finer-grained or multi-informant clinical variation. Fifth, although FIML allowed us to retain participants with incomplete MRI or symptom data, patterns of missingness and selective attrition may still have introduced bias, highlighting the need for future work incorporating explicit missing-data diagnostics or sensitivity analyses.

Sixth, because the cohort is drawn from a large U.S. sample, generalisability to other cultural or socio-environmental contexts may be constrained. Seventh, although pubertal timing was not included as a covariate in the present analyses, we acknowledge this as a meaningful limitation given the centrality of puberty to adolescent brain maturation and the emergence of internalising psychopathology. The inclusion of sex as a grouping variable provides partial accounting for systematic sex differences in pubertal tempo, and the SES sensitivity analysis offers reassurance that findings are not attributable to a major sociodemographic confound. Nonetheless, pubertal timing, hormonal change, and their interactions with brain maturational trajectories represent important unmeasured factors, and future work incorporating longitudinal pubertal assessments within a developmental coupling framework would be a valuable and necessary extension of the present findings. Finally, the BLGC models capture coordinated change but cannot determine directionality or causal pathways, underscoring the need for future causal designs. The observed effect sizes were small to moderate, and while this is consistent with typical brain–behaviour associations in large community-based neuroimaging studies, the modest magnitudes mean that findings should be interpreted cautiously and require replication in clinically ascertained samples before conclusions about clinical relevance can be drawn.

### 4.2. Conclusion

Adolescence is marked by rapid and uneven change across neurobiological, emotional, and behavioural domains, and the present findings indicate that it is the *tempo* of brain maturation that most meaningfully aligns with internalising and externalising symptoms. We show that accelerations in divergence from normative brain maturation track with parallel accelerations in psychopathology across adolescence. The results support a broader developmental view in which vulnerability arises not from absolute levels of neurobiological features, but from deviations from canonical developmental profiles – patterns in which an individual’s brain maturational trajectory diverges from age-expected norms in tandem with worsening symptom trajectories, offering a framework for understanding why adolescence is a sensitive period for emerging mental health problems.

## Data Availability

Data used in the present study were drawn from the ABCD Study release 6.0, containing data from baseline up until the six-year visit (nbdc-datahub.org/abcd-release-6-0). All ABCD Study data is stored in the NIH Brain Development Cohorts (NBDC) Data Hub, which is available for authorised users with approved Data Use Certification (DUC) (lead investigator: Westlye). The 6.0 release has been assigned the DOI 10.82525/jy7n-g441

## 5. Acknowledgements

Data used in the preparation of this article were obtained from the <u>Adolescent Brain Cognitive Development™ (ABCD) Study</u>, held in the <u>NIH Brain Development Cohorts Data Sharing Platform</u>. This is a multisite, longitudinal study designed to recruit more than 10,000 children aged ∼8–10 and follow them over 10 years into early adulthood. The ABCD Study® is supported by the National Institutes of Health and additional federal partners under award numbers: U01DA041048, U01DA050989, U01DA051016, U01DA041022, U01DA051018, U01DA051037, U01DA050987, U01DA041174, U01DA041106, U01DA041117, U01DA041028, U01DA041134, U01DA050988, U01DA051039, U01DA041156, U01DA041025, U01DA041120, U01DA051038, U01DA041148, U01DA041093, U01DA041089, U24DA041123, U24DA041147. A full list of supporters is available at <u>Federal Partners - ABCD Study</u>. ABCD Consortium investigators designed and implemented the study and/or provided data but did not necessarily participate in the analysis or writing of this report. This manuscript reflects the views of the authors and may not reflect the opinions or views of the NIH or ABCD Consortium investigators.

Data was handled inside Service for Sensitive Data (TSD), a platform owned by the University of Oslo, operated, and developed by the TSD service group at the University of Oslo IT-Department (USIT). Computations were also performed using resources provided by UNINETT Sigma2 – the National Infrastructure for High Performance Computing and Data Storage in Norway (NS9666S).

## 8. CRediT author contribution statement

**Beck, Dani**: Writing – Original draft, review & editing, Project administration, Visualisation, Methodology, Formal analysis, Data curation, Conceptualisation. **Carrick, Chloe:** Writing – review & editing, Formal analysis, Conceptualisation. **Aksnes, Eira R.:** Writing – review & editing, Conceptualisation. **MacSweeney, Niamh.:** Writing – review & editing. **Westlye, Lars T.:** Writing – review & editing, Funding acquisition. **Tamnes, Christian K.:** Writing – review & editing, Project administration, Funding acquisition, Conceptualisation.

