## Supplementary Material for "Synchrony between brain age and internalising and externalising symptoms across adolescence"

| **SI Table 1**. *Test* sample descriptive statistics for each modality of brain-age prediction imaging per sex and timepoint. | | | | |
| --- | --- | --- | --- | --- |
|  | T1 | dMRI | Rs-fMRI | Multi-modal |
| N Males | 7907 (53.3%) | 6853 (52.8%) | 6931 (53.4%) | 6139 (52.2%) |
| N Females | 6931 (46.7%) | 6122 (47.2%) | 6059 (46.6%) | 5631 (47.8%) |
| N wave 1 | 5672 (38.2%) | 5112 (39.4%) | 4685 (36.1%) | 4472 (38.0%) |
| N wave 2 | 3993 (26.9%) | 3420 (26.4%) | 3459 (26.6%) | 3070 (26.1%) |
| N wave 3 | 3132 (21.1%) | 2721 (21.0%) | 2962 (22.8%) | 2596 (22.1%) |
| N wave 4 | 2041 (13.8%) | 1722 (13.3%) | 1884 (14.5%) | 1632 (13.9%) |
| Age; total | 12.24 (SD = 2.29, range = 9.0-17.7) | 12.19 (SD = 2.29, range = 8.3-17.7) | 12.35 (SD = 2.30, range = 9.0-17.7) | 12.28 (SD = 2.30, range = 9.0-17.7) |
| Age; wave 1 | 9.96 (SD = 0.62, range = 9.0-11.0) | 9.96 (SD = 0.63, range = 8.3-11.1) | 9.98 (SD = 0.63, range = 9.0-11.1) | 9.97 (SD = 0.63, range = 8.9-11.1) |
| Age; wave 2 | 12.00 (SD = 0.65, range = 10.7-13.9) | 12.00 (SD = 0.65, range = 10.2-13.8) | 12.00 (SD = 0.65, range = 10.6-13.9) | 12.02 (SD = 0.65, range = 10.2-13.8) |
| Age; wave 3 | 14.17 (SD = 0.72, range = 12.5-16.6) | 14.16 (SD = 0.72, range = 12.5-16.0) | 14.15 (SD = 0.71, range = 12.6-16.1) | 14.16 (SD = 0.72, range = 12.5-15.9) |
| Age; wave 4 | 16.08 (SD = 0.65, range = 14.6-17.7) | 16.10 (SD = 0.65, range = 14.6-17.7) | 16.09 (SD = 0.67, range = 14.7-17.7) | 16.12 (SD = 0.65, range = 14.6-17.7) |

**Note**: N = observations in the dataset.

| **SI Table 2**. *Training* sample descriptive statistics for each modality of brain-age prediction imaging per sex and timepoint. | | | | |
| --- | --- | --- | --- | --- |
|  | T1 | dMRI | Rs-fMRI | Multi-modal |
| N Males | 7698 (52.2%) | 6663 (51.4%) | 6610 (50.6%) | 5931 50.5% |
| N Females | 7058 (47.8%) | 6311 (48.6%) | 6461 (49.4%) | 5807 49.5% |
| N wave 1 | 5652 (38.3%) | 5125 (39.5%) | 4709 (36.0%) | 4493 38.3% |
| N wave 2 | 3904 (26.5%) | 3418 (26.3%) | 3481 (26.6%) | 3045 25.9% |
| N wave 3 | 3164 (21.4%) | 2738 (21.1%) | 2934 (22.4%) | 2585 22.0% |
| N wave 4 | 2036 (13.8%) | 1693 (13.0%) | 1947 (14.9%) | 1615 13.8% |
| Age; total | 12.25 (SD = 2.30, range = 8.3-17.7) | 12.21 (SD = 2.29, range = 8.9-17.7) | 12.38 (SD = 2.31, range = 9.0-17.7) | 12.29 (SD = 2.30, range = 9.0-17.7) |
| Age; wave 1 | 9.96 (SD = 0.63, range = 8.3-11.3) | 9.97 (SD = 0.62, range = 8.9-11.3) | 9.98 (SD = 0.63, range = 9.0-11.3) | 9.99 (SD = 0.62, range = 9.0-11.3) |
| Age; wave 2 | 12.00 (SD = 0.65, range = 10.2-13.8) | 12.02 (SD = 0.65, range = 10.7-13.9) | 12.02 (SD = 0.66, range = 10.23-13.85) | 12.03 (SD = 0.66, range = 10.6-13.9) |
| Age; wave 3 | 14.16 (SD = 0.73, range = 12.5-16.1) | 14.19 (SD = 0.72, range = 12.6-16.6) | 14.19 (SD = 0.73, range = 12.5-16.1) | 14.20 (SD = 0.73, range = 12.5-16.1) |
| Age; wave 4 | 16.11 (SD = 0.66, range = 14.7-17.7) | 16.13 (SD = 0.66, range = 14.7-17.7) | 16.09 (SD = 0.65, range = 14.6-17.7) | 16.11 (SD = 0.66, range = 14.7-17.7) |

**SI Section 1**. MRI acquisition and processing

For T1-weighted MRI acquisitions on Siemens and Philips 3T scanners, a matrix of 256 × 256, 176 slices (Siemens) and 225 slices (Philips) with a field of view (FOV) of 256 × 256, echo time (TE)/repetition time (TR) (ms) of 2.88/2500 (Siemens) and 2.9/6.31 (Philips), flip angle of 8°. On GE scanners, the same matrix and FOV were used with TE/TR (ms) of 2/2500 and a flip angle of 8°. The spatial resolution was consistent at 1.0 × 1.0 × 1.0 mm across all three platforms. The dMRI acquisition (1.7 mm isotropic) uses multiband EPI with slice acceleration factor 3 and includes 96 diffusion directions, seven b = 0 frames, and four b-values (6 directions with b = 500 s/mm^2^, 15 directions with b = 1000 s/mm^2^, 15 directions with b = 2000 s/mm^2^, and 60 directions with b = 3000 s/mm^2^). For rs-fMRI acquisition, the following scanning parameters were used: matrix of 90 × 90, 60 slices, FOV = 216 × 216, TE/TR = 800/30, flip angle = 52° and resolution = 2.4 × 2.4 × 2.4 mm. The fMRI acquisitions (2.4 mm isotropic, TR = 800 ms) used multiband EPI with slice acceleration factor 6 and were split into 2-4 5-minute scanning sessions, in which participants were instructed to keep their eyes open and fixate on a crosshair.

For T1-weighted MRI data, cortical surface reconstruction and subcortical segmentation was performed with FreeSurfer v7.1.1^1,2^. This processing includes motion correction and averaging^3^, removal of non-brain tissue (Ségonne et al., 2004), automated Talairach transformation, segmentation of the subcortical white matter and deep grey matter volumetric structures^2,4^, intensity normalisation^5^, tessellation of the grey matter white matter boundary, automated topology correction^6,7^, and surface deformation^1^. From the ABCD Data Repository, we obtained tabulated data including total and regional measures of cortical surface area, thickness, volume, sulcal depth, intensity-grey-white contrast, and subcortical volume (397 measures).

For DTI data, processing was carried out using AtlasTrack, a probabilistic atlas-based method for automated segmentation of white matter fibre tracts^8^. A detailed pipeline can be found in Hagler et al. (2019). Briefly, diffusion tensor parameters are calculated using a standard, linear estimation approach with log-transformed diffusion-weighted (DW) signals^10^, whereby two tensor models are fitted. In the first DTI model fit (DTI inner shell, or DTI_IS_), frames with b > 1000 s/mm^2^ are excluded from tensor fitting (leaving 6 directions at b = 500 s/mm^2^ and 15 directions at b = 1000 s/mm^2^) so that the derived diffusivity measures better correspond to those from traditional, single-b-value acquisitions. In the second DTI model fit (DTI full shell, or DTI_FS_), all gradient strengths/shells (6 directions at b = 500 s/mm^2^, 15 directions at b = 1000 s/mm^2^, 15 directions at b = 2000 s/mm^2^, and 60 directions at b = 3000 s/mm^2^) are included. For the current study, both full and inner shell tissue properties of functional anisotropy (FA) and mean (MD), longitudinal (or axial, AD), and transverse (or radial, RD) diffusivity were extracted for total and regional features (576 measures).

For rs-fMRI, preprocessing steps are outlined in detail in Hagler et al. (2019). Briefly, head motion is corrected by registering each frame to the first using AFNI’s 3dvolreg^11^, while B_0_ distortions are corrected using the same reversing polarity method used for the dMRI^12^. Preprocessing also includes steps to avoid signal drop-out, corrections for distortions due to gradient nonlinearities^13^ and between-scan motion correction. Additional rs-fMRI processing steps included removal of initial frames, normalisation, regression, temporal filtering, and calculation of ROI-average time courses. Measures of functional connectivity were computed using a seed-based, correlational approach^14^, where average time courses were calculated for cortical surface-based ROIs using a functionally-defined parcellation based on resting-state functional connectivity patterns^15^ and for subcortical ROIs^2^.

Correlation coefficients between the average time courses of each ROI-pairing were then calculated. Here, correlations between unique pairs of ROIs are obtained and Fisher transformed into z-statistics. Connectivity measures represent the averaged Fisher-transformed correlations of all the unique pairs of ROIs either within a cortical network, between cortical networks, or between cortical networks and subcortical regions. ROIs were grouped into Gordon networks and within and between network correlation strength was extracted by calculating the mean correlation coefficient of the respective ROI-pairings. Together with the correlations between the average time course within a network and each subcortical ROI, these coefficients served as functional connectivity measures for the current study. From the ABCD Data Repository, we obtained functional connectivity within and between parcellations from the Gordon network, including subcortical data (416 measures).

For MRI data, quality check procedures followed the standard protocol described in Hagler et al. (2019)^9^. Briefly, participants with excessive head motion or poor data quality were excluded from the curated data release by the ABCD team. Additional quality assurance was carried out following extraction of data using the recommendations for data cleaning provided by the ABCD team (using data structure abcd imgincl01). Here, recommendation criteria for include/exclude is marked by 1/0, respectively, and removal of data marked 0 was carried out. Following this procedure, T1-weighted data was reduced from 30,276 observations (obs) from 4 time points (TP1: 11,755; TP2: 8,092; TP3: 6,343; TP4: 4,086) to 29,594 (TP1: 11,324; TP2: 7,897; TP3: 6,296; TP4: 4,077). DTI data was reduced from 29,165 obs (TP1: 11,130; TP2: 7,842; TP3: 6,224; TP4: 3,969) to 25,949 obs (TP1: 10,237; TP2: 6,838; TP3: 5,459; TP4: 3,415), and rs-fMRI data from 29,526 obs (TP1: 11,269; TP2: 7,924; TP3: 6,274; TP4: 4,059) to 26,061 obs (TP1: 9,394; TP2: 6,940; TP3: 5,896; TP4: 3,831). The dataset for the multi-modal brain age model was formed by merging the aforementioned data sets. The sample of each modality was then split into brain age training and test sets, the demographics of which are documented in SI Tables 1 and 2.

**SI Section 2**. LongComBat

The ABCD Study data is collected from 21 different sites in the U.S, and children are scanned using thirty-four different magnetic resonance imaging (MRI) scanners from three manufacturing brands (3-T Siemens Prisma, General Electric 750, or Phillips). Due to the technical variability of using multiple scanners, noise and bias can be introduced into the estimation of biological features of interest. Longitudinal ComBat is a powerful harmonisation procedure for longitudinal datasets and controls for type I error better than unharmonized data with scanner included as a covariate^16^. In the current study, LongCombat was implemented on baseline, two-, four-, and six-year follow-up observations (obs) of T1 (obs = 29,594), DTI (obs = 25,949), and rs-fMRI (obs = 26,061) data from the ABCD Study cohort, with each MRI modality being harmonised separately, including covariates of age, sex, and timepoint following recommendations from Beer et al. (2020). SI Figures 1-3 shows MRI data from each modality for selected regions prior to and post harmonisation. Note that the number of batches (34) exceeds the number of recruitment sites (21) because some sites operated more than one scanner model or underwent scanner upgrades during data collection. LongComBat was therefore applied at the scanner level, as this represents the primary unit of technical variability in multi-site MRI data.

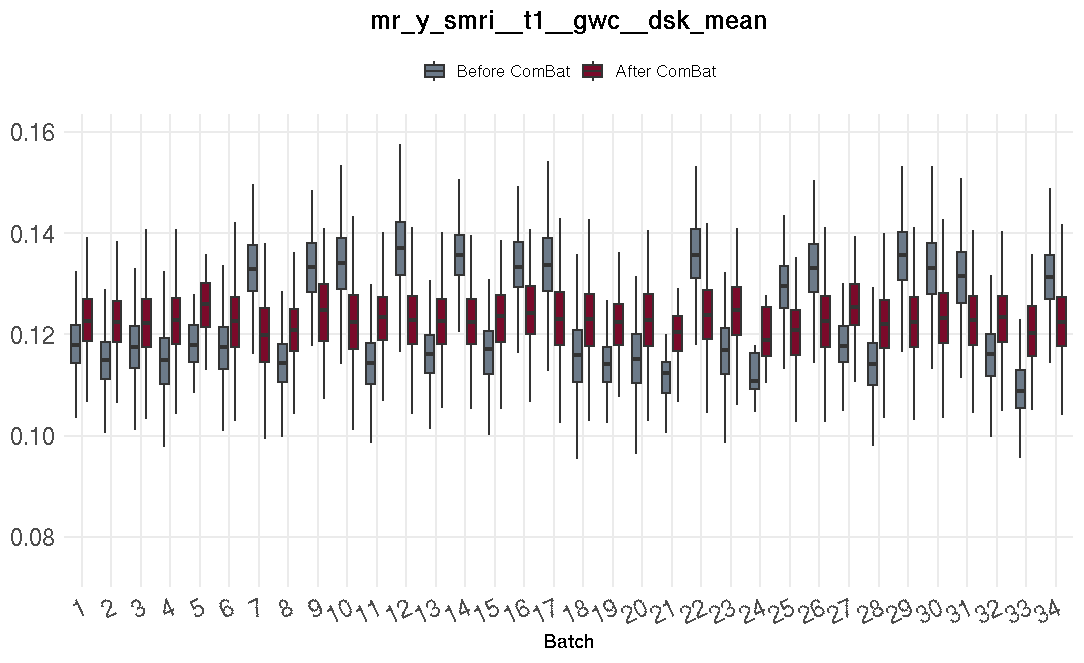

**SI Figure 1**. Showing before and after longComBat harmonisation for a selected measure (regional gray/white contrast) from the t1-weighted dataset. Imaging data from each site (batch) shown in blue before harmonisation and in red after harmonisation.

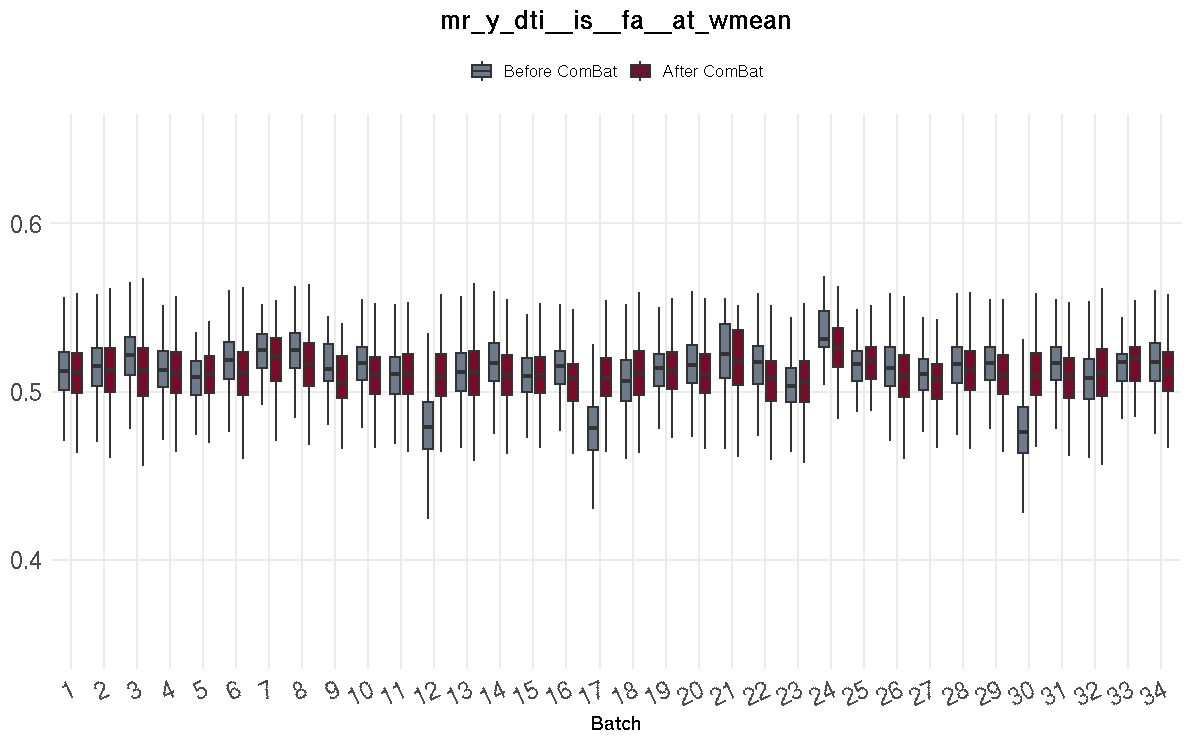

**SI Figure 2**. Showing before and after longComBat harmonisation for a selected measure (fractional anisotropy) from the diffusion-MRI dataset. Imaging data from each site (batch) shown in blue before harmonisation and in red after harmonisation.

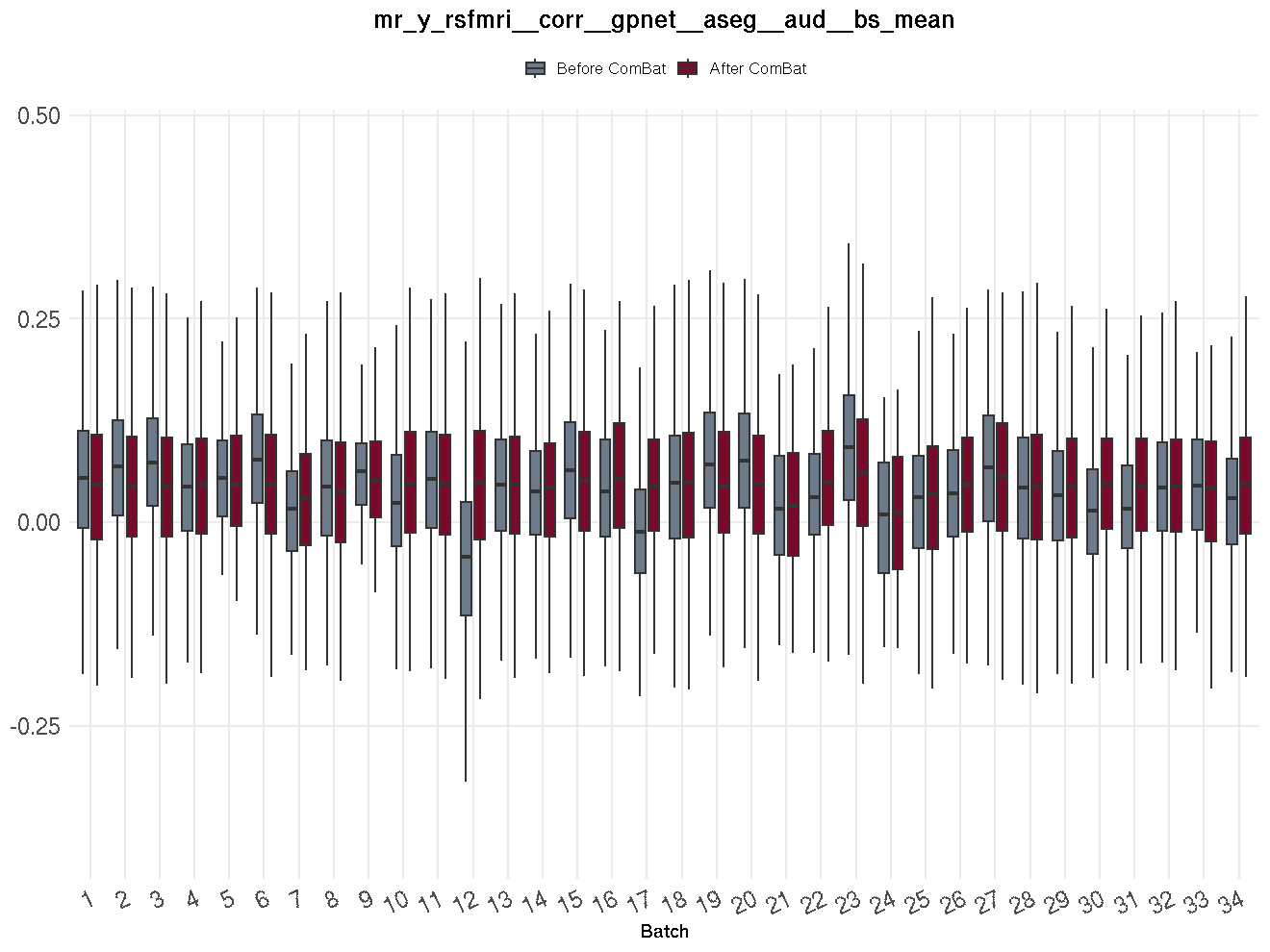

**SI Figure 3**. Showing before and after longComBat harmonisation for a selected measure (brain stem) from the rs-fMRI dataset. Imaging data from each site (batch) shown in blue before harmonisation and in red after harmonisation.

**SI Section 3.** Brain age prediction training and test sample overview

Our imaging data selection was informed by prior brain age studies^17–23^. We chose validated metrics from previous studies: fractional anisotropy, diffusivity measures for DTI; cortical surface area, thickness, subcortical volume for T1; functional connectivity for rs-fMRI. This yielded 416 features for rs-fMRI and 576 for DTI. To match feature counts and reduce bias, we added intensity-grey-white contrast and sulcal depth to the T1 model, increasing its features to 397. We additionally trained a multi-modal model, which includes all the features from the aforementioned models (1389 features in total). For each brain modality (T1, DTI, rs-fMRI, multi-modal), 50% of the data was used as the hold-out test sample and 50% was used for the model training and validation, split 80% and 20% respectively. The sample sizes of each modality are provided in SI Table 1 (test) and SI Table 2 (training). Performance metrics for the test sample are provided in the main manuscript (Table 1). To assess model generalisation, performance metrics were compared between the internal validation set (20% of the training sample) and the hold-out test set. Validation and test set performance were near-identical across all four modalities: T1 (validation: MAE = 1.00, RMSE = 1.27, R² = 0.70; test: MAE = 1.00, RMSE = 1.28, R² = 0.69), dMRI (validation: MAE = 0.92, RMSE = 1.17, R² = 0.74; test: MAE = 0.90, RMSE = 1.15, R² = 0.75), rs-fMRI (validation: MAE = 1.36, RMSE = 1.69, R² = 0.47; test: MAE = 1.34, RMSE = 1.67, R² = 0.47), and multimodal (validation: MAE = 0.86, RMSE = 1.08, R² = 0.78; test: MAE = 0.85, RMSE = 1.09, R² = 0.78). This indicates that hyperparameter selection was not overly optimistic and that the models generalise well to unseen data. Prior to bias correction, raw BAG values showed the expected negative correlation with chronological age across all modalities (r = −0.53 to −0.74), confirming the presence of age bias that was subsequently corrected using the validation set regression procedure described in the Methods. Training set metrics are not reported as they are uninformative for gradient boosting algorithms, which by design fit training data very closely regardless of generalisation performance.

**SI Table 3**. T1-weighted brain-age model features.

thk dsk bstmps lh mean, thk dsk cac lh mean, thk dsk cmfrt lh mean, thk dsk cn lh mean, thk dsk er lh mean, thk dsk ff lh mean, thk dsk ic lh mean, thk dsk ins lh mean, thk dsk iprt lh mean, thk dsk itmp lh mean, thk dsk lg lh mean, thk dsk lobfrt lh mean, thk dsk locc lh mean, thk dsk mobfrt lh mean, thk dsk mtmp lh mean, thk dsk pactr lh mean, thk dsk pcc lh mean, thk dsk pcg lh mean, thk dsk pfrt lh mean, thk dsk ph lh mean, thk dsk pob lh mean, thk dsk poctr lh mean, thk dsk pop lh mean, thk dsk prcn lh mean, thk dsk prctr lh mean, thk dsk ptg lh mean, thk dsk ptmp lh mean, thk dsk rac lh mean, thk dsk rmfrt lh mean, thk dsk sfrt lh mean, thk dsk sm lh mean, thk dsk sprt lh mean, thk dsk stmp lh mean, thk dsk ttmp lh mean, thk dsk bstmps rh mean, thk dsk cac rh mean, thk dsk cmfrt rh mean, thk dsk cn rh mean, thk dsk er rh mean, thk dsk ff rh mean, thk dsk ic rh mean, thk dsk ins rh mean, thk dsk iprt rh mean, thk dsk itmp rh mean, thk dsk lg rh mean, thk dsk lobfrt rh mean, thk dsk locc rh mean, thk dsk mobfrt rh mean, thk dsk mtmp rh mean, thk dsk pactr rh mean, thk dsk pcc rh mean, thk dsk pcg rh mean, thk dsk pfrt rh mean, thk dsk ph rh mean, thk dsk pob rh mean, thk dsk poctr rh mean, thk dsk pop rh mean, thk dsk prcn rh mean, thk dsk prctr rh mean, thk dsk ptg rh mean, thk dsk ptmp rh mean, thk dsk rac rh mean, thk dsk rmfrt rh mean, thk dsk sfrt rh mean, thk dsk sm rh mean, thk dsk sprt rh mean, thk dsk stmp rh mean, thk dsk ttmp rh mean, thk dsk lh mean, thk dsk rh mean, thk dsk mean, area dsk bstmps lh sum, area dsk cac lh sum, area dsk cmfrt lh sum, area dsk cn lh sum, area dsk er lh sum, area dsk ff lh sum, area dsk ic lh sum, area dsk ins lh sum, area dsk iprt lh sum, area dsk itmp lh sum, area dsk lg lh sum, area dsk lobfrt lh sum, area dsk locc lh sum, area dsk mobfrt lh sum, area dsk mtmp lh sum, area dsk pactr lh sum, area dsk pcc lh sum, area dsk pcg lh sum, area dsk pfrt lh sum, area dsk ph lh sum, area dsk pob lh sum, area dsk poctr lh sum, area dsk pop lh sum, area dsk prcn lh sum, area dsk prctr lh sum, area dsk ptg lh sum, area dsk ptmp lh sum, area dsk rac lh sum, area dsk rmfrt lh sum, area dsk sfrt lh sum, area dsk sm lh sum, area dsk sprt lh sum, area dsk stmp lh sum, area dsk ttmp lh sum, area dsk bstmps rh sum, area dsk cac rh sum, area dsk cmfrt rh sum, area dsk cn rh sum, area dsk er rh sum, area dsk ff rh sum, area dsk ic rh sum, area dsk ins rh sum, area dsk iprt rh sum, area dsk itmp rh sum, area dsk lg rh sum, area dsk lobfrt rh sum, area dsk locc rh sum, area dsk mobfrt rh sum, area dsk mtmp rh sum, area dsk pactr rh sum, area dsk pcc rh sum, area dsk pcg rh sum, area dsk pfrt rh sum, area dsk ph rh sum, area dsk pob rh sum, area dsk poctr rh sum, area dsk pop rh sum, area dsk prcn rh sum, area dsk prctr rh sum, area dsk ptg rh sum, area dsk ptmp rh sum, area dsk rac rh sum, area dsk rmfrt rh sum, area dsk sfrt rh sum, area dsk sm rh sum, area dsk sprt rh sum, area dsk stmp rh sum, area dsk ttmp rh sum, area dsk lh sum, area dsk rh sum, area dsk sum, sulc dsk bstmps lh mean, sulc dsk cac lh mean, sulc dsk cmfrt lh mean, sulc dsk cn lh mean, sulc dsk er lh mean, sulc dsk ff lh mean, sulc dsk ic lh mean, sulc dsk ins lh mean, sulc dsk iprt lh mean, sulc dsk itmp lh mean, sulc dsk lg lh mean, sulc dsk lobfrt lh mean, sulc dsk locc lh mean, sulc dsk mobfrt lh mean, sulc dsk mtmp lh mean, sulc dsk pactr lh mean, sulc dsk pcc lh mean, sulc dsk pcg lh mean, sulc dsk pfrt lh mean, sulc dsk ph lh mean, sulc dsk pob lh mean, sulc dsk poctr lh mean, sulc dsk pop lh mean, sulc dsk prcn lh mean, sulc dsk prctr lh mean, sulc dsk ptg lh mean, sulc dsk ptmp lh mean, sulc dsk rac lh mean, sulc dsk rmfrt lh mean, sulc dsk sfrt lh mean, sulc dsk sm lh mean, sulc dsk sprt lh mean, sulc dsk stmp lh mean, sulc dsk ttmp lh mean, sulc dsk bstmps rh mean, sulc dsk cac rh mean, sulc dsk cmfrt rh mean, sulc dsk cn rh mean, sulc dsk er rh mean, sulc dsk ff rh mean, sulc dsk ic rh mean, sulc dsk ins rh mean, sulc dsk iprt rh mean, sulc dsk itmp rh mean, sulc dsk lg rh mean, sulc dsk lobfrt rh mean, sulc dsk locc rh mean, sulc dsk mobfrt rh mean, sulc dsk mtmp rh mean, sulc dsk pactr rh mean, sulc dsk pcc rh mean, sulc dsk pcg rh mean, sulc dsk pfrt rh mean, sulc dsk ph rh mean, sulc dsk pob rh mean, sulc dsk poctr rh mean, sulc dsk pop rh mean, sulc dsk prcn rh mean, sulc dsk prctr rh mean, sulc dsk ptg rh mean, sulc dsk ptmp rh mean, sulc dsk rac rh mean, sulc dsk rmfrt rh mean, sulc dsk sfrt rh mean, sulc dsk sm rh mean, sulc dsk sprt rh mean, sulc dsk stmp rh mean, sulc dsk ttmp rh mean, sulc dsk lh mean, sulc dsk rh mean, sulc dsk mean, vol dsk bstmps lh sum, vol dsk cac lh sum, vol dsk cmfrt lh sum, vol dsk cn lh sum, vol dsk er lh sum, vol dsk ff lh sum, vol dsk ic lh sum, vol dsk ins lh sum, vol dsk iprt lh sum, vol dsk itmp lh sum, vol dsk lg lh sum, vol dsk lobfrt lh sum, vol dsk locc lh sum, vol dsk mobfrt lh sum, vol dsk mtmp lh sum, vol dsk pactr lh sum, vol dsk pcc lh sum, vol dsk pcg lh sum, vol dsk pfrt lh sum, vol dsk ph lh sum, vol dsk pob lh sum, vol dsk poctr lh sum, vol dsk pop lh sum, vol dsk prcn lh sum, vol dsk prctr lh sum, vol dsk ptg lh sum, vol dsk ptmp lh sum, vol dsk rac lh sum, vol dsk rmfrt lh sum, vol dsk sfrt lh sum, vol dsk sm lh sum, vol dsk sprt lh sum, vol dsk stmp lh sum, vol dsk ttmp lh sum, vol dsk bstmps rh sum, vol dsk cac rh sum, vol dsk cmfrt rh sum, vol dsk cn rh sum, vol dsk er rh sum, vol dsk ff rh sum, vol dsk ic rh sum, vol dsk ins rh sum, vol dsk iprt rh sum, vol dsk itmp rh sum, vol dsk lg rh sum, vol dsk lobfrt rh sum, vol dsk locc rh sum, vol dsk mobfrt rh sum, vol dsk mtmp rh sum, vol dsk pactr rh sum, vol dsk pcc rh sum, vol dsk pcg rh sum, vol dsk pfrt rh sum, vol dsk ph rh sum, vol dsk pob rh sum, vol dsk poctr rh sum, vol dsk pop rh sum, vol dsk prcn rh sum, vol dsk prctr rh sum, vol dsk ptg rh sum, vol dsk ptmp rh sum, vol dsk rac rh sum, vol dsk rmfrt rh sum, vol dsk sfrt rh sum, vol dsk sm rh sum, vol dsk sprt rh sum, vol dsk stmp rh sum, vol dsk ttmp rh sum, vol dsk lh sum, vol dsk rh sum, vol dsk sum.x, vol dsk sum.y, vol aseg ab lh sum, vol aseg ag lh sum, vol aseg cbc lh sum, vol aseg cbwm lh sum, vol aseg cd lh sum, vol aseg cwm lh sum, vol aseg hc lh sum, vol aseg ilv lh sum, vol aseg lv lh sum, vol aseg pl lh sum, vol aseg pt lh sum, vol aseg th lh sum, vol aseg vdc lh sum, vol aseg ab rh sum, vol aseg ag rh sum, vol aseg cbc rh sum, vol aseg cbwm rh sum, vol aseg cd rh sum, vol aseg cwm rh sum, vol aseg hc rh sum, vol aseg ilv rh sum, vol aseg lv rh sum, vol aseg pl rh sum, vol aseg pt rh sum, vol aseg th rh sum, vol aseg vdc rh sum, vol aseg 3rdv sum, vol aseg 4thv sum, vol aseg avs sum, vol aseg bs sum, vol aseg cca sum, vol aseg ccam sum, vol aseg ccc sum, vol aseg ccp sum, vol aseg ccpm sum, vol aseg csf sum, vol aseg icv sum, vol aseg lvs sum, vol aseg scgv sum, vol aseg stv sum, vol aseg whb sum, vol aseg wmh sum, t1 gwc dsk bstmps lh mean, t1 gwc dsk cac lh mean, t1 gwc dsk cmfrt lh mean, t1 gwc dsk cn lh mean, t1 gwc dsk er lh mean, t1 gwc dsk ff lh mean, t1 gwc dsk ic lh mean, t1 gwc dsk ins lh mean, t1 gwc dsk iprt lh mean, t1 gwc dsk itmp lh mean, t1 gwc dsk lg lh mean, t1 gwc dsk lobfrt lh mean, t1 gwc dsk locc lh mean, t1 gwc dsk mobfrt lh mean, t1 gwc dsk mtmp lh mean, t1 gwc dsk pactr lh mean, t1 gwc dsk pcc lh mean, t1 gwc dsk pcg lh mean, t1 gwc dsk pfrt lh mean, t1 gwc dsk ph lh mean, t1 gwc dsk pob lh mean, t1 gwc dsk poctr lh mean, t1 gwc dsk pop lh mean, t1 gwc dsk prcn lh mean, t1 gwc dsk prctr lh mean, t1 gwc dsk ptg lh mean, t1 gwc dsk ptmp lh mean, t1 gwc dsk rac lh mean, t1 gwc dsk rmfrt lh mean, t1 gwc dsk sfrt lh mean, t1 gwc dsk sm lh mean, t1 gwc dsk sprt lh mean, t1 gwc dsk stmp lh mean, t1 gwc dsk ttmp lh mean, t1 gwc dsk bstmps rh mean, t1 gwc dsk cac rh mean, t1 gwc dsk cmfrt rh mean, t1 gwc dsk cn rh mean, t1 gwc dsk er rh mean, t1 gwc dsk ff rh mean, t1 gwc dsk ic rh mean, t1 gwc dsk ins rh mean, t1 gwc dsk iprt rh mean, t1 gwc dsk itmp rh mean, t1 gwc dsk lg rh mean, t1 gwc dsk lobfrt rh mean, t1 gwc dsk locc rh mean, t1 gwc dsk mobfrt rh mean, t1 gwc dsk mtmp rh mean, t1 gwc dsk pactr rh mean, t1 gwc dsk pcc rh mean, t1 gwc dsk pcg rh mean, t1 gwc dsk pfrt rh mean, t1 gwc dsk ph rh mean, t1 gwc dsk pob rh mean, t1 gwc dsk poctr rh mean, t1 gwc dsk pop rh mean, t1 gwc dsk prcn rh mean, t1 gwc dsk prctr rh mean, t1 gwc dsk ptg rh mean, t1 gwc dsk ptmp rh mean, t1 gwc dsk rac rh mean, t1 gwc dsk rmfrt rh mean, t1 gwc dsk sfrt rh mean, t1 gwc dsk sm rh mean, t1 gwc dsk sprt rh mean, t1 gwc dsk stmp rh mean, t1 gwc dsk ttmp rh mean, t1 gwc dsk lh mean, t1 gwc dsk rh mean, t1 gwc dsk mean

**SI Table 4**. Diffusion MRI brain-age model features.

fa at atr lh wmean, fa at cgc lh wmean, fa at cgh lh wmean, fa at cst lh wmean, fa at fscs lh wmean, fa at fx lh wmean, fa at fxcut lh wmean, fa at ifo lh wmean, fa at ifsfc lh wmean, fa at ilf lh wmean, fa at pscs lh wmean, fa at pslf lh wmean, fa at scs lh wmean, fa at sifc lh wmean, fa at slf lh wmean, fa at tslf lh wmean, fa at unc lh wmean, fa at atr rh wmean, fa at cgc rh wmean, fa at cgh rh wmean, fa at cst rh wmean, fa at fscs rh wmean, fa at fx rh wmean, fa at fxcut rh wmean, fa at ifo rh wmean, fa at ifsfc rh wmean, fa at ilf rh wmean, fa at pscs rh wmean, fa at pslf rh wmean, fa at scs rh wmean, fa at sifc rh wmean, fa at slf rh wmean, fa at tslf rh wmean, fa at unc rh wmean, fa at cc wmean, fa at fmaj wmean, fa at fmin wmean, fa at lh wmean, fa at rh wmean, fa at nocc lh wmean, fa at nocc rh wmean, fa at wmean, fa aseg ab lh mean, fa aseg ag lh mean, fa aseg cbc lh mean, fa aseg cbwm lh mean, fa aseg cd lh mean, fa aseg cwm lh mean, fa aseg hc lh mean, fa aseg ilv lh mean, fa aseg lv lh mean, fa aseg pl lh mean, fa aseg pt lh mean, fa aseg th lh mean, fa aseg vdc lh mean, fa aseg ab rh mean, fa aseg ag rh mean, fa aseg cbc rh mean, fa aseg cbwm rh mean, fa aseg cd rh mean, fa aseg cwm rh mean, fa aseg hc rh mean, fa aseg ilv rh mean, fa aseg lv rh mean, fa aseg pl rh mean, fa aseg pt rh mean, fa aseg th rh mean, fa aseg vdc rh mean, fa aseg 3rdv mean, fa aseg 4thv mean, fa aseg bs mean, fa aseg csf mean, fa at atr lh wmean, fa at cgc lh wmean, fa at cgh lh wmean, fa at cst lh wmean, fa at fscs lh wmean, fa at fx lh wmean, fa at fxcut lh wmean, fa at ifo lh wmean, fa at ifsfc lh wmean, fa at ilf lh wmean, fa at pscs lh wmean, fa at pslf lh wmean, fa at scs lh wmean, fa at sifc lh wmean, fa at slf lh wmean, fa at tslf lh wmean, fa at unc lh wmean, fa at atr rh wmean, fa at cgc rh wmean, fa at cgh rh wmean, fa at cst rh wmean, fa at fscs rh wmean, fa at fx rh wmean, fa at fxcut rh wmean, fa at ifo rh wmean, fa at ifsfc rh wmean, fa at ilf rh wmean, fa at pscs rh wmean, fa at pslf rh wmean, fa at scs rh wmean, fa at sifc rh wmean, fa at slf rh wmean, fa at tslf rh wmean, fa at unc rh wmean, fa at cc wmean, fa at fmaj wmean, fa at fmin wmean, fa at lh wmean, fa at rh wmean, fa at nocc lh wmean, fa at nocc rh wmean, fa at wmean, fa aseg ab lh mean, fa aseg ag lh mean, fa aseg cbc lh mean, fa aseg cbwm lh mean, fa aseg cd lh mean, fa aseg cwm lh mean, fa aseg hc lh mean, fa aseg ilv lh mean, fa aseg lv lh mean, fa aseg pl lh mean, fa aseg pt lh mean, fa aseg th lh mean, fa aseg vdc lh mean, fa aseg ab rh mean, fa aseg ag rh mean, fa aseg cbc rh mean, fa aseg cbwm rh mean, fa aseg cd rh mean, fa aseg cwm rh mean, fa aseg hc rh mean, fa aseg ilv rh mean, fa aseg lv rh mean, fa aseg pl rh mean, fa aseg pt rh mean, fa aseg th rh mean, fa aseg vdc rh mean, fa aseg 3rdv mean, fa aseg 4thv mean, fa aseg bs mean, fa aseg csf mean, md at atr lh wmean, md at cgc lh wmean, md at cgh lh wmean, md at cst lh wmean, md at fscs lh wmean, md at fx lh wmean, md at fxcut lh wmean, md at ifo lh wmean, md at ifsfc lh wmean, md at ilf lh wmean, md at pscs lh wmean, md at pslf lh wmean, md at scs lh wmean, md at sifc lh wmean, md at slf lh wmean, md at tslf lh wmean, md at unc lh wmean, md at atr rh wmean, md at cgc rh wmean, md at cgh rh wmean, md at cst rh wmean, md at fscs rh wmean, md at fx rh wmean, md at fxcut rh wmean, md at ifo rh wmean, md at ifsfc rh wmean, md at ilf rh wmean, md at pscs rh wmean, md at pslf rh wmean, md at scs rh wmean, md at sifc rh wmean, md at slf rh wmean, md at tslf rh wmean, md at unc rh wmean, md at cc wmean, md at fmaj wmean, md at fmin wmean, md at lh wmean, md at rh wmean, md at nocc lh wmean, md at nocc rh wmean, md at wmean, md aseg ab lh mean, md aseg ag lh mean, md aseg cbc lh mean, md aseg cbwm lh mean, md aseg cd lh mean, md aseg cwm lh mean, md aseg hc lh mean, md aseg ilv lh mean, md aseg lv lh mean, md aseg pl lh mean, md aseg pt lh mean, md aseg th lh mean, md aseg vdc lh mean, md aseg ab rh mean, md aseg ag rh mean, md aseg cbc rh mean, md aseg cbwm rh mean, md aseg cd rh mean, md aseg cwm rh mean, md aseg hc rh mean, md aseg ilv rh mean, md aseg lv rh mean, md aseg pl rh mean, md aseg pt rh mean, md aseg th rh mean, md aseg vdc rh mean, md aseg 3rdv mean, md aseg 4thv mean, md aseg bs mean, md aseg csf mean, md at atr lh wmean, md at cgc lh wmean, md at cgh lh wmean, md at cst lh wmean, md at fscs lh wmean, md at fx lh wmean, md at fxcut lh wmean, md at ifo lh wmean, md at ifsfc lh wmean, md at ilf lh wmean, md at pscs lh wmean, md at pslf lh wmean, md at scs lh wmean, md at sifc lh wmean, md at slf lh wmean, md at tslf lh wmean, md at unc lh wmean, md at atr rh wmean, md at cgc rh wmean, md at cgh rh wmean, md at cst rh wmean, md at fscs rh wmean, md at fx rh wmean, md at fxcut rh wmean, md at ifo rh wmean, md at ifsfc rh wmean, md at ilf rh wmean, md at pscs rh wmean, md at pslf rh wmean, md at scs rh wmean, md at sifc rh wmean, md at slf rh wmean, md at tslf rh wmean, md at unc rh wmean, md at cc wmean, md at fmaj wmean, md at fmin wmean, md at lh wmean, md at rh wmean, md at nocc lh wmean, md at nocc rh wmean, md at wmean, md aseg ab lh mean, md aseg ag lh mean, md aseg cbc lh mean, md aseg cbwm lh mean, md aseg cd lh mean, md aseg cwm lh mean, md aseg hc lh mean, md aseg ilv lh mean, md aseg lv lh mean, md aseg pl lh mean, md aseg pt lh mean, md aseg th lh mean, md aseg vdc lh mean, md aseg ab rh mean, md aseg ag rh mean, md aseg cbc rh mean, md aseg cbwm rh mean, md aseg cd rh mean, md aseg cwm rh mean, md aseg hc rh mean, md aseg ilv rh mean, md aseg lv rh mean, md aseg pl rh mean, md aseg pt rh mean, md aseg th rh mean, md aseg vdc rh mean, md aseg 3rdv mean, md aseg 4thv mean, md aseg bs mean, md aseg csf mean, ld at atr lh wmean, ld at cgc lh wmean, ld at cgh lh wmean, ld at cst lh wmean, ld at fscs lh wmean, ld at fx lh wmean, ld at fxcut lh wmean, ld at ifo lh wmean, ld at ifsfc lh wmean, ld at ilf lh wmean, ld at pscs lh wmean, ld at pslf lh wmean, ld at scs lh wmean, ld at sifc lh wmean, ld at slf lh wmean, ld at tslf lh wmean, ld at unc lh wmean, ld at atr rh wmean, ld at cgc rh wmean, ld at cgh rh wmean, ld at cst rh wmean, ld at fscs rh wmean, ld at fx rh wmean, ld at fxcut rh wmean, ld at ifo rh wmean, ld at ifsfc rh wmean, ld at ilf rh wmean, ld at pscs rh wmean, ld at pslf rh wmean, ld at scs rh wmean, ld at sifc rh wmean, ld at slf rh wmean, ld at tslf rh wmean, ld at unc rh wmean, ld at cc wmean, ld at fmaj wmean, ld at fmin wmean, ld at lh wmean, ld at rh wmean, ld at nocc lh wmean, ld at nocc rh wmean, ld at wmean, ld aseg ab lh mean, ld aseg ag lh mean, ld aseg cbc lh mean, ld aseg cbwm lh mean, ld aseg cd lh mean, ld aseg cwm lh mean, ld aseg hc lh mean, ld aseg ilv lh mean, ld aseg lv lh mean, ld aseg pl lh mean, ld aseg pt lh mean, ld aseg th lh mean, ld aseg vdc lh mean, ld aseg ab rh mean, ld aseg ag rh mean, ld aseg cbc rh mean, ld aseg cbwm rh mean, ld aseg cd rh mean, ld aseg cwm rh mean, ld aseg hc rh mean, ld aseg ilv rh mean, ld aseg lv rh mean, ld aseg pl rh mean, ld aseg pt rh mean, ld aseg th rh mean, ld aseg vdc rh mean, ld aseg 3rdv mean, ld aseg 4thv mean, ld aseg bs mean, ld aseg csf mean, ld at atr lh wmean, ld at cgc lh wmean, ld at cgh lh wmean, ld at cst lh wmean, ld at fscs lh wmean, ld at fx lh wmean, ld at fxcut lh wmean, ld at ifo lh wmean, ld at ifsfc lh wmean, ld at ilf lh wmean, ld at pscs lh wmean, ld at pslf lh wmean, ld at scs lh wmean, ld at sifc lh wmean, ld at slf lh wmean, ld at tslf lh wmean, ld at unc lh wmean, ld at atr rh wmean, ld at cgc rh wmean, ld at cgh rh wmean, ld at cst rh wmean, ld at fscs rh wmean, ld at fx rh wmean, ld at fxcut rh wmean, ld at ifo rh wmean, ld at ifsfc rh wmean, ld at ilf rh wmean, ld at pscs rh wmean, ld at pslf rh wmean, ld at scs rh wmean, ld at sifc rh wmean, ld at slf rh wmean, ld at tslf rh wmean, ld at unc rh wmean, ld at cc wmean, ld at fmaj wmean, ld at fmin wmean, ld at lh wmean, ld at rh wmean, ld at nocc lh wmean, ld at nocc rh wmean, ld at wmean, ld aseg ab lh mean, ld aseg ag lh mean, ld aseg cbc lh mean, ld aseg cbwm lh mean, ld aseg cd lh mean, ld aseg cwm lh mean, ld aseg hc lh mean, ld aseg ilv lh mean, ld aseg lv lh mean, ld aseg pl lh mean, ld aseg pt lh mean, ld aseg th lh mean, ld aseg vdc lh mean, ld aseg ab rh mean, ld aseg ag rh mean, ld aseg cbc rh mean, ld aseg cbwm rh mean, ld aseg cd rh mean, ld aseg cwm rh mean, ld aseg hc rh mean, ld aseg ilv rh mean, ld aseg lv rh mean, ld aseg pl rh mean, ld aseg pt rh mean, ld aseg th rh mean, ld aseg vdc rh mean, ld aseg 3rdv mean, ld aseg 4thv mean, ld aseg bs mean, ld aseg csf mean, td at atr lh wmean, td at cgc lh wmean, td at cgh lh wmean, td at cst lh wmean, td at fscs lh wmean, td at fx lh wmean, td at fxcut lh wmean, td at ifo lh wmean, td at ifsfc lh wmean, td at ilf lh wmean, td at pscs lh wmean, td at pslf lh wmean, td at scs lh wmean, td at sifc lh wmean, td at slf lh wmean, td at tslf lh wmean, td at unc lh wmean, td at atr rh wmean, td at cgc rh wmean, td at cgh rh wmean, td at cst rh wmean, td at fscs rh wmean, td at fx rh wmean, td at fxcut rh wmean, td at ifo rh wmean, td at ifsfc rh wmean, td at ilf rh wmean, td at pscs rh wmean, td at pslf rh wmean, td at scs rh wmean, td at sifc rh wmean, td at slf rh wmean, td at tslf rh wmean, td at unc rh wmean, td at cc wmean, td at fmaj wmean, td at fmin wmean, td at lh wmean, td at rh wmean, td at nocc lh wmean, td at nocc rh wmean, td at wmean, td aseg ab lh mean, td aseg ag lh mean, td aseg cbc lh mean, td aseg cbwm lh mean, td aseg cd lh mean, td aseg cwm lh mean, td aseg hc lh mean, td aseg ilv lh mean, td aseg lv lh mean, td aseg pl lh mean, td aseg pt lh mean, td aseg th lh mean, td aseg vdc lh mean, td aseg ab rh mean, td aseg ag rh mean, td aseg cbc rh mean, td aseg cbwm rh mean, td aseg cd rh mean, td aseg cwm rh mean, td aseg hc rh mean, td aseg ilv rh mean, td aseg lv rh mean, td aseg pl rh mean, td aseg pt rh mean, td aseg th rh mean, td aseg vdc rh mean, td aseg 3rdv mean, td aseg 4thv mean, td aseg bs mean, td aseg csf mean, td at atr lh wmean, td at cgc lh wmean, td at cgh lh wmean, td at cst lh wmean, td at fscs lh wmean, td at fx lh wmean, td at fxcut lh wmean, td at ifo lh wmean, td at ifsfc lh wmean, td at ilf lh wmean, td at pscs lh wmean, td at pslf lh wmean, td at scs lh wmean, td at sifc lh wmean, td at slf lh wmean, td at tslf lh wmean, td at unc lh wmean, td at atr rh wmean, td at cgc rh wmean, td at cgh rh wmean, td at cst rh wmean, td at fscs rh wmean, td at fx rh wmean, td at fxcut rh wmean, td at ifo rh wmean, td at ifsfc rh wmean, td at ilf rh wmean, td at pscs rh wmean, td at pslf rh wmean, td at scs rh wmean, td at sifc rh wmean, td at slf rh wmean, td at tslf rh wmean, td at unc rh wmean, td at cc wmean, td at fmaj wmean, td at fmin wmean, td at lh wmean, td at rh wmean, td at nocc lh wmean, td at nocc rh wmean, td at wmean, td aseg ab lh mean, td aseg ag lh mean, td aseg cbc lh mean, td aseg cbwm lh mean, td aseg cd lh mean, td aseg cwm lh mean, td aseg hc lh mean, td aseg ilv lh mean, td aseg lv lh mean, td aseg pl lh mean, td aseg pt lh mean, td aseg th lh mean, td aseg vdc lh mean, td aseg ab rh mean, td aseg ag rh mean, td aseg cbc rh mean, td aseg cbwm rh mean, td aseg cd rh mean, td aseg cwm rh mean, td aseg hc rh mean, td aseg ilv rh mean, td aseg lv rh mean, td aseg pl rh mean, td aseg pt rh mean, td aseg th rh mean, td aseg vdc rh mean, td aseg 3rdv mean, td aseg 4thv mean, td aseg bs mean, td aseg csf mean

**SI Table 5**. Resting state fMRI brain-age model features.

corr gpnet aud aud mean, corr gpnet aud cio mean, corr gpnet aud cip mean, corr gpnet aud def mean, corr gpnet aud doa mean, corr gpnet aud frp mean, corr gpnet aud non mean, corr gpnet aud ret mean, corr gpnet aud sal mean, corr gpnet aud smh mean, corr gpnet aud smm mean, corr gpnet aud vea mean, corr gpnet aud vis mean, corr gpnet cio aud mean, corr gpnet cio cio mean, corr gpnet cio cip mean, corr gpnet cio def mean, corr gpnet cio doa mean, corr gpnet cio frp mean, corr gpnet cio non mean, corr gpnet cio ret mean, corr gpnet cio sal mean, corr gpnet cio smh mean, corr gpnet cio smm mean, corr gpnet cio vea mean, corr gpnet cio vis mean, corr gpnet cip aud mean, corr gpnet cip cio mean, corr gpnet cip cip mean, corr gpnet cip def mean, corr gpnet cip doa mean, corr gpnet cip frp mean, corr gpnet cip non mean, corr gpnet cip ret mean, corr gpnet cip sal mean, corr gpnet cip smh mean, corr gpnet cip smm mean, corr gpnet cip vea mean, corr gpnet cip vis mean, corr gpnet def aud mean, corr gpnet def cio mean, corr gpnet def cip mean, corr gpnet def def mean, corr gpnet def doa mean, corr gpnet def frp mean, corr gpnet def non mean, corr gpnet def ret mean, corr gpnet def sal mean, corr gpnet def smh mean, corr gpnet def smm mean, corr gpnet def vea mean, corr gpnet def vis mean, corr gpnet doa aud mean, corr gpnet doa cio mean, corr gpnet doa cip mean, corr gpnet doa def mean, corr gpnet doa doa mean, corr gpnet doa frp mean, corr gpnet doa non mean, corr gpnet doa ret mean, corr gpnet doa sal mean, corr gpnet doa smh mean, corr gpnet doa smm mean, corr gpnet doa vea mean, corr gpnet doa vis mean, corr gpnet frp aud mean, corr gpnet frp cio mean, corr gpnet frp cip mean, corr gpnet frp def mean, corr gpnet frp doa mean, corr gpnet frp frp mean, corr gpnet frp non mean, corr gpnet frp ret mean, corr gpnet frp sal mean, corr gpnet frp smh mean, corr gpnet frp smm mean, corr gpnet frp vea mean, corr gpnet frp vis mean, corr gpnet non aud mean, corr gpnet non cio mean, corr gpnet non cip mean, corr gpnet non def mean, corr gpnet non doa mean, corr gpnet non frp mean, corr gpnet non non mean, corr gpnet non ret mean, corr gpnet non sal mean, corr gpnet non smh mean, corr gpnet non smm mean, corr gpnet non vea mean, corr gpnet non vis mean, corr gpnet ret aud mean, corr gpnet ret cio mean, corr gpnet ret cip mean, corr gpnet ret def mean, corr gpnet ret doa mean, corr gpnet ret frp mean, corr gpnet ret non mean, corr gpnet ret ret mean, corr gpnet ret sal mean, corr gpnet ret smh mean, corr gpnet ret smm mean, corr gpnet ret vea mean, corr gpnet ret vis mean, corr gpnet sal aud mean, corr gpnet sal cio mean, corr gpnet sal cip mean, corr gpnet sal def mean, corr gpnet sal doa mean, corr gpnet sal frp mean, corr gpnet sal non mean, corr gpnet sal ret mean, corr gpnet sal sal mean, corr gpnet sal smh mean, corr gpnet sal smm mean, corr gpnet sal vea mean, corr gpnet sal vis mean, corr gpnet smh aud mean, corr gpnet smh cio mean, corr gpnet smh cip mean, corr gpnet smh def mean, corr gpnet smh doa mean, corr gpnet smh frp mean, corr gpnet smh non mean, corr gpnet smh ret mean, corr gpnet smh sal mean, corr gpnet smh smh mean, corr gpnet smh smm mean, corr gpnet smh vea mean, corr gpnet smh vis mean, corr gpnet smm aud mean, corr gpnet smm cio mean, corr gpnet smm cip mean, corr gpnet smm def mean, corr gpnet smm doa mean, corr gpnet smm frp mean, corr gpnet smm non mean, corr gpnet smm ret mean, corr gpnet smm sal mean, corr gpnet smm smh mean, corr gpnet smm smm mean, corr gpnet smm vea mean, corr gpnet smm vis mean, corr gpnet vea aud mean, corr gpnet vea cio mean, corr gpnet vea cip mean, corr gpnet vea def mean, corr gpnet vea doa mean, corr gpnet vea frp mean, corr gpnet vea non mean, corr gpnet vea ret mean, corr gpnet vea sal mean, corr gpnet vea smh mean, corr gpnet vea smm mean, corr gpnet vea vea mean, corr gpnet vea vis mean, corr gpnet vis aud mean, corr gpnet vis cio mean, corr gpnet vis cip mean, corr gpnet vis def mean, corr gpnet vis doa mean, corr gpnet vis frp mean, corr gpnet vis non mean, corr gpnet vis ret mean, corr gpnet vis sal mean, corr gpnet vis smh mean, corr gpnet vis smm mean, corr gpnet vis vea mean, corr gpnet vis vis mean, corr gpnet aseg aud ab lh mean, corr gpnet aseg aud ag lh mean, corr gpnet aseg aud cbc lh mean, corr gpnet aseg aud cd lh mean, corr gpnet aseg aud hc lh mean, corr gpnet aseg aud pl lh mean, corr gpnet aseg aud pt lh mean, corr gpnet aseg aud th lh mean, corr gpnet aseg aud vdc lh mean, corr gpnet aseg cio ab lh mean, corr gpnet aseg cio ag lh mean, corr gpnet aseg cio cbc lh mean, corr gpnet aseg cio cd lh mean, corr gpnet aseg cio hc lh mean, corr gpnet aseg cio pl lh mean, corr gpnet aseg cio pt lh mean, corr gpnet aseg cio th lh mean, corr gpnet aseg cio vdc lh mean, corr gpnet aseg cip ab lh mean, corr gpnet aseg cip ag lh mean, corr gpnet aseg cip cbc lh mean, corr gpnet aseg cip cd lh mean, corr gpnet aseg cip hc lh mean, corr gpnet aseg cip pl lh mean, corr gpnet aseg cip pt lh mean, corr gpnet aseg cip th lh mean, corr gpnet aseg cip vdc lh mean, corr gpnet aseg def ab lh mean, corr gpnet aseg def ag lh mean, corr gpnet aseg def cbc lh mean, corr gpnet aseg def cd lh mean, corr gpnet aseg def hc lh mean, corr gpnet aseg def pl lh mean, corr gpnet aseg def pt lh mean, corr gpnet aseg def th lh mean, corr gpnet aseg def vdc lh mean, corr gpnet aseg doa ab lh mean, corr gpnet aseg doa ag lh mean, corr gpnet aseg doa cbc lh mean, corr gpnet aseg doa cd lh mean, corr gpnet aseg doa hc lh mean, corr gpnet aseg doa pl lh mean, corr gpnet aseg doa pt lh mean, corr gpnet aseg doa th lh mean, corr gpnet aseg doa vdc lh mean, corr gpnet aseg frp ab lh mean, corr gpnet aseg frp ag lh mean, corr gpnet aseg frp cbc lh mean, corr gpnet aseg frp cd lh mean, corr gpnet aseg frp hc lh mean, corr gpnet aseg frp pl lh mean, corr gpnet aseg frp pt lh mean, corr gpnet aseg frp th lh mean, corr gpnet aseg frp vdc lh mean, corr gpnet aseg non ab lh mean, corr gpnet aseg non ag lh mean, corr gpnet aseg non cbc lh mean, corr gpnet aseg non cd lh mean, corr gpnet aseg non hc lh mean, corr gpnet aseg non pl lh mean, corr gpnet aseg non pt lh mean, corr gpnet aseg non th lh mean, corr gpnet aseg non vdc lh mean, corr gpnet aseg ret ab lh mean, corr gpnet aseg ret ag lh mean, corr gpnet aseg ret cbc lh mean, corr gpnet aseg ret cd lh mean, corr gpnet aseg ret hc lh mean, corr gpnet aseg ret pl lh mean, corr gpnet aseg ret pt lh mean, corr gpnet aseg ret th lh mean, corr gpnet aseg ret vdc lh mean, corr gpnet aseg sal ab lh mean, corr gpnet aseg sal ag lh mean, corr gpnet aseg sal cbc lh mean, corr gpnet aseg sal cd lh mean, corr gpnet aseg sal hc lh mean, corr gpnet aseg sal pl lh mean, corr gpnet aseg sal pt lh mean, corr gpnet aseg sal th lh mean, corr gpnet aseg sal vdc lh mean, corr gpnet aseg smh ab lh mean, corr gpnet aseg smh ag lh mean, corr gpnet aseg smh cbc lh mean, corr gpnet aseg smh cd lh mean, corr gpnet aseg smh hc lh mean, corr gpnet aseg smh pl lh mean, corr gpnet aseg smh pt lh mean, corr gpnet aseg smh th lh mean, corr gpnet aseg smh vdc lh mean, corr gpnet aseg smm ab lh mean, corr gpnet aseg smm ag lh mean, corr gpnet aseg smm cbc lh mean, corr gpnet aseg smm cd lh mean, corr gpnet aseg smm hc lh mean, corr gpnet aseg smm pl lh mean, corr gpnet aseg smm pt lh mean, corr gpnet aseg smm th lh mean, corr gpnet aseg smm vdc lh mean, corr gpnet aseg vea ab lh mean, corr gpnet aseg vea ag lh mean, corr gpnet aseg vea cbc lh mean, corr gpnet aseg vea cd lh mean, corr gpnet aseg vea hc lh mean, corr gpnet aseg vea pl lh mean, corr gpnet aseg vea pt lh mean, corr gpnet aseg vea th lh mean, corr gpnet aseg vea vdc lh mean, corr gpnet aseg vis ab lh mean, corr gpnet aseg vis ag lh mean, corr gpnet aseg vis cbc lh mean, corr gpnet aseg vis cd lh mean, corr gpnet aseg vis hc lh mean, corr gpnet aseg vis pl lh mean, corr gpnet aseg vis pt lh mean, corr gpnet aseg vis th lh mean, corr gpnet aseg vis vdc lh mean, corr gpnet aseg aud ab rh mean, corr gpnet aseg aud ag rh mean, corr gpnet aseg aud cbc rh mean, corr gpnet aseg aud cd rh mean, corr gpnet aseg aud hc rh mean, corr gpnet aseg aud pl rh mean, corr gpnet aseg aud pt rh mean, corr gpnet aseg aud th rh mean, corr gpnet aseg aud vdc rh mean, corr gpnet aseg cio ab rh mean, corr gpnet aseg cio ag rh mean, corr gpnet aseg cio cbc rh mean, corr gpnet aseg cio cd rh mean, corr gpnet aseg cio hc rh mean, corr gpnet aseg cio pl rh mean, corr gpnet aseg cio pt rh mean, corr gpnet aseg cio th rh mean, corr gpnet aseg cio vdc rh mean, corr gpnet aseg cip ab rh mean, corr gpnet aseg cip ag rh mean, corr gpnet aseg cip cbc rh mean, corr gpnet aseg cip cd rh mean, corr gpnet aseg cip hc rh mean, corr gpnet aseg cip pl rh mean, corr gpnet aseg cip pt rh mean, corr gpnet aseg cip th rh mean, corr gpnet aseg cip vdc rh mean, corr gpnet aseg def ab rh mean, corr gpnet aseg def ag rh mean, corr gpnet aseg def cbc rh mean, corr gpnet aseg def cd rh mean, corr gpnet aseg def hc rh mean, corr gpnet aseg def pl rh mean, corr gpnet aseg def pt rh mean, corr gpnet aseg def th rh mean, corr gpnet aseg def vdc rh mean, corr gpnet aseg doa ab rh mean, corr gpnet aseg doa ag rh mean, corr gpnet aseg doa cbc rh mean, corr gpnet aseg doa cd rh mean, corr gpnet aseg doa hc rh mean, corr gpnet aseg doa pl rh mean, corr gpnet aseg doa pt rh mean, corr gpnet aseg doa th rh mean, corr gpnet aseg doa vdc rh mean, corr gpnet aseg frp ab rh mean, corr gpnet aseg frp ag rh mean, corr gpnet aseg frp cbc rh mean, corr gpnet aseg frp cd rh mean, corr gpnet aseg frp hc rh mean, corr gpnet aseg frp pl rh mean, corr gpnet aseg frp pt rh mean, corr gpnet aseg frp th rh mean, corr gpnet aseg frp vdc rh mean, corr gpnet aseg non ab rh mean, corr gpnet aseg non ag rh mean, corr gpnet aseg non cbc rh mean, corr gpnet aseg non cd rh mean, corr gpnet aseg non hc rh mean, corr gpnet aseg non pl rh mean, corr gpnet aseg non pt rh mean, corr gpnet aseg non th rh mean, corr gpnet aseg non vdc rh mean, corr gpnet aseg ret ab rh mean, corr gpnet aseg ret ag rh mean, corr gpnet aseg ret cbc rh mean, corr gpnet aseg ret cd rh mean, corr gpnet aseg ret hc rh mean, corr gpnet aseg ret pl rh mean, corr gpnet aseg ret pt rh mean, corr gpnet aseg ret th rh mean, corr gpnet aseg ret vdc rh mean, corr gpnet aseg sal ab rh mean, corr gpnet aseg sal ag rh mean, corr gpnet aseg sal cbc rh mean, corr gpnet aseg sal cd rh mean, corr gpnet aseg sal hc rh mean, corr gpnet aseg sal pl rh mean, corr gpnet aseg sal pt rh mean, corr gpnet aseg sal th rh mean, corr gpnet aseg sal vdc rh mean, corr gpnet aseg smh ab rh mean, corr gpnet aseg smh ag rh mean, corr gpnet aseg smh cbc rh mean, corr gpnet aseg smh cd rh mean, corr gpnet aseg smh hc rh mean, corr gpnet aseg smh pl rh mean, corr gpnet aseg smh pt rh mean, corr gpnet aseg smh th rh mean, corr gpnet aseg smh vdc rh mean, corr gpnet aseg smm ab rh mean, corr gpnet aseg smm ag rh mean, corr gpnet aseg smm cbc rh mean, corr gpnet aseg smm cd rh mean, corr gpnet aseg smm hc rh mean, corr gpnet aseg smm pl rh mean, corr gpnet aseg smm pt rh mean, corr gpnet aseg smm th rh mean, corr gpnet aseg smm vdc rh mean, corr gpnet aseg vea ab rh mean, corr gpnet aseg vea ag rh mean, corr gpnet aseg vea cbc rh mean, corr gpnet aseg vea cd rh mean, corr gpnet aseg vea hc rh mean, corr gpnet aseg vea pl rh mean, corr gpnet aseg vea pt rh mean, corr gpnet aseg vea th rh mean, corr gpnet aseg vea vdc rh mean, corr gpnet aseg vis ab rh mean, corr gpnet aseg vis ag rh mean, corr gpnet aseg vis cbc rh mean, corr gpnet aseg vis cd rh mean, corr gpnet aseg vis hc rh mean, corr gpnet aseg vis pl rh mean, corr gpnet aseg vis pt rh mean, corr gpnet aseg vis th rh mean, corr gpnet aseg vis vdc rh mean, corr gpnet aseg aud bs mean, corr gpnet aseg cio bs mean, corr gpnet aseg cip bs mean, corr gpnet aseg def bs mean, corr gpnet aseg doa bs mean, corr gpnet aseg frp bs mean, corr gpnet aseg non bs mean, corr gpnet aseg ret bs mean, corr gpnet aseg sal bs mean, corr gpnet aseg smh bs mean, corr gpnet aseg smm bs mean, corr gpnet aseg vea bs mean, corr gpnet aseg vis bs mean

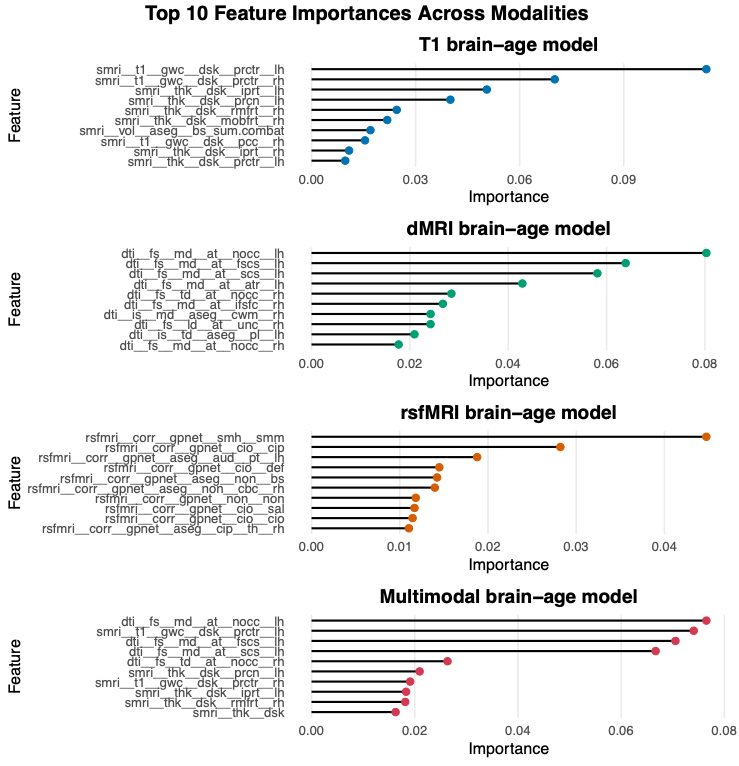

**SI Figure 4**. **Feature importance profiles for brain-age prediction across four imaging modalities.** Variable importance estimates are shown for models trained on T1-weighted structural MRI, diffusion MRI, rs-fMRI, and the combined multi-modal feature set. Each panel displays the relative contribution of individual features to brain-age prediction accuracy within that modality, highlighting modality-specific patterns and shared contributors to the multimodal model. For reference, the top contributing features in each modality were: T1 – grey/white matter contrast and cortical thickness in frontal and precentral regions, and brainstem volume; dMRI – mean diffusivity in non-occipital white matter tracts; rs-fMRI – connectivity between somatomotor and cingulo-opercular networks; multimodal – mean diffusivity in non-occipital tracts and grey/white matter contrast in precentral cortex.

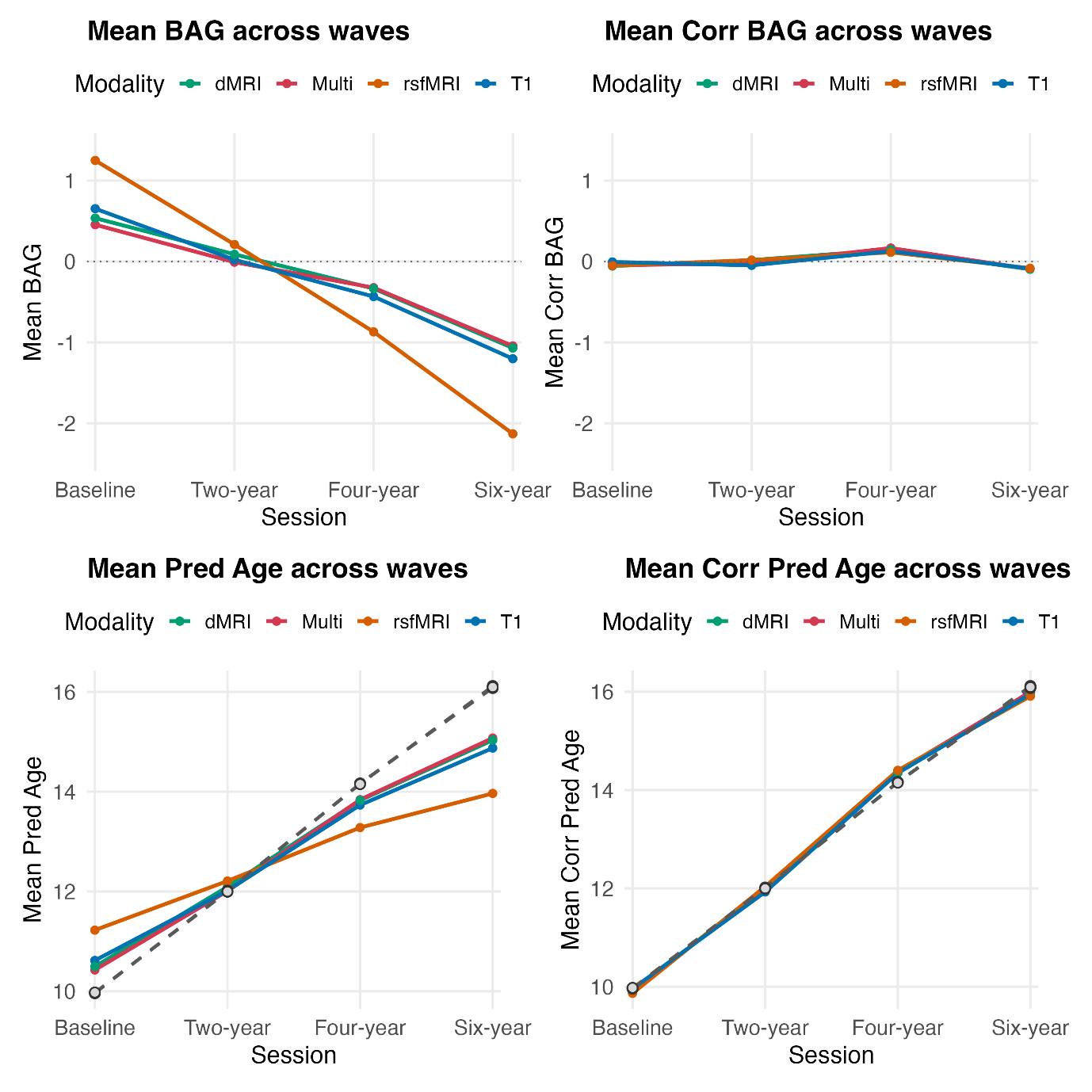

**SI Figure 5**. Four panels show the mean trajectories (± points) across study waves (baseline, two-year, four-year, six-year) for three imaging modalities (T1, dMRI, rs-fMRI) and the multi-modal model. Top row: mean brain-age gap (BAG) and mean corrected brain-age gap (Corr BAG). Bottom row: mean predicted age and mean corrected predicted age (Corr Pred Age). Dashed grey lines in bottom row with circular markers indicate the mean chronological age at each wave.

**SI Section 4**. Brief Problems Monitor quality control

To minimise downstream imputation burden and reduce potential bias from highly incomplete cases, we evaluated missingness at the participant level using item-level variables from the BPM subscales (internalising and externalising). For each participant, we evaluated number of available waves, total number or item-level data, and proportion of missing cells across all item indicators. Participants with ≥ 40% missing data across these item-level measures were flagged for exclusion. This threshold was chosen to ensure sufficient observed information for stable imputation while avoiding excessive model dependence for individuals with very sparse data. Only one participant exceeded this ≥ 40% missingness criterion and was removed from further analyses.

Next, some rows in the BPM dataset contained missing age values (mh_ y_ bpm_ age) at session ses-00M. Because this session occurs approximately 6 months after baseline, missing age values were estimated using available age information from adjacent visits. Two rules were applied to fill missing mh_ y_ bpm_ age at ses-00M: 1) If a participant had a valid baseline age, the missing ses-00M age was set to: baseline age + 0.5 years, 2) If baseline age was unavailable but the participant had a valid age at ses-01A, the missing ses-00M age was set to: ses-01A age − 0.5 years. Only rows where: session id = ses-00M *and* mh_ y_ bpm_ age was missing were updated. Existing, non-missing age values at ses-00M remained unchanged. This step ensured that age was available for all ses-00M observations bar 0.2%.

Item-level missingness in the BPM questionnaire was minimal (See SI Figure 6 below), ranging from approximately 2.8–3.4% across measurement occasions. Because missingness was substantially below commonly recommended thresholds for multiple imputation (e.g., ≥5–10%), and because our longitudinal structural equation models (BLGC) were estimated using full-information maximum likelihood (FIML) – which provides unbiased parameter estimates under missing-at-random assumptions while retaining all partially observed cases – we elected not to perform multiple imputation. Given the extremely low rate of item non-response, additional imputation was unlikely to materially improve estimation precision or reduce bias.

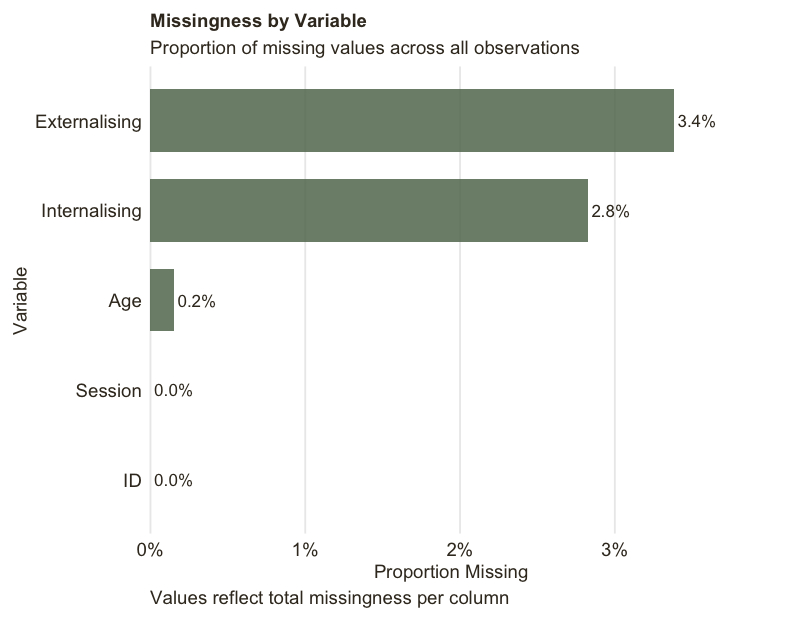

**SI Figure 6**. Proportion of missing data for each variable in the mental-health data set.

**SI Section 5**. Bivariate latent growth curve (BLGC) model selection procedure.

Because brain MRI and symptom data differed in their number of assessments (4 vs. 10 waves) and temporal spacing (biennial MRI, semi-annual → annual BPM), we implemented a structured model-selection procedure to identify an appropriate functional form for the BLGCs. We compared model fit and convergence between models fit with linear, quadratic, and latent basis slopes. We first evaluated several linear specifications. A wave-indexed linear model (BAG: 0–3; BPM: 0–9) yielded suboptimal fit across modalities (CFI ≈ .89–.94, RMSEA ≈ .055–.067, SRMR ≈ .066–.082). A second linear model used interval-based loadings reflecting the approximate chronological spacing (MRI: 0–2–4–6 years; BPM: 0.5–1–1.5 and so on, with annual spacing for the final two waves). Although this factor loading structure better represented the timing of each assessment, model fit and parameter stability did not improve. Finally, we tested a latent-basis model, where we fixed the first factor loading to 0, the last factor loading to 1, and freely estimated the factor loadings of the intervening timepoints. This approach was used to accommodate nonlinear change between timepoints. Across modalities, the latent-basis model frequently produced inadmissible solutions, including negative residual variances (Heywood cases), non–positive-definite covariance matrices, and unstable slope-factor variances, indicating insufficient information for freely estimated nonlinear forms.

We next estimated quadratic models. Conventional, uncentered polynomial loadings (0–3 for BAG; 0–9 for BPM) improved overall fit but produced multicollinearity between linear and quadratic slopes (*r* ≈ .90), occasional boundary variances, and convergence instability – most notably for diffusion and multimodal BAG. Quadratic terms were included to capture potential nonlinear developmental change, consistent with evidence that both brain maturation and psychopathology exhibit acceleration and deceleration across adolescence, rather than constant rates of change. To address estimation instability, we reparameterised time using centred polynomial loadings, which reduce dependency among intercept, linear slope, and curvature terms and improve parameter interpretability. **Although centring does not strictly orthogonalise linear and quadratic growth factors, it substantially attenuates their correlation and mitigates multicollinearity in practice.** For BAG, centred linear loadings were coded −1.5, −0.5, 0.5, 1.5 with corresponding squared values for the quadratic factor; for symptoms, linear loadings were coded −4.5 to 4.5 in unit increments, again with squared loadings for the quadratic factor. This centred quadratic model converged cleanly and yielded stable parameters for T1, diffusion, and multimodal BAG, markedly reduced linear–quadratic correlations (r ≈ .05–.25), and provided the strongest overall fit (CFI ≈ .95–.98, RMSEA ≈ .04–.07, SRMR ≈ .04–.08). In contrast, rs-fMRI BAG remained problematic under both centred quadratic and latent-basis specifications, producing recurrent Heywood cases and non–positive-definite covariance matrices despite acceptable global fit indices. Therefore, for rs-fMRI BAG only, we adopted a simpler linear growth model (intercept and slope), which converged cleanly and produced plausible variance and covariance estimates. Symptom trajectories retained the centred quadratic specification. Quadratic terms were retained only in modalities where they improved model fit and could be estimated reliably, avoiding overfitting where nonlinear effects were unsupported. Thus, the centred quadratic model was selected as the optimal growth specification for T1, dMRI, and multimodal BAG (and for both symptom domains), whereas rs-fMRI BAG was modelled linearly. All subsequent BAG–symptom coupling analyses and sex difference tests are based on these final modality-specific growth models.

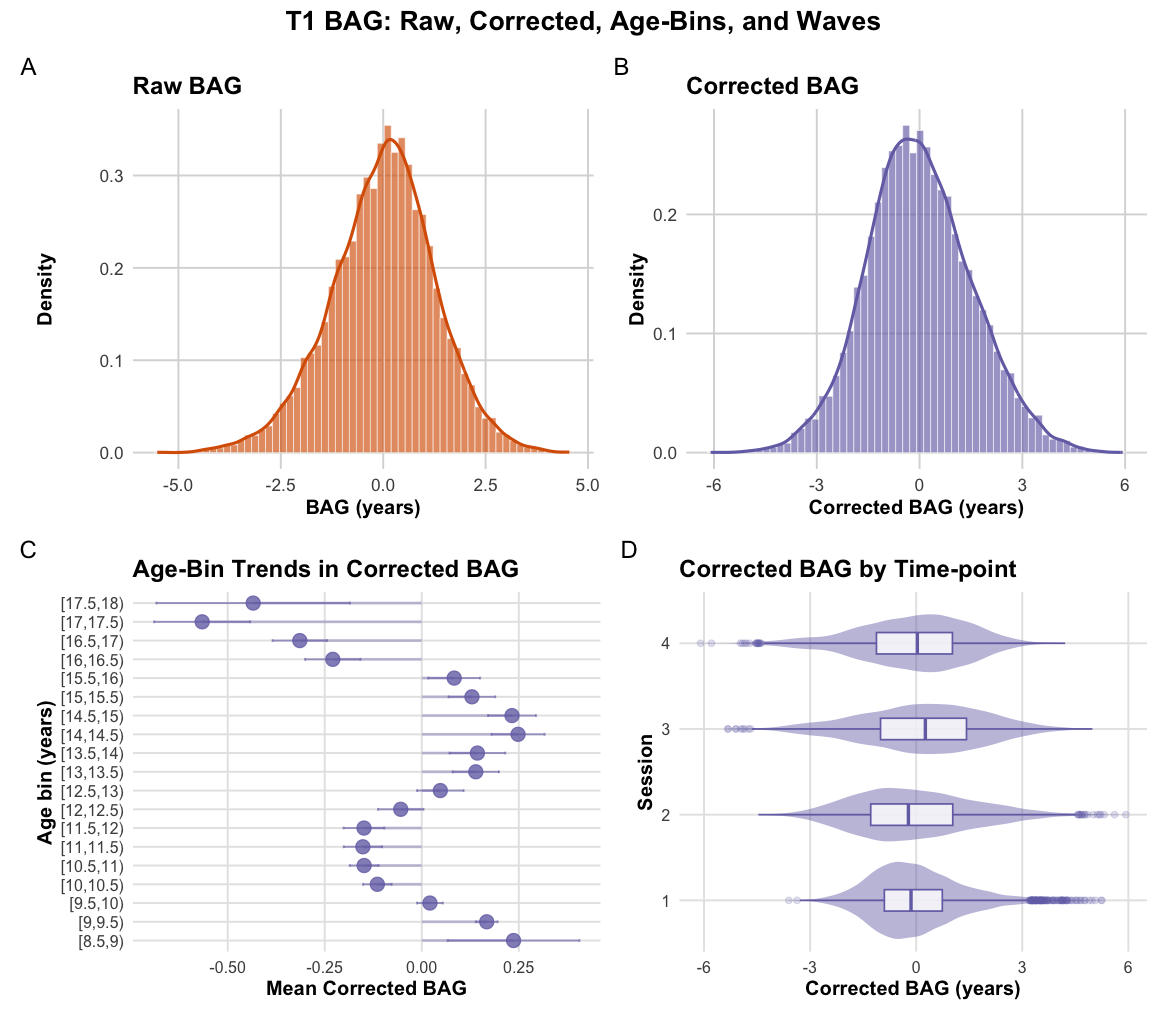

**SI Figure 7. T1-weighted brain–age gap (BAG) distributions and summary patterns.** (A) Distribution of raw BAG. (B) Distribution of bias-corrected BAG, showing a narrower and more symmetric shape relative to the raw estimate. (C) Mean corrected BAG across 0.5-year chronological-age bins. Mean values fluctuate slightly above and below zero depending on the age bin. (D) Corrected BAG stratified by imaging wave, illustrating modest between-wave differences.

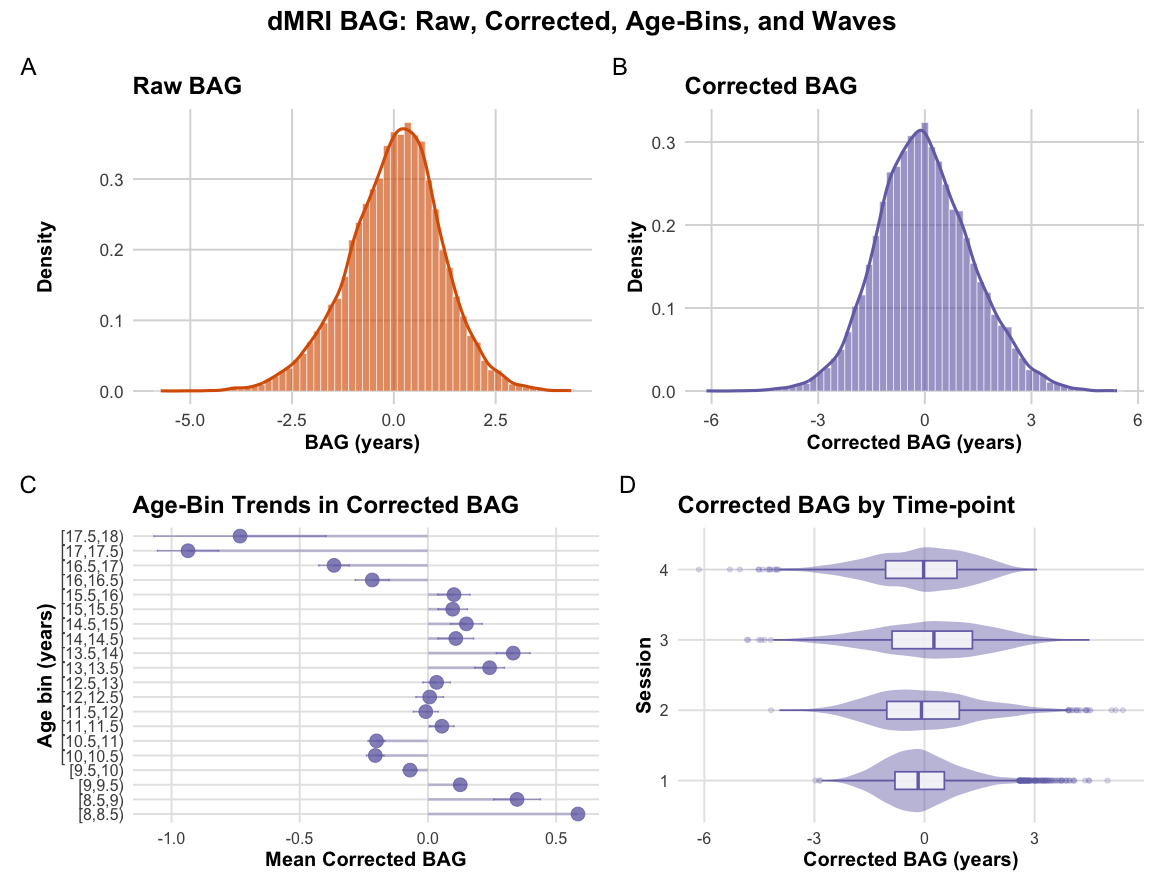

**SI Figure 8.** **Diffusion MRI BAG distributions and summary patterns.** (A) Distribution of raw BAG. (B) Distribution of bias-corrected BAG. (C) Mean corrected BAG across 0.5-year chronological-age bins. (D) Corrected BAG stratified by imaging wave.

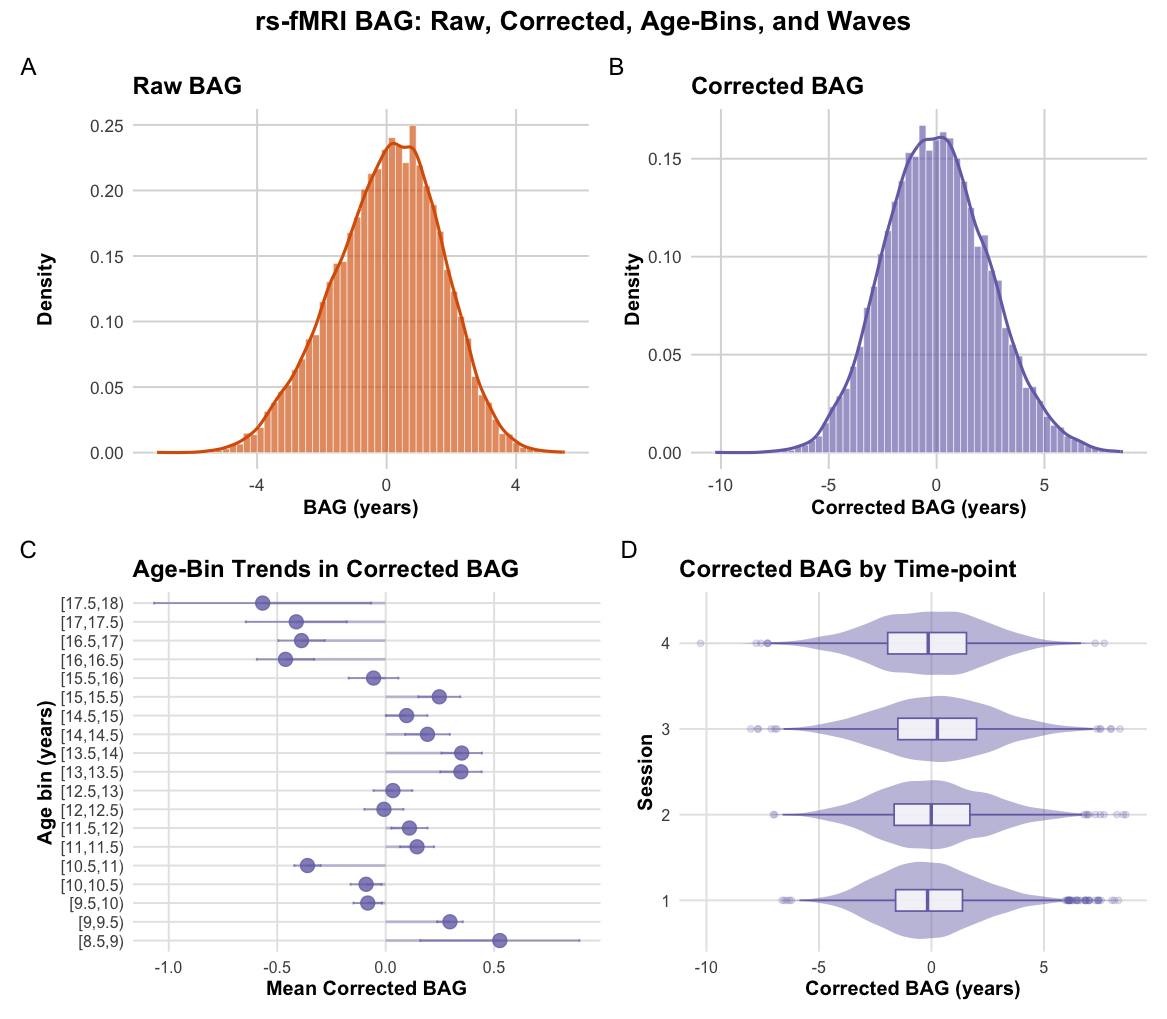

**SI Figure 9.** **Resting state functional MRI BAG distributions and summary patterns.** (A) Distribution of raw BAG. (B) Distribution of bias-corrected BAG. (C) Mean corrected BAG across 0.5-year chronological-age bins. (D) Corrected BAG stratified by imaging wave.

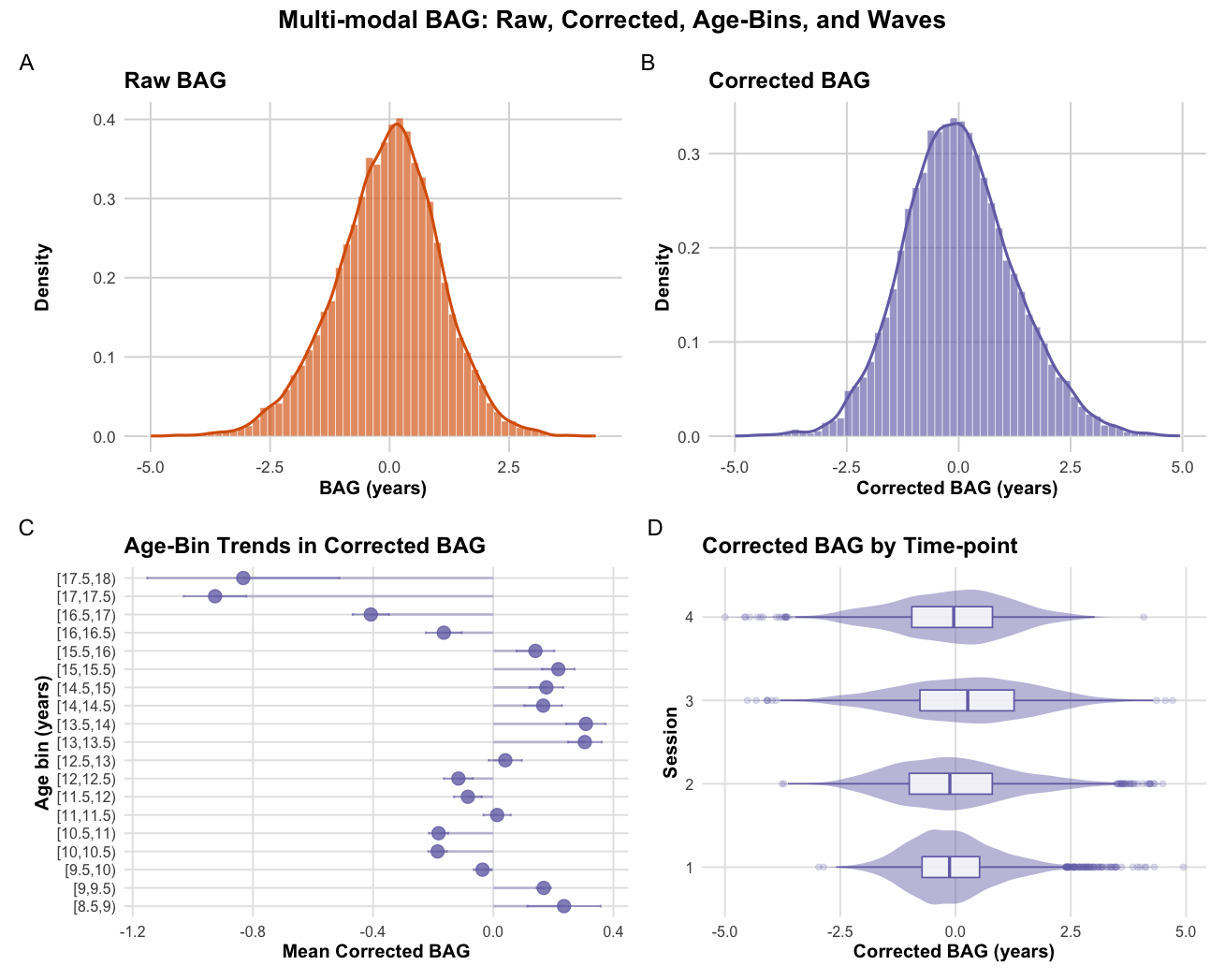

**SI Figure 10. Multi-modal MRI BAG distributions and summary patterns.** (A) Distribution of raw BAG. (B) Distribution of bias-corrected BAG. (C) Mean corrected BAG across 0.5-year chronological-age bins. (D) Corrected BAG stratified by imaging wave.

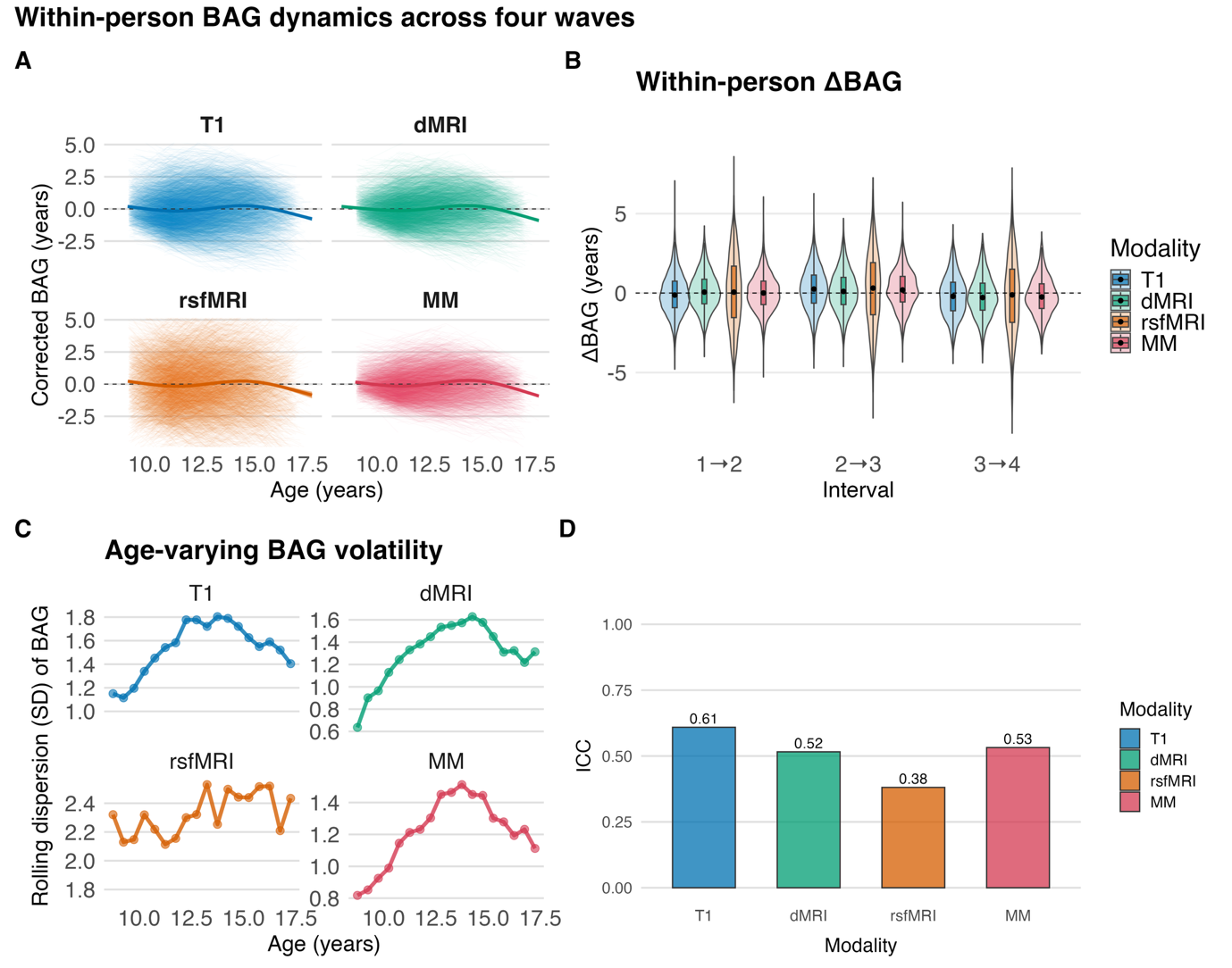

**SI Figure 11. Within-person change in corrected BAG**. (A) Individual corrected BAG trajectories (thin lines) are shown against age for each imaging modality, overlaid with a penalized‐spline GAM and 95% confidence ribbon. Mean BAG remains approximately flat following bias correction, while individual variability is evident with modest modality differences. (B) Distributions of within-person change (ΔBAG) between adjacent sessions (1→2, 2→3, 3→4) reveal small, symmetric fluctuations centred near zero across modalities, indicating limited systematic increases or decreases in BAG during adolescence. (C) Rolling dispersion (SD) of corrected BAG within narrow age bins (0.5 years) highlights age-specific heterogeneity in BAG expression. Variability differs by modality, with T1, dMRI, and multimodal BAG showing more stable variability across age. (D) Longitudinal stability of corrected BAG was quantified using random intercept intraclass correlations (ICCs) within each modality. Stability varied substantially: T1 showed the highest ICC, followed by multimodal and dMRI, whereas rsfMRI demonstrated the lowest stability, consistent with greater state dependence. Together, these panels show that mean BAG is largely stable across adolescence, but individual-level fluctuations and temporal reliability differ slightly across modalities.

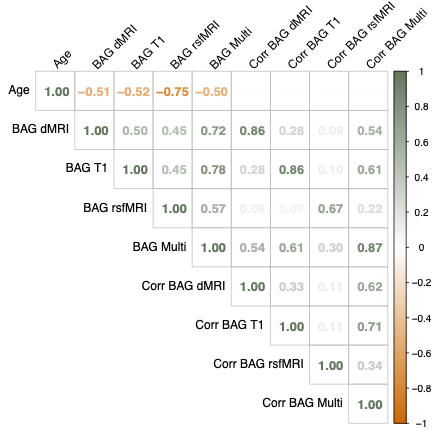

**SI Figure 12**. Correlation matrix showing the associations between age and modality-specific brain-age gap (BAG) for raw and age-bias corrected (Corr) values.

| **SI Table 6.** Model fit indices for the bivariate latent growth curve models across imaging modalities and internalising and externalising domains. | | | | | | |
| --- | --- | --- | --- | --- | --- | --- |
| **Model** | **CFI** | **TLI** | **RMSEA** | **SRMR** | **AIC** | **BIC** |
| T1 internalising | 0.96 | 0.96 | 0.05 | 0.04 | 233342.25 | 233889.76 |
| T1 externalising | 0.97 | 0.97 | 0.04 | 0.04 | 218044.34 | 218591.85 |
| dMRI internalising | 0.96 | 0.96 | 0.04 | 0.05 | 200697.24 | 201233.47 |
| dMRI externalising | 0.98 | 0.97 | 0.04 | 0.04 | 186865.83 | 187303.96 |
| rs-fMRI internalising | 0.95 | 0.95 | 0.04 | 0.04 | 235328.77 | 235779.18 |
| rs-fMRI externalising | 0.97 | 0.97 | 0.04 | 0.04 | 220142.65 | 220593.06 |
| Multimodal internalising | 0.96 | 0.95 | 0.05 | 0.06 | 190127.76 | 190661.08 |
| Multimodal externalising | 0.97 | 0.97 | 0.04 | 0.05 | 176665.44 | 177198.76 |

| **SI Table 7**. **Results of sex-difference tests for latent covariances.** Scaled Satorra–Bentler χ² difference tests comparing freely estimated and equality-constrained multigroup models. Significant results indicate that at least one latent covariance differs by sex. All modalities except the dMRI–externalising model showed evidence of sex-specific covariance structure. | | | | | | | | |
| --- | --- | --- | --- | --- | --- | --- | --- | --- |
| **Modality** | **Outcome** | **df free** | **χ² free** | **χ² equal constraint** | **χ² diff** | **df diff** | ***P* value** | **Sex diff** |
| Multimodal | EXT | 156 | 787.78 | 829.61 | 38.32 | 15 | *p* < .01 | Yes |
| Multimodal | INT | 156 | 1027.90 | 1221.18 | 164.93 | 15 | *p* < .01 | Yes |
| T1 | EXT | 156 | 952.91 | 1005.11 | 48.82 | 15 | *p* < .01 | Yes |
| T1 | INT | 156 | 1151.69 | 1366.05 | 186.29 | 15 | *p* < .01 | Yes |
| dMRI | EXT | 156 | 743.46 | 761.76 | 16.18 | 15 | 0.37 | No |
| dMRI | INT | 156 | 917.19 | 1067.87 | 128.84 | 15 | *p* < .01 | Yes |
| rsfMRI | EXT | 170 | 808.15 | 840.64 | 29.09 | 10 | *p* < .01 | Yes |
| rsfMRI | INT | 170 | 1049.39 | 1216.79 | 135.09 | 10 | *p* < .01 | Yes |

**SI Table 8**. **Full parameter estimates from the final multi-group bivariate latent growth curve (BLGC) models.** Parameter estimates for all eight BLGC models examining associations between brain-age gap (BAG) trajectories and internalising or externalising symptoms across imaging modalities (T1, dMRI, rs-fMRI, multimodal).

| **Model** | **Effect** | **Est** | **SE** | **P value** | **Est std** | **Sex** |
| --- | --- | --- | --- | --- | --- | --- |
| Multi EXT | bag i ~~ bag i | 1.351 | 0.069 | < 0.01 | 1 | Female |
| Multi EXT | bag i ~~ bag i | 1.335 | 0.062 | < 0.01 | 1 | Male |
| Multi INT | bag i ~~ bag i | 1.353 | 0.069 | < 0.01 | 1 | Female |
| Multi INT | bag i ~~ bag i | 1.336 | 0.062 | < 0.01 | 1 | Male |
| T1 EXT | bag i ~~ bag i | 2.322 | 0.093 | < 0.01 | 1 | Female |
| T1 EXT | bag i ~~ bag i | 2.11 | 0.08 | < 0.01 | 1 | Male |
| T1 INT | bag i ~~ bag i | 2.333 | 0.093 | < 0.01 | 1 | Female |
| T1 INT | bag i ~~ bag i | 2.108 | 0.08 | < 0.01 | 1 | Male |
| dMRI EXT | bag i ~~ bag i | 1.614 | 0.062 | < 0.01 | 1 | Female |
| dMRI EXT | bag i ~~ bag i | 1.585 | 0.058 | < 0.01 | 1 | Male |
| dMRI INT | bag i ~~ bag i | 1.597 | 0.077 | < 0.01 | 1 | Female |
| dMRI INT | bag i ~~ bag i | 1.599 | 0.072 | < 0.01 | 1 | Male |
| rsfMRI EXT | bag i ~~ bag i | 1.569 | 0.107 | < 0.01 | 1 | Female |
| rsfMRI EXT | bag i ~~ bag i | 1.85 | 0.112 | < 0.01 | 1 | Male |
| rsfMRI INT | bag i ~~ bag i | 1.573 | 0.107 | < 0.01 | 1 | Female |
| rsfMRI INT | bag i ~~ bag i | 1.851 | 0.112 | < 0.01 | 1 | Male |
| Multi EXT | bag i ~~ bag q | -0.179 | 0.024 | < 0.01 | -0.591 | Female |
| Multi EXT | bag i ~~ bag q | -0.195 | 0.022 | < 0.01 | -0.595 | Male |
| Multi INT | bag i ~~ bag q | -0.181 | 0.024 | < 0.01 | -0.598 | Female |
| Multi INT | bag i ~~ bag q | -0.195 | 0.022 | < 0.01 | -0.597 | Male |
| T1 EXT | bag i ~~ bag q | -0.31 | 0.03 | < 0.01 | -0.618 | Female |
| T1 EXT | bag i ~~ bag q | -0.237 | 0.026 | < 0.01 | -0.624 | Male |
| T1 INT | bag i ~~ bag q | -0.317 | 0.031 | < 0.01 | -0.622 | Female |
| T1 INT | bag i ~~ bag q | -0.236 | 0.026 | < 0.01 | -0.627 | Male |
| dMRI EXT | bag i ~~ bag q | -0.24 | 0.019 | < 0.01 | -0.707 | Female |
| dMRI EXT | bag i ~~ bag q | -0.24 | 0.019 | < 0.01 | -0.625 | Male |
| dMRI INT | bag i ~~ bag q | -0.228 | 0.027 | < 0.01 | -0.723 | Female |
| dMRI INT | bag i ~~ bag q | -0.249 | 0.027 | < 0.01 | -0.624 | Male |
| Multi EXT | bag i ~~ bag s | 0.092 | 0.021 | < 0.01 | 0.2 | Female |
| Multi EXT | bag i ~~ bag s | 0.108 | 0.019 | < 0.01 | 0.263 | Male |
| Multi INT | bag i ~~ bag s | 0.09 | 0.021 | < 0.01 | 0.195 | Female |
| Multi INT | bag i ~~ bag s | 0.107 | 0.019 | < 0.01 | 0.263 | Male |
| T1 EXT | bag i ~~ bag s | 0.146 | 0.027 | < 0.01 | 0.228 | Female |
| T1 EXT | bag i ~~ bag s | 0.134 | 0.024 | < 0.01 | 0.248 | Male |
| T1 INT | bag i ~~ bag s | 0.14 | 0.027 | < 0.01 | 0.22 | Female |
| T1 INT | bag i ~~ bag s | 0.134 | 0.024 | < 0.01 | 0.25 | Male |
| dMRI EXT | bag i ~~ bag s | 0.107 | 0.016 | < 0.01 | 0.228 | Female |
| dMRI EXT | bag i ~~ bag s | 0.107 | 0.016 | < 0.01 | 0.213 | Male |
| dMRI INT | bag i ~~ bag s | 0.125 | 0.024 | < 0.01 | 0.276 | Female |
| dMRI INT | bag i ~~ bag s | 0.092 | 0.023 | < 0.01 | 0.179 | Male |
| rsfMRI EXT | bag i ~~ bag s | 0.145 | 0.062 | 0.02 | 0.276 | Female |
| rsfMRI EXT | bag i ~~ bag s | 0.199 | 0.058 | < 0.01 | 0.324 | Male |
| rsfMRI INT | bag i ~~ bag s | 0.145 | 0.063 | 0.02 | 0.281 | Female |
| rsfMRI INT | bag i ~~ bag s | 0.2 | 0.059 | < 0.01 | 0.324 | Male |
| Multi EXT | bag i ~~ ext q | -0.009 | 0.003 | < 0.01 | -0.139 | Female |
| Multi EXT | bag i ~~ ext q | -0.003 | 0.003 | 0.23 | -0.059 | Male |
| T1 EXT | bag i ~~ ext q | -0.007 | 0.003 | 0.03 | -0.085 | Female |
| T1 EXT | bag i ~~ ext q | -0.004 | 0.003 | 0.15 | -0.06 | Male |
| dMRI EXT | bag i ~~ ext q | -0.004 | 0.002 | 0.08 | -0.056 | Female |
| dMRI EXT | bag i ~~ ext q | -0.004 | 0.002 | 0.08 | -0.057 | Male |
| rsfMRI EXT | bag i ~~ ext q | -0.009 | 0.003 | < 0.01 | -0.143 | Female |
| rsfMRI EXT | bag i ~~ ext q | 0.002 | 0.003 | 0.53 | 0.032 | Male |
| Multi EXT | bag i ~~ ext s | -0.005 | 0.009 | 0.56 | -0.021 | Female |
| Multi EXT | bag i ~~ ext s | 0.012 | 0.008 | 0.13 | 0.053 | Male |
| T1 EXT | bag i ~~ ext s | -0.008 | 0.01 | 0.43 | -0.025 | Female |
| T1 EXT | bag i ~~ ext s | 0.005 | 0.009 | 0.63 | 0.015 | Male |
| dMRI EXT | bag i ~~ ext s | -0.004 | 0.006 | 0.52 | -0.015 | Female |
| dMRI EXT | bag i ~~ ext s | -0.004 | 0.006 | 0.52 | -0.016 | Male |
| rsfMRI EXT | bag i ~~ ext s | 0.007 | 0.011 | 0.51 | 0.028 | Female |
| rsfMRI EXT | bag i ~~ ext s | -0.002 | 0.01 | 0.84 | -0.007 | Male |
| Multi INT | bag i ~~ int q | -0.011 | 0.004 | < 0.01 | -0.11 | Female |
| Multi INT | bag i ~~ int q | -0.002 | 0.003 | 0.57 | -0.024 | Male |
| T1 INT | bag i ~~ int q | -0.005 | 0.004 | 0.28 | -0.039 | Female |
| T1 INT | bag i ~~ int q | 0.001 | 0.003 | 0.78 | 0.01 | Male |
| dMRI INT | bag i ~~ int q | -0.008 | 0.004 | 0.07 | -0.076 | Female |
| dMRI INT | bag i ~~ int q | -0.001 | 0.003 | 0.75 | -0.014 | Male |
| rsfMRI INT | bag i ~~ int q | -0.007 | 0.005 | 0.19 | -0.063 | Female |
| rsfMRI INT | bag i ~~ int q | 0.002 | 0.004 | 0.64 | 0.022 | Male |
| Multi INT | bag i ~~ int s | -0.012 | 0.014 | 0.40 | -0.029 | Female |
| Multi INT | bag i ~~ int s | 0.016 | 0.01 | 0.10 | 0.053 | Male |
| T1 INT | bag i ~~ int s | -0.002 | 0.015 | 0.88 | -0.004 | Female |
| T1 INT | bag i ~~ int s | 0.014 | 0.011 | 0.22 | 0.038 | Male |
| dMRI INT | bag i ~~ int s | 0 | 0.014 | 0.99 | 0 | Female |
| dMRI INT | bag i ~~ int s | -0.003 | 0.011 | 0.79 | -0.009 | Male |
| rsfMRI INT | bag i ~~ int s | 0.005 | 0.016 | 0.75 | 0.013 | Female |
| rsfMRI INT | bag i ~~ int s | 0.011 | 0.014 | 0.43 | 0.031 | Male |
| Multi EXT | bag i ~~ out i | 0.114 | 0.053 | 0.03 | 0.063 | Female |
| Multi EXT | bag i ~~ out i | 0.119 | 0.05 | 0.02 | 0.069 | Male |
| Multi INT | bag i ~~ out i | 0.098 | 0.068 | 0.15 | 0.042 | Female |
| Multi INT | bag i ~~ out i | 0.099 | 0.049 | 0.04 | 0.058 | Male |
| T1 EXT | bag i ~~ out i | 0.18 | 0.061 | < 0.01 | 0.076 | Female |
| T1 EXT | bag i ~~ out i | 0.092 | 0.057 | 0.11 | 0.041 | Male |
| T1 INT | bag i ~~ out i | 0.105 | 0.078 | 0.18 | 0.034 | Female |
| T1 INT | bag i ~~ out i | 0.02 | 0.056 | 0.72 | 0.009 | Male |
| dMRI EXT | bag i ~~ out i | 0.032 | 0.04 | 0.42 | 0.016 | Female |
| dMRI EXT | bag i ~~ out i | 0.032 | 0.04 | 0.42 | 0.016 | Male |
| dMRI INT | bag i ~~ out i | 0.039 | 0.074 | 0.60 | 0.015 | Female |
| dMRI INT | bag i ~~ out i | -0.024 | 0.054 | 0.66 | -0.012 | Male |
| rsfMRI EXT | bag i ~~ out i | 0.166 | 0.065 | 0.01 | 0.086 | Female |
| rsfMRI EXT | bag i ~~ out i | -0.006 | 0.062 | 0.93 | -0.003 | Male |
| rsfMRI INT | bag i ~~ out i | 0.116 | 0.083 | 0.16 | 0.047 | Female |
| rsfMRI INT | bag i ~~ out i | -0.052 | 0.064 | 0.41 | -0.025 | Male |
| Multi EXT | bag q ~~ bag q | 0.068 | 0.014 | < 0.01 | 1 | Female |
| Multi EXT | bag q ~~ bag q | 0.08 | 0.014 | < 0.01 | 1 | Male |
| Multi INT | bag q ~~ bag q | 0.068 | 0.014 | < 0.01 | 1 | Female |
| Multi INT | bag q ~~ bag q | 0.08 | 0.014 | < 0.01 | 1 | Male |
| T1 EXT | bag q ~~ bag q | 0.108 | 0.018 | < 0.01 | 1 | Female |
| T1 EXT | bag q ~~ bag q | 0.068 | 0.016 | < 0.01 | 1 | Male |
| T1 INT | bag q ~~ bag q | 0.112 | 0.018 | < 0.01 | 1 | Female |
| T1 INT | bag q ~~ bag q | 0.067 | 0.016 | < 0.01 | 1 | Male |
| dMRI EXT | bag q ~~ bag q | 0.071 | 0.015 | < 0.01 | 1 | Female |
| dMRI EXT | bag q ~~ bag q | 0.093 | 0.014 | < 0.01 | 1 | Male |
| dMRI INT | bag q ~~ bag q | 0.062 | 0.017 | < 0.01 | 1 | Female |
| dMRI INT | bag q ~~ bag q | 0.1 | 0.016 | < 0.01 | 1 | Male |
| Multi EXT | bag q ~~ bag s | 0.01 | 0.02 | 0.60 | 0.098 | Female |
| Multi EXT | bag q ~~ bag s | 0.01 | 0.019 | 0.59 | 0.101 | Male |
| Multi INT | bag q ~~ bag s | 0.011 | 0.02 | 0.58 | 0.105 | Female |
| Multi INT | bag q ~~ bag s | 0.009 | 0.019 | 0.63 | 0.092 | Male |
| T1 EXT | bag q ~~ bag s | -0.018 | 0.026 | 0.50 | -0.128 | Female |
| T1 EXT | bag q ~~ bag s | -0.007 | 0.024 | 0.75 | -0.077 | Male |
| T1 INT | bag q ~~ bag s | -0.017 | 0.026 | 0.52 | -0.12 | Female |
| T1 INT | bag q ~~ bag s | -0.008 | 0.024 | 0.75 | -0.08 | Male |
| dMRI EXT | bag q ~~ bag s | 0.009 | 0.015 | 0.55 | 0.094 | Female |
| dMRI EXT | bag q ~~ bag s | 0.009 | 0.015 | 0.55 | 0.076 | Male |
| dMRI INT | bag q ~~ bag s | -0.017 | 0.023 | 0.45 | -0.194 | Female |
| dMRI INT | bag q ~~ bag s | 0.031 | 0.021 | 0.14 | 0.237 | Male |
| Multi EXT | bag q ~~ ext i | -0.019 | 0.021 | 0.35 | -0.048 | Female |
| Multi EXT | bag q ~~ ext i | -0.051 | 0.02 | 0.01 | -0.121 | Male |
| T1 EXT | bag q ~~ ext i | -0.1 | 0.023 | < 0.01 | -0.194 | Female |
| T1 EXT | bag q ~~ ext i | -0.068 | 0.02 | < 0.01 | -0.167 | Male |
| dMRI EXT | bag q ~~ ext i | -0.024 | 0.016 | 0.13 | -0.056 | Female |
| dMRI EXT | bag q ~~ ext i | -0.024 | 0.016 | 0.13 | -0.049 | Male |
| Multi EXT | bag q ~~ ext s | 0.005 | 0.003 | 0.15 | 0.087 | Female |
| Multi EXT | bag q ~~ ext s | 0.001 | 0.003 | 0.84 | 0.012 | Male |
| T1 EXT | bag q ~~ ext s | 0.006 | 0.004 | 0.11 | 0.082 | Female |
| T1 EXT | bag q ~~ ext s | -0.001 | 0.003 | 0.76 | -0.019 | Male |
| dMRI EXT | bag q ~~ ext s | 0.001 | 0.003 | 0.57 | 0.026 | Female |
| dMRI EXT | bag q ~~ ext s | 0.001 | 0.003 | 0.57 | 0.024 | Male |
| Multi INT | bag q ~~ int i | -0.068 | 0.026 | < 0.01 | -0.131 | Female |
| Multi INT | bag q ~~ int i | -0.057 | 0.019 | < 0.01 | -0.136 | Male |
| T1 INT | bag q ~~ int i | -0.073 | 0.029 | 0.01 | -0.109 | Female |
| T1 INT | bag q ~~ int i | -0.018 | 0.02 | 0.36 | -0.045 | Male |
| dMRI INT | bag q ~~ int i | -0.021 | 0.028 | 0.46 | -0.041 | Female |
| dMRI INT | bag q ~~ int i | -0.014 | 0.021 | 0.50 | -0.029 | Male |
| Multi INT | bag q ~~ int s | 0.003 | 0.005 | 0.57 | 0.034 | Female |
| Multi INT | bag q ~~ int s | 0.001 | 0.004 | 0.81 | 0.014 | Male |
| T1 INT | bag q ~~ int s | -0.001 | 0.005 | 0.90 | -0.007 | Female |
| T1 INT | bag q ~~ int s | -0.001 | 0.004 | 0.76 | -0.018 | Male |
| dMRI INT | bag q ~~ int s | 0.003 | 0.006 | 0.55 | 0.041 | Female |
| dMRI INT | bag q ~~ int s | 0 | 0.004 | 0.97 | -0.002 | Male |
| Multi EXT | bag q ~~ out q | 0.003 | 0.001 | < 0.01 | 0.243 | Female |
| Multi EXT | bag q ~~ out q | 0.002 | 0.001 | 0.12 | 0.124 | Male |
| Multi INT | bag q ~~ out q | 0.005 | 0.001 | < 0.01 | 0.228 | Female |
| Multi INT | bag q ~~ out q | 0.002 | 0.001 | 0.20 | 0.09 | Male |
| T1 EXT | bag q ~~ out q | 0.006 | 0.001 | < 0.01 | 0.317 | Female |
| T1 EXT | bag q ~~ out q | 0.003 | 0.001 | < 0.01 | 0.236 | Male |
| T1 INT | bag q ~~ out q | 0.004 | 0.002 | < 0.01 | 0.152 | Female |
| T1 INT | bag q ~~ out q | 0 | 0.001 | 0.98 | -0.002 | Male |
| dMRI EXT | bag q ~~ out q | 0.002 | 0.001 | < 0.01 | 0.157 | Female |
| dMRI EXT | bag q ~~ out q | 0.002 | 0.001 | < 0.01 | 0.139 | Male |
| dMRI INT | bag q ~~ out q | 0.004 | 0.002 | < 0.01 | 0.213 | Female |
| dMRI INT | bag q ~~ out q | 0 | 0.001 | 0.99 | -0.001 | Male |
| Multi EXT | bag s ~~ bag s | 0.159 | 0.02 | < 0.01 | 1 | Female |
| Multi EXT | bag s ~~ bag s | 0.127 | 0.019 | < 0.01 | 1 | Male |
| Multi INT | bag s ~~ bag s | 0.157 | 0.02 | < 0.01 | 1 | Female |
| Multi INT | bag s ~~ bag s | 0.125 | 0.019 | < 0.01 | 1 | Male |
| T1 EXT | bag s ~~ bag s | 0.176 | 0.023 | < 0.01 | 1 | Female |
| T1 EXT | bag s ~~ bag s | 0.138 | 0.021 | < 0.01 | 1 | Male |
| T1 INT | bag s ~~ bag s | 0.175 | 0.023 | < 0.01 | 1 | Female |
| T1 INT | bag s ~~ bag s | 0.137 | 0.021 | < 0.01 | 1 | Male |
| dMRI EXT | bag s ~~ bag s | 0.137 | 0.021 | < 0.01 | 1 | Female |
| dMRI EXT | bag s ~~ bag s | 0.159 | 0.02 | < 0.01 | 1 | Male |
| dMRI INT | bag s ~~ bag s | 0.128 | 0.023 | < 0.01 | 1 | Female |
| dMRI INT | bag s ~~ bag s | 0.167 | 0.02 | < 0.01 | 1 | Male |
| rsfMRI EXT | bag s ~~ bag s | 0.174 | 0.073 | < 0.01 | 1 | Female |
| rsfMRI EXT | bag s ~~ bag s | 0.205 | 0.063 | < 0.01 | 1 | Male |
| rsfMRI INT | bag s ~~ bag s | 0.168 | 0.073 | 0.02 | 1 | Female |
| rsfMRI INT | bag s ~~ bag s | 0.206 | 0.063 | < 0.01 | 1 | Male |
| Multi EXT | bag s ~~ ext i | -0.013 | 0.022 | 0.56 | -0.021 | Female |
| Multi EXT | bag s ~~ ext i | -0.005 | 0.021 | 0.81 | -0.01 | Male |
| T1 EXT | bag s ~~ ext i | -0.058 | 0.023 | 0.01 | -0.088 | Female |
| T1 EXT | bag s ~~ ext i | -0.056 | 0.021 | < 0.01 | -0.096 | Male |
| dMRI EXT | bag s ~~ ext i | -0.021 | 0.017 | 0.23 | -0.036 | Female |
| dMRI EXT | bag s ~~ ext i | -0.021 | 0.017 | 0.23 | -0.033 | Male |
| rsfMRI EXT | bag s ~~ ext i | -0.045 | 0.045 | 0.32 | -0.07 | Female |
| rsfMRI EXT | bag s ~~ ext i | -0.046 | 0.041 | 0.26 | -0.068 | Male |
| Multi EXT | bag s ~~ ext q | 0 | 0.001 | 0.81 | 0.013 | Female |
| Multi EXT | bag s ~~ ext q | 0.001 | 0.001 | 0.64 | 0.03 | Male |
| T1 EXT | bag s ~~ ext q | 0.002 | 0.001 | 0.10 | 0.089 | Female |
| T1 EXT | bag s ~~ ext q | 0.002 | 0.001 | 0.12 | 0.094 | Male |
| dMRI EXT | bag s ~~ ext q | 0.001 | 0.001 | 0.13 | 0.072 | Female |
| dMRI EXT | bag s ~~ ext q | 0.001 | 0.001 | 0.13 | 0.067 | Male |
| rsfMRI EXT | bag s ~~ ext q | -0.003 | 0.002 | 0.25 | -0.128 | Female |
| rsfMRI EXT | bag s ~~ ext q | 0.002 | 0.002 | 0.38 | 0.093 | Male |
| Multi INT | bag s ~~ int i | -0.041 | 0.027 | 0.13 | -0.051 | Female |
| Multi INT | bag s ~~ int i | -0.03 | 0.021 | 0.16 | -0.057 | Male |
| T1 INT | bag s ~~ int i | -0.028 | 0.028 | 0.32 | -0.033 | Female |
| T1 INT | bag s ~~ int i | -0.048 | 0.021 | 0.02 | -0.084 | Male |
| dMRI INT | bag s ~~ int i | -0.022 | 0.03 | 0.47 | -0.029 | Female |
| dMRI INT | bag s ~~ int i | -0.041 | 0.024 | 0.09 | -0.065 | Male |
| rsfMRI INT | bag s ~~ int i | 0.061 | 0.058 | 0.30 | 0.075 | Female |
| rsfMRI INT | bag s ~~ int i | -0.066 | 0.044 | 0.13 | -0.096 | Male |
| Multi INT | bag s ~~ int q | 0.001 | 0.001 | 0.36 | 0.04 | Female |
| Multi INT | bag s ~~ int q | 0.001 | 0.001 | 0.26 | 0.07 | Male |
| T1 INT | bag s ~~ int q | 0.001 | 0.002 | 0.46 | 0.035 | Female |
| T1 INT | bag s ~~ int q | 0.001 | 0.001 | 0.32 | 0.053 | Male |
| dMRI INT | bag s ~~ int q | 0.002 | 0.002 | 0.24 | 0.07 | Female |
| dMRI INT | bag s ~~ int q | 0.002 | 0.001 | 0.24 | 0.065 | Male |
| rsfMRI INT | bag s ~~ int q | -0.004 | 0.003 | 0.29 | -0.105 | Female |
| rsfMRI INT | bag s ~~ int q | 0.002 | 0.003 | 0.49 | 0.068 | Male |
| Multi EXT | bag s ~~ out s | 0.009 | 0.004 | 0.02 | 0.103 | Female |
| Multi EXT | bag s ~~ out s | 0.007 | 0.003 | 0.03 | 0.107 | Male |
| Multi INT | bag s ~~ out s | 0.007 | 0.006 | 0.19 | 0.054 | Female |
| Multi INT | bag s ~~ out s | 0.008 | 0.005 | 0.09 | 0.085 | Male |
| T1 EXT | bag s ~~ out s | 0.003 | 0.004 | 0.41 | 0.034 | Female |
| T1 EXT | bag s ~~ out s | 0 | 0.004 | 0.90 | -0.006 | Male |
| T1 INT | bag s ~~ out s | -0.003 | 0.005 | 0.60 | -0.021 | Female |
| T1 INT | bag s ~~ out s | 0.006 | 0.004 | 0.12 | 0.068 | Male |
| dMRI EXT | bag s ~~ out s | 0.004 | 0.003 | 0.16 | 0.049 | Female |
| dMRI EXT | bag s ~~ out s | 0.004 | 0.003 | 0.16 | 0.048 | Male |
| dMRI INT | bag s ~~ out s | 0.006 | 0.006 | 0.31 | 0.054 | Female |
| dMRI INT | bag s ~~ out s | 0.008 | 0.005 | 0.10 | 0.072 | Male |
| rsfMRI EXT | bag s ~~ out s | 0.011 | 0.008 | 0.18 | 0.125 | Female |
| rsfMRI EXT | bag s ~~ out s | 0.011 | 0.007 | 0.12 | 0.116 | Male |
| rsfMRI INT | bag s ~~ out s | 0.027 | 0.011 | 0.02 | 0.198 | Female |
| rsfMRI INT | bag s ~~ out s | 0.013 | 0.009 | 0.153 | 0.112 | Male |
| Multi EXT | ext i ~~ ext i | 2.39 | 0.102 | < 0.01 | 1 | Female |
| Multi EXT | ext i ~~ ext i | 2.24 | 0.096 | < 0.01 | 1 | Male |
| T1 EXT | ext i ~~ ext i | 2.438 | 0.091 | < 0.01 | 1 | Female |
| T1 EXT | ext i ~~ ext i | 2.417 | 0.091 | < 0.01 | 1 | Male |
| dMRI EXT | ext i ~~ ext i | 2.484 | 0.107 | < 0.01 | 1 | Female |
| dMRI EXT | ext i ~~ ext i | 2.521 | 0.098 | < 0.01 | 1 | Male |
| rsfMRI EXT | ext i ~~ ext i | 2.392 | 0.104 | < 0.01 | 1 | Female |
| rsfMRI EXT | ext i ~~ ext i | 2.302 | 0.084 | < 0.01 | 1 | Male |
| Multi EXT | ext q ~~ ext q | 0.003 | 0 | < 0.01 | 1 | Female |
| Multi EXT | ext q ~~ ext q | 0.002 | 0 | < 0.01 | 1 | Male |
| T1 EXT | ext q ~~ ext q | 0.003 | 0 | < 0.01 | 1 | Female |
| T1 EXT | ext q ~~ ext q | 0.003 | 0 | < 0.01 | 1 | Male |
| dMRI EXT | ext q ~~ ext q | 0.003 | 0 | < 0.01 | 1 | Female |
| dMRI EXT | ext q ~~ ext q | 0.003 | 0 | < 0.01 | 1 | Male |
| rsfMRI EXT | ext q ~~ ext q | 0.003 | 0 | < 0.01 | 1 | Female |
| rsfMRI EXT | ext q ~~ ext q | 0.002 | 0 | < 0.01 | 1 | Male |
| Multi EXT | ext q ~~ ext s | 0.003 | 0.001 | < 0.01 | 0.226 | Female |
| Multi EXT | ext q ~~ ext s | 0.001 | 0.001 | 0.05 | 0.13 | Male |
| T1 EXT | ext q ~~ ext s | 0.003 | 0.001 | < 0.01 | 0.272 | Female |
| T1 EXT | ext q ~~ ext s | 0.002 | 0.001 | < 0.01 | 0.173 | Male |
| dMRI EXT | ext q ~~ ext s | 0.002 | 0.001 | < 0.01 | 0.177 | Female |
| dMRI EXT | ext q ~~ ext s | 0.002 | 0.001 | < 0.01 | 0.188 | Male |
| rsfMRI EXT | ext q ~~ ext s | 0.003 | 0.001 | < 0.01 | 0.27 | Female |
| rsfMRI EXT | ext q ~~ ext s | 0.002 | 0.001 | < 0.01 | 0.223 | Male |
| Multi EXT | ext s ~~ ext s | 0.048 | 0.003 | < 0.01 | 1 | Female |
| Multi EXT | ext s ~~ ext s | 0.038 | 0.003 | < 0.01 | 1 | Male |
| T1 EXT | ext s ~~ ext s | 0.045 | 0.003 | < 0.01 | 1 | Female |
| T1 EXT | ext s ~~ ext s | 0.044 | 0.003 | < 0.01 | 1 | Male |
| dMRI EXT | ext s ~~ ext s | 0.047 | 0.003 | < 0.01 | 1 | Female |
| dMRI EXT | ext s ~~ ext s | 0.042 | 0.003 | < 0.01 | 1 | Male |
| rsfMRI EXT | ext s ~~ ext s | 0.047 | 0.003 | < 0.01 | 1 | Female |
| rsfMRI EXT | ext s ~~ ext s | 0.041 | 0.003 | < 0.01 | 1 | Male |
| Multi INT | int i ~~ int i | 4.009 | 0.181 | < 0.01 | 1 | Female |
| Multi INT | int i ~~ int i | 2.143 | 0.119 | < 0.01 | 1 | Male |
| T1 INT | int i ~~ int i | 4.005 | 0.166 | < 0.01 | 1 | Female |
| T1 INT | int i ~~ int i | 2.399 | 0.125 | < 0.01 | 1 | Male |
| dMRI INT | int i ~~ int i | 4.205 | 0.185 | < 0.01 | 1 | Female |
| dMRI INT | int i ~~ int i | 2.403 | 0.132 | < 0.01 | 1 | Male |
| rsfMRI INT | int i ~~ int i | 3.873 | 0.157 | < 0.01 | 1 | Female |
| rsfMRI INT | int i ~~ int i | 2.328 | 0.116 | < 0.01 | 1 | Male |
| Multi INT | int q ~~ int q | 0.007 | 0.001 | < 0.01 | 1 | Female |
| Multi INT | int q ~~ int q | 0.004 | 0 | < 0.01 | 1 | Male |
| T1 INT | int q ~~ int q | 0.006 | 0.001 | < 0.01 | 1 | Female |
| T1 INT | int q ~~ int q | 0.004 | 0 | < 0.01 | 1 | Male |
| dMRI INT | int q ~~ int q | 0.006 | 0.001 | < 0.01 | 1 | Female |
| dMRI INT | int q ~~ int q | 0.004 | 0 | < 0.01 | 1 | Male |
| rsfMRI INT | int q ~~ int q | 0.007 | 0.001 | < 0.01 | 1 | Female |
| rsfMRI INT | int q ~~ int q | 0.004 | 0 | < 0.01 | 1 | Male |
| Multi INT | int q ~~ int s | 0.01 | 0.002 | < 0.01 | 0.327 | Female |
| Multi INT | int q ~~ int s | 0.004 | 0.001 | < 0.01 | 0.263 | Male |
| T1 INT | int q ~~ int s | 0.007 | 0.001 | < 0.01 | 0.271 | Female |
| T1 INT | int q ~~ int s | 0.004 | 0.001 | < 0.01 | 0.252 | Male |
| dMRI INT | int q ~~ int s | 0.007 | 0.001 | < 0.01 | 0.285 | Female |
| dMRI INT | int q ~~ int s | 0.003 | 0.001 | < 0.01 | 0.197 | Male |
| rsfMRI INT | int q ~~ int s | 0.008 | 0.001 | < 0.01 | 0.28 | Female |
| rsfMRI INT | int q ~~ int s | 0.003 | 0.001 | < 0.01 | 0.184 | Male |
| Multi INT | int s ~~ int s | 0.121 | 0.007 | < 0.01 | 1 | Female |
| Multi INT | int s ~~ int s | 0.069 | 0.005 | < 0.01 | 1 | Male |
| T1 INT | int s ~~ int s | 0.102 | 0.006 | < 0.01 | 1 | Female |
| T1 INT | int s ~~ int s | 0.064 | 0.004 | < 0.01 | 1 | Male |
| dMRI INT | int s ~~ int s | 0.106 | 0.006 | < 0.01 | 1 | Female |
| dMRI INT | int s ~~ int s | 0.066 | 0.004 | < 0.01 | 1 | Male |
| rsfMRI INT | int s ~~ int s | 0.108 | 0.006 | < 0.01 | 1 | Female |
| rsfMRI INT | int s ~~ int s | 0.065 | 0.004 | < 0.01 | 1 | Male |
| Multi EXT | out i ~~ out q | -0.04 | 0.004 | < 0.01 | -0.478 | Female |
| Multi EXT | out i ~~ out q | -0.03 | 0.004 | < 0.01 | -0.415 | Male |
| Multi INT | out i ~~ out q | -0.07 | 0.008 | < 0.01 | -0.413 | Female |
| Multi INT | out i ~~ out q | -0.03 | 0.006 | < 0.01 | -0.346 | Male |
| T1 EXT | out i ~~ out q | -0.045 | 0.004 | < 0.01 | -0.513 | Female |
| T1 EXT | out i ~~ out q | -0.033 | 0.004 | < 0.01 | -0.423 | Male |
| T1 INT | out i ~~ out q | -0.073 | 0.007 | < 0.01 | -0.459 | Female |
| T1 INT | out i ~~ out q | -0.035 | 0.005 | < 0.01 | -0.366 | Male |
| dMRI EXT | out i ~~ out q | -0.037 | 0.003 | < 0.01 | -0.457 | Female |
| dMRI EXT | out i ~~ out q | -0.037 | 0.003 | < 0.01 | -0.459 | Male |
| dMRI INT | out i ~~ out q | -0.074 | 0.008 | < 0.01 | -0.458 | Female |
| dMRI INT | out i ~~ out q | -0.042 | 0.006 | < 0.01 | -0.436 | Male |
| rsfMRI EXT | out i ~~ out q | -0.04 | 0.004 | < 0.01 | -0.501 | Female |
| rsfMRI EXT | out i ~~ out q | -0.031 | 0.004 | < 0.01 | -0.42 | Male |
| rsfMRI INT | out i ~~ out q | -0.078 | 0.008 | < 0.01 | -0.472 | Female |
| rsfMRI INT | out i ~~ out q | -0.034 | 0.005 | < 0.01 | -0.371 | Male |
| Multi EXT | out i ~~ out s | 0.037 | 0.013 | < 0.01 | 0.11 | Female |
| Multi EXT | out i ~~ out s | -0.004 | 0.011 | 0.72 | -0.013 | Male |
| Multi INT | out i ~~ out s | 0.222 | 0.023 | < 0.01 | 0.319 | Female |
| Multi INT | out i ~~ out s | 0.055 | 0.016 | < 0.01 | 0.143 | Male |
| T1 EXT | out i ~~ out s | 0.026 | 0.011 | 0.01 | 0.078 | Female |
| T1 EXT | out i ~~ out s | -0.023 | 0.011 | 0.03 | -0.07 | Male |
| T1 INT | out i ~~ out s | 0.204 | 0.02 | < 0.01 | 0.318 | Female |
| T1 INT | out i ~~ out s | 0.029 | 0.014 | 0.04 | 0.073 | Male |
| dMRI EXT | out i ~~ out s | 0.015 | 0.009 | 0.09 | 0.045 | Female |
| dMRI EXT | out i ~~ out s | 0.015 | 0.009 | 0.09 | 0.047 | Male |
| dMRI INT | out i ~~ out s | 0.205 | 0.022 | < 0.01 | 0.306 | Female |
| dMRI INT | out i ~~ out s | 0.05 | 0.016 | < 0.01 | 0.125 | Male |
| rsfMRI EXT | out i ~~ out s | 0.023 | 0.012 | 0.05 | 0.069 | Female |
| rsfMRI EXT | out i ~~ out s | -0.015 | 0.011 | 0.16 | -0.049 | Male |
| rsfMRI INT | out i ~~ out s | 0.185 | 0.02 | < 0.01 | 0.287 | Female |
| rsfMRI INT | out i ~~ out s | 0.057 | 0.015 | < 0.01 | 0.146 | Male |

| **SI Table 9. Condensed BAG–symptom coupling estimates.** Summary of the latent covariance parameters of interest from the final BLGC models. | | | | |
| --- | --- | --- | --- | --- |
| **Model** | **Sex** | **Intercept** | **Quadratic** | **Slope** |
| Multi EXT | Female | 0.060 | 0.240* | 0.100 |
| Multi EXT | Male | 0.070 | 0.120 | 0.110 |
| Multi INT | Female | 0.040 | 0.230** | 0.050 |
| Multi INT | Male | 0.060 | 0.090 | 0.080 |
| T1 EXT | Female | 0.080* | 0.320*** | 0.030 |
| T1 EXT | Male | 0.040 | 0.240* | -0.010 |
| T1 INT | Female | 0.030 | 0.150* | -0.020 |
| T1 INT | Male | 0.010 | -0.000 | 0.070 |
| dMRI EXT | Female | 0.020 | 0.160* | 0.050 |
| dMRI EXT | Male | 0.020 | 0.140* | 0.050 |
| dMRI INT | Female | 0.010 | 0.210* | 0.050 |
| dMRI INT | Male | -0.010 | -0.000 | 0.070 |
| rsfMRI EXT | Female | 0.090* | NA | 0.120 |
| rsfMRI EXT | Male | -0.000 | NA | 0.120 |
| rsfMRI INT | Female | 0.050 | NA | 0.200 |
| rsfMRI INT | Male | -0.030 | NA | 0.110 |

Note: Asterisks indicate statistical significance of the within-sex BAG–symptom covariance after Benjamini–Hochberg false discovery rate correction across all primary coupling tests (**p* FDR < .05, ***p* FDR < .01, **p FDR < .001).

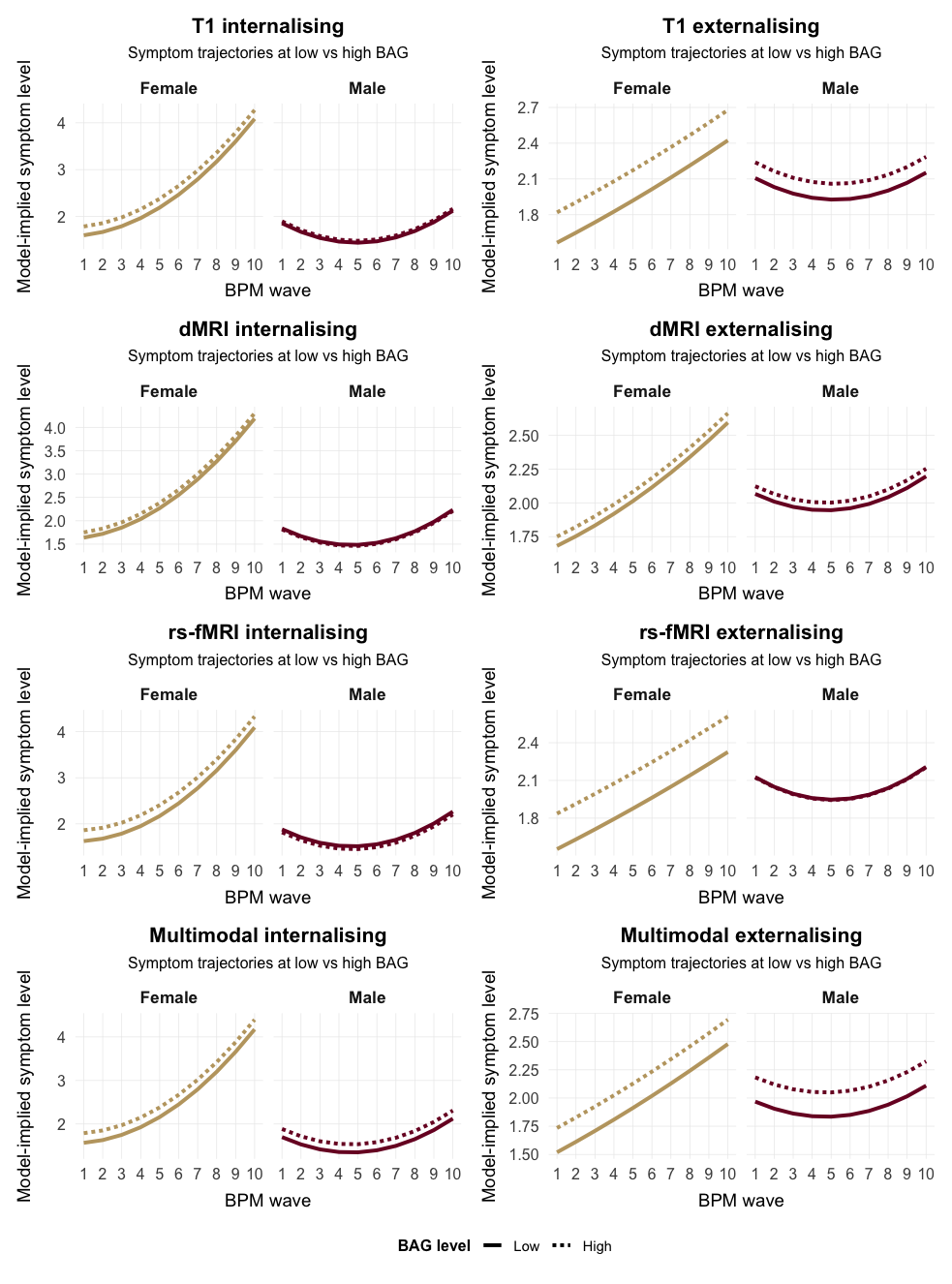

**Figure 13. Model-implied internalising and externalising trajectories over BPM waves at low versus high BAG.** For each modality (T1-weighted, dMRI, rs-fMRI, and multimodal) and symptom domain (internalising, externalising), lines show the model-implied symptom course across the 10 Brief Problem Monitor (BPM) waves derived from the fitted bivariate latent growth curve models. “Low” and “High” BAG are defined as −1 SD and +1 SD on the BAG intercept factor (bag i), respectively, and are translated into corresponding expected symptom levels via the estimated BAG–symptom intercept coupling (bag i ~~ int i / ext i), while symptom slope and quadratic factors are held at their latent means. Panels are stratified by sex.

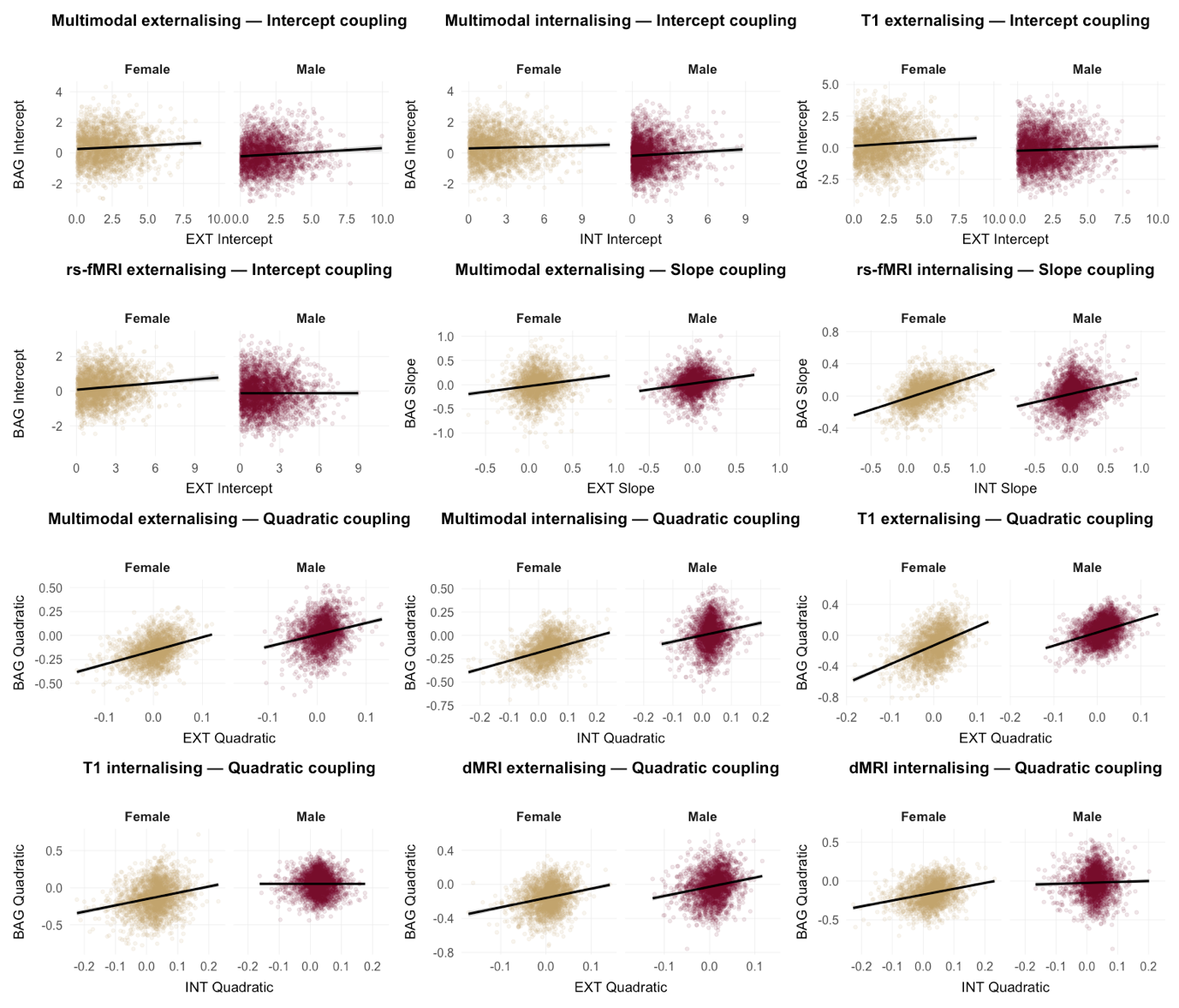

**Figure 14. Sex-stratified scatter plots of latent growth components for BAG and symptoms in significant couplings.** Each panel shows individual factor scores from the fitted bivariate latent growth curve models, with the BAG component on the y-axis and the corresponding symptom component on the x-axis (intercept, slope, or quadratic, depending on the coupling). Points represent participants’ estimated latent factor scores, and the overlaid line indicates the least-squares regression fit for visualization. Panels are stratified by sex. Only model × component combinations with significant BAG–symptom couplings (for at least one sex) are shown.

| **SI Table 10. Tests of sex differences in brain age gap–psychopathology couplings.** Targeted likelihood-ratio tests assessed sex differences in specific BAG–symptom couplings (intercept–intercept, slope–slope, and quadratic–quadratic, where applicable) across imaging modalities. *P*-values were corrected using the Benjamini–Hochberg false discovery rate (FDR). Quadratic couplings were not estimated in linear rs-fMRI models, and dMRI externalising models were pooled. | | | | | | | |
| --- | --- | --- | --- | --- | --- | --- | --- |
| **Model** | **Coupling** | **χ² diff** | **Df diff** | ***P* value** | ***P* FDR** | **Sig** | **Sig FDR** |
| Multi EXT | Intercept–Intercept | 0.006 | 1 | 0.939 | 0.995 |  |  |
| Multi EXT | Quadratic–Quadratic | 1.227 | 1 | 0.268 | 0.616 |  |  |
| Multi EXT | Slope–Slope | 0.094 | 1 | 0.759 | 0.995 |  |  |
| Multi INT | Intercept–Intercept | 0 | 1 | 0.995 | 0.995 |  |  |
| Multi INT | Quadratic–Quadratic | 3.535 | 1 | 0.06 | 0.285 |  |  |
| Multi INT | Slope–Slope | 0.004 | 1 | 0.949 | 0.995 |  |  |
| T1 EXT | Intercept–Intercept | 1.112 | 1 | 0.292 | 0.616 |  |  |
| T1 EXT | Quadratic–Quadratic | 2.838 | 1 | 0.092 | 0.338 |  |  |
| T1 EXT | Slope–Slope | 0.464 | 1 | 0.496 | 0.726 |  |  |
| T1 INT | Intercept–Intercept | 0.79 | 1 | 0.374 | 0.646 |  |  |
| T1 INT | Quadratic–Quadratic | 4.416 | 1 | 0.036 | 0.285 | * |  |
| T1 INT | Slope–Slope | 1.865 | 1 | 0.172 | 0.467 |  |  |
| dMRI INT | Intercept–Intercept | 0.462 | 1 | 0.497 | 0.726 |  |  |
| dMRI INT | Quadratic–Quadratic | 4.411 | 1 | 0.036 | 0.285 | * |  |
| dMRI INT | Slope–Slope | 0.026 | 1 | 0.871 | 0.995 |  |  |
| rsfMRI EXT | Intercept–Intercept | 3.676 | 1 | 0.055 | 0.285 |  |  |
| rsfMRI EXT | Quadratic–Quadratic | NA | NA | NA | NA |  |  |
| rsfMRI EXT | Slope–Slope | 0.003 | 1 | 0.954 | 0.995 |  |  |
| rsfMRI INT | Intercept–Intercept | 2.601 | 1 | 0.107 | 0.338 |  |  |
| rsfMRI INT | Quadratic–Quadratic | NA | NA | NA | NA |  |  |
| rsfMRI INT | Slope–Slope | 0.911 | 1 | 0.34 | 0.646 |  |  |

| **SI Table 11. SES sensitivity analysis**: BAG–symptom coupling estimates with socioeconomic status as a time-invariant covariate. Standardised cross-process covariance estimates (Est std), standard errors (SE), and p-values are reported for intercept–intercept, slope–slope, and quadratic–quadratic BAG–symptom couplings across all eight BLGC models, with a composite baseline socioeconomic status variable – derived from parental and partner income and educational attainment at baseline – included as a covariate regressed onto all latent growth factors. Results are shown separately for females and males where sex-stratified models were estimated, and as a single pooled estimate for the dMRI externalising model consistent with the primary analysis. | | | | | |
| --- | --- | --- | --- | --- | --- |
| **Model** | **Sex** | **Effect** | **Est std** | **SE** | ***P* value** |
| Multi EXT | Female | bag i ~~ out i | 0.051 | 0.052 | 0.085 |
| Multi EXT | Male | bag i ~~ out i | 0.052 | 0.049 | 0.072 |
| Multi INT | Female | bag i ~~ out i | 0.04 | 0.068 | 0.18 |
| Multi INT | Male | bag i ~~ out i | 0.049 | 0.049 | 0.092 |
| T1 EXT | Female | bag i ~~ out i | 0.064 | 0.06 | 0.013 |
| T1 EXT | Male | bag i ~~ out i | 0.032 | 0.056 | 0.2 |
| T1 INT | Female | bag i ~~ out i | 0.032 | 0.078 | 0.21 |
| T1 INT | Male | bag i ~~ out i | 0.003 | 0.055 | 0.9 |
| dMRI EXT | Female | bag i ~~ out i | 0.01 | 0.039 | 0.6 |
| dMRI EXT | Male | bag i ~~ out i | 0.01 | 0.039 | 0.6 |
| dMRI INT | Female | bag i ~~ out i | 0.013 | 0.074 | 0.65 |
| dMRI INT | Male | bag i ~~ out i | -0.014 | 0.054 | 0.61 |
| rsfMRI EXT | Female | bag i ~~ out i | 0.072 | 0.064 | 0.031 |
| rsfMRI EXT | Male | bag i ~~ out i | -0.028 | 0.061 | 0.35 |
| rsfMRI INT | Female | bag i ~~ out i | 0.041 | 0.082 | 0.21 |
| rsfMRI INT | Male | bag i ~~ out i | -0.04 | 0.063 | 0.2 |
| Multi EXT | Female | bag q ~~ out q | 0.235 | 0.001 | 0.003 |
| Multi EXT | Male | bag q ~~ out q | 0.12 | 0.001 | 0.14 |
| Multi INT | Female | bag q ~~ out q | 0.226 | 0.001 | 0 |
| Multi INT | Male | bag q ~~ out q | 0.096 | 0.001 | 0.18 |
| T1 EXT | Female | bag q ~~ out q | 0.315 | 0.001 | 0 |
| T1 EXT | Male | bag q ~~ out q | 0.227 | 0.001 | 0.005 |
| T1 INT | Female | bag q ~~ out q | 0.151 | 0.002 | 0.01 |
| T1 INT | Male | bag q ~~ out q | -0.003 | 0.001 | 0.97 |
| dMRI EXT | Female | bag q ~~ out q | 0.159 | 0.001 | 0.01 |
| dMRI EXT | Male | bag q ~~ out q | 0.138 | 0.001 | 0.01 |
| dMRI INT | Female | bag q ~~ out q | 0.216 | 0.002 | 0.009 |
| dMRI INT | Male | bag q ~~ out q | 0.004 | 0.001 | 0.95 |
| Multi EXT | Female | bag s ~~ out s | 0.101 | 0.004 | 0.023 |
| Multi EXT | Male | bag s ~~ out s | 0.108 | 0.003 | 0.034 |
| Multi INT | Female | bag s ~~ out s | 0.05 | 0.006 | 0.22 |
| Multi INT | Male | bag s ~~ out s | 0.085 | 0.005 | 0.094 |
| T1 EXT | Female | bag s ~~ out s | 0.032 | 0.004 | 0.43 |
| T1 EXT | Male | bag s ~~ out s | -0.011 | 0.003 | 0.81 |
| T1 INT | Female | bag s ~~ out s | -0.03 | 0.005 | 0.45 |
| T1 INT | Male | bag s ~~ out s | 0.062 | 0.004 | 0.16 |
| dMRI EXT | Female | bag s ~~ out s | 0.047 | 0.003 | 0.18 |
| dMRI EXT | Male | bag s ~~ out s | 0.046 | 0.003 | 0.18 |
| dMRI INT | Female | bag s ~~ out s | 0.042 | 0.006 | 0.43 |
| dMRI INT | Male | bag s ~~ out s | 0.071 | 0.005 | 0.1 |
| rsfMRI EXT | Female | bag s ~~ out s | 0.125 | 0.008 | 0.2 |
| rsfMRI EXT | Male | bag s ~~ out s | 0.107 | 0.007 | 0.16 |
| rsfMRI INT | Female | bag s ~~ out s | 0.16 | 0.011 | 0.062 |
| rsfMRI INT | Male | bag s ~~ out s | 0.112 | 0.009 | 0.16 |
